# High-resolution spatiotemporal estimates of total and wildfire PM_2.5_ in the contiguous United States, 2018-2024: Implications for population exposure and mortality burden

**DOI:** 10.64898/2026.11.11.26367854

**Authors:** Ningrui Liu, Yeonseung Cheong, Timothy Larson, Elena Austin, Edmund Seto

**Affiliations:** Department of Environmental and Occupational Health Sciences, University of Washington, Seattle, WA, USA; Department of Civil and Environmental Engineering, University of Washington, Seattle, WA, USA

**Keywords:** Fine particulate matter (PM_2.5_), Machine learning, Wildfire, Contiguous United States (CONUS), Mortality burden

## Abstract

Wildfire smoke has become an increasing public health concern in the contiguous United States (CONUS). Previous studies have provided high-resolution estimates of either total PM_2.5_ or wildfire-related PM_2.5_ during the past two decades, but few have focused on the most recent wildfire-intensive years, and jointly estimated both to quantify wildfire contributions to total PM_2.5_ exposure. This study developed a high-resolution machine learning framework to estimate daily 1-km concentrations of total and wildfire-related PM_2.5_ across the CONUS from 2018 to 2024. This framework incorporates chemical precursors and wildfire smoke density as predictors and relies on publicly accessible data sources enabling future updates. The model achieved high performance with cross-validated R^2^ of 0.72 and 0.89 for daily and annual estimates, respectively. Elevated total PM_2.5_ concentrations were observed along the west coast and in several metropolitan areas across the CONUS. Wildfire smoke contributed substantially to total PM_2.5_ concentrations during major wildfire years, accounting for approximately 35% of PM_2.5_ along the west coast in 2020, and over 25% in the eastern U.S. in 2023. Wildfire-related PM_2.5_ mortality accounted for 20% of PM_2.5_-attributable deaths along the west coast in 2020 (30 times that in 2019), and 24% in the northeastern U.S. in 2023 (14 times that in 2019). These findings highlight the growing role of wildfire smoke as a major driver of total PM_2.5_ exposure and associated health risks in both western and eastern U.S., underscoring the need for sustained nationwide public health interventions to reduce wildfire smoke exposure.

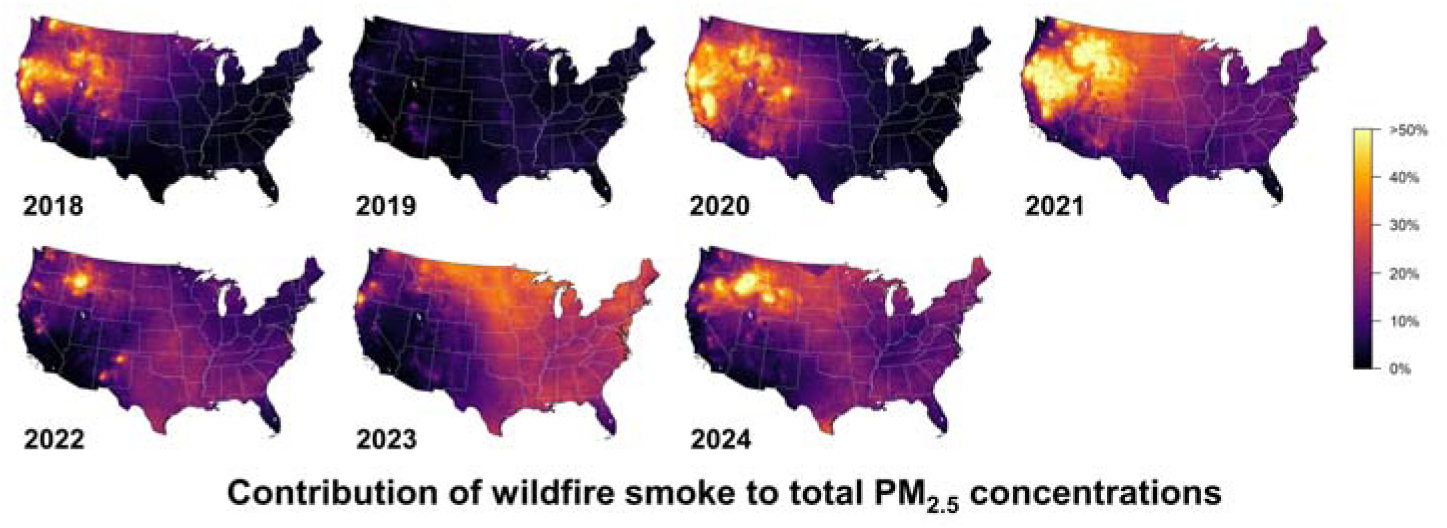
Graphical Abstract.

**Highlights:**

- Daily 1-km concentrations of total and wildfire PM_2.5_ were predicted for the CONUS.
- Elevated PM_2.5_ was observed along the U.S. west coast and metropolitan areas.
- Wildfire was a major driver of PM_2.5_ variability in both western and eastern U.S.
- Wildfire contributed over 20% to PM_2.5_-related mortality in severe wildfire years.

## 1 Introduction

Fine particulate matter (PM_2.5_) is associated with many adverse health outcomes, such as lung cancer, diabetes, and cardiovascular diseases. ^1–4^ The Global Burden of Disease (GBD) ranked PM_2.5_ as the third leading risk factors globally in 2023. ^5^ Therefore, accurately quantifying population exposure to PM_2.5_ at high spatial and temporal resolution is essential for assessing exposure disparities and associated health burdens. Higher spatial resolution can substantially reduce exposure misclassification in health risk assessment and environmental epidemiological studies. Higher temporal resolution can also help identify short-term peak exposures that would be obscured in annual averages. Due to stringent regulation of anthropogenic emissions in the United States (U.S.), ambient PM_2.5_ concentrations have shown an overall decreasing trend over the past two decades. ^6^ However, increasingly severe wildfire events in recent years have dramatically elevated short-term PM_2.5_ exposure at the daily scale. ^7^ Such episodic pollution events cannot be adequately captured by annual average estimates.

Previous studies have applied machine learning (ML) algorithms to estimate high-resolution ambient ground-level PM_2.5_ concentrations in the U.S. As one of the pioneering and most widely cited studies, Di et al. developed an ensemble model combining random forest, gradient boosting, and neural networks to estimate daily average PM_2.5_ concentrations at 1-km spatial resolution across the contiguous United States (CONUS) from 2000 to 2015. ^8^ Their model incorporated satellite-derived aerosol optical depth (AOD), chemical transport model (CTM) simulations, meteorological variables, and land-use characteristics as predictors. ^8^ Subsequent studies have adopted similar frameworks to estimate 1-km daily PM_2.5_ concentrations across the U.S. for different periods, using slightly different predictor sets and ML algorithms, such as deep forest and extreme gradient boosting (XGBoost). ^9–16^ However, most existing studies either focused on periods before 2020 or trained models over long historical timeframes starting in the early 2000s, when ambient PM_2.5_ concentrations were primarily driven by anthropogenic emissions. ^8–10, 12–19^ In contrast, few studies have specifically focused on the most recent decade with substantial wildfire smoke impacts in the U.S. ^11, 20^ Among the studies covering these wildfire-influenced years, Wei et al. developed a global model, which may not fully capture U.S.-specific emission patterns, regulatory environments, and wildfire characteristics. ^20^ Moreover, few studies explicitly incorporated wildfire-related predictors to characterize the complex relationships between wildfire activity and PM_2.5_ concentrations. ^14, 16, 21^

As wildfire smoke has become the major driver of ambient PM_2.5_ in U.S. recently, many studies have attempted to apportion total PM_2.5_ concentrations to wildfire and non-wildfire sources. The first type of studies has relied on fire contributions simulated by CTMs. Some of these studies directly derived wildfire-attributable PM_2.5_ using CTMs with fire emission inventories, ^22, 23^ while others first obtained the proportion of CTM-based wildfire PM_2.5_ in CTM-based total PM_2.5_ and then applied the proportion to total PM_2.5_ concentrations predicted from ML models. ^24, 25^ Although these CTM-based approaches offer strong physical interpretability, they may introduce substantial uncertainty in wildfire attribution due to uncertainties in fire emission inventories and CTM simulations, as the attribution is not directly constrained by ground-level observations. As a complement to CTM-based approaches, the second type of studies has leveraged statistical approaches to partition wildfire and non-wildfire air pollutants based on ground-level and satellite-based data sources. ^7, 16, 26–29^ One widely used statistical approach, developed by Burke et al., integrated ground-level PM_2.5_ measurements with smoke plume data to estimate the difference between observed total PM_2.5_ and counterfactual non-wildfire PM_2.5_ concentrations. ^7^ This framework was further extended by Childs et al., who incorporated these wildfire PM_2.5_ estimates into a ML model to produce 10-km daily estimates of wildfire PM_2.5_ concentrations across the CONUS. ^28^ Although the smoke plume boundaries drawn by National Oceanic and Atmospheric Administration (NOAA) analysts used in this approach may lead to bias in wildfire attribution, this approach has been well validated against ground-level observations and can be extended to large spatial scales due to relatively low computation costs. ^28^ Nevertheless, a gap remains in the proportion of wildfire PM_2.5_ in total PM_2.5_ across the U.S., ^16^ particularly during recent wildfire-dominated years.

To address these critical gaps in recent wildfire-era exposure characterization, this study focuses on the wildfire-influenced period after 2018 and incorporates wildfire-related predictors into a machine learning framework. Specifically, the objectives of this study are to (1) provide accurate 1-km daily estimates of ambient PM_2.5_ concentrations across the CONUS from 2018 to 2024; (2) estimate 1-km daily wildfire-related PM_2.5_ concentrations and their annual contribution to total PM_2.5_; and (3) quantify county-level long-term mortality burdens attributable to both total and wildfire PM_2.5_.

## 2 Methods

### 2.1 Data collection

Data were collected across the CONUS between 2018 and 2024 from multiple sources. Hourly ground-level PM_2.5_ mass concentrations were obtained from the Air Quality System (AQS) operated by United States Environmental Protection Agency (U.S. EPA), with a total of 1013 regulatory monitoring stations across the CONUS. ^30^ To improve our ML model performance, a series of relevant and available features were selected, including aerosol optical depth (AOD), atmospheric precursor concentrations, meteorological variables, population, economy level, land-use variables, and wildfire indicator. Details can be found in the **Supporting Information (SI) Table S1**.

AOD has been successfully used to predict ambient PM_2.5_, and is commonly used in ML-based PM_2.5_ studies. Satellite AOD at 550 nm was retrieved from the Multi-angle Implementation of Atmospheric Correction (MAIAC) AOD product (MCD19A2), which combined Terra and Aqua Moderate Resolution Imaging Spectroradiometer (MODIS) products and had a 1-km resolution. ^31^ To address missing values of the satellite AOD product (e.g., cloud coverage), additional gap-free modeled hourly AOD product was used from Goddard Earth Observing System composition forecast (GEOS-CF) in 0.25°X0.25° resolution (method for imputation of missing 1-km satellite AOD described in Section 2.2). ^32^

In addition, simulated ground-level PM_2.5_ concentrations were extracted from the GEOS-CF model (0.25°X0.25°, hourly). ^32^ The GEOS-CF product also provided the simulated concentrations of multiple other atmospheric PM_2.5_ precursors, including ammonia (NH_3_), nitrogen oxide (NO), nitrogen dioxide (NO_2_), sulfur dioxide (SO_2_), formaldehyde, benzene, toluene, xylenes, and isoprene. ^32^ We also included anthropogenic emission of monoterpenes and non-methane volatile organic compounds (VOCs) from the Copernicus Atmosphere Monitoring Service (CAMS) global dataset (0.1°X0.1°, monthly) to capture the possible impact of other precursors. ^33^

Meteorological conditions and their vertical profiles are also vital in predicting ambient PM_2.5_ concentrations with high spatiotemporal resolution. Therefore, we included temperature, dewpoint temperature, precipitation, solar radiation, northward and eastward components of wind speed, evaporation, surface pressure, and boundary layer height (BLH) from ERA5-Land (0.1°X0.1°, hourly) and ERA5 (0.25°X0.25°, hourly) reanalysis datasets. ^34, 35^ Since the CONUS experienced severe wildfire events during the study period (2018-2024), we obtained wildfire smoke density (polygon, daily) from NOAA’s Hazard Mapping System (HMS) smoke product. ^36^ Four levels of smoke density are reported in the HMS dataset, including none, light, medium, and heavy smoke.

As indicators of anthropometric activity, emissions, and terrain effects, we included additional datasets. We collected WorldPop population (1 km, annual) and Visible Infrared Imaging Radiometer Suite (VIIRS) nighttime lights (500m, monthly). ^37, 38^ We also considered several land-use variables as proxies of local emissions, including normalized difference vegetation index (NDVI) from MOD13A3 (1 km, monthly), ^39^ percentage of 16 land cover types from National Land Cover Database (NLCD, 30 m, annual), ^40^ road density using all roads from Topologically Integrated Geographic Encoding and Referencing (TIGER) database (annual). ^41^ In addition, elevation data were obtained from Shuttle Radar Topography Mission (SRTM, 90 m). ^42^

All hourly data were aggregated to daily average using UTC time zone for consistency across the CONUS. And all raster data were projected onto a predefined 1-km resolution grid covering the CONUS using the NAD83 / CONUS Albers Equal Area Conic projection coordinated system (EPSG:5070).

### 2.2 Model development of total PM_2.5_

As other studies have previously described, total PM_2.5_ prediction was developed using a four-dimensional space-time extremely randomized trees (4D-STET) model. ^20, 43^ Extremely randomized trees (extra-trees) is a tree-based ensemble learning algorithm, which introduces additional randomness in split selection, thereby improving robustness to noisy data and highly correlated predictors compared with the random forest model. The “four-dimensional space-time” framework incorporates the Euclidean spherical spatial coordinates and triangular spiral temporal representations to account for spatiotemporal heterogeneity. To address the missingness of satellite AOD data, we adopted a two-stage modeling strategy. ^20^ The first stage fit the 4D-STET model to fill the AOD gaps using simulated AOD from GEOS-CF, selected meteorological data, NDVI, elevation, and spatiotemporal terms. Then the second stage applied the 4D-STET model with the gap-filled AOD, all meteorological data, population, nighttime lights, all land-use variables, and wildfire smoke density to predict ground-level daily average total PM_2.5_ concentrations. The hyperparameters in the two 4D-STET models were carefully tuned based on minimizing the out-of-bag (OOB) errors. Details about model structures and hyperparameter tuning are available in **SI Section S1**. With the optimal hyperparameters, the final 4D-STET model was used to predict the daily average total PM_2.5_ concentrations at each grid across the CONUS and in each day between 2018 and 2024.

The final 4D-STET model of ground-level total PM_2.5_ was evaluated by 10-fold cross-validation (CV) approaches. Three CV strategies were applied: Sample-based CV, station-based CV, and day-based CV. ^20^ Specifically, all data samples, regulatory monitoring stations, or days were randomly divided into ten folds separately, with nine folds used for training and the remaining one for validation. This procedure was repeated ten times so that each fold served as the validation dataset once. Although sample-based CV was most commonly used in previous ML-based studies, the assumption of independent samples in sample-based CV was likely violated for spatiotemporally correlated PM_2.5_ data in this study, which potentially resulted in data leakage and overly optimistic estimates of model performance. In contrast, station-based and day-based CV can account for spatial and temporal autocorrelation within the dataset and evaluate model generalizability from both spatial and temporal perspectives. Spatial generalization was particularly important because the regulatory monitoring stations were not evenly distributed across CONUS. We pooled the observations and predictions from validation datasets in ten runs, and then calculated a series of out-of-sample evaluation metrics, including the coefficient of determination (R^2^), root mean squared error (RMSE), mean absolute error (MAE), mean bias error (MBE), and normalized RMSE (NRMSE, defined as RMSE divided by the mean of all observations). To evaluate monthly and annual averages of daily total PM_2.5_ prediction, we also estimated the out-of-sample performance metrics of monthly and annual average predictions based on the above pooled daily CV results, using grid-months and grid-years with at least 80% of observations, respectively.

In addition to evaluating the model predictive performance, we examined model interpretability using the mean absolute SHapley Additive exPlanations (SHAP) values to quantify the relative contributions of individual features to PM_2.5_ predictions. ^44, 45^ Partial dependence plots (PDPs) were further generated for selected features ranking among the highest according to their mean absolute SHAP values, in order to characterize the marginal relationships between total PM_2.5_ levels and individual features. ^46^

### 2.3 Estimation of wildfire PM_2.5_

Based on the daily 1-km total PM_2.5_ predictions obtained in Section 2.2, wildfire-related PM_2.5_ concentrations could be further estimated following previous studies’ approach ^7, 28^. The PM_2.5_ anomalies were first defined as

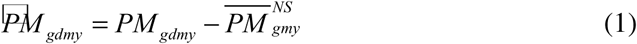

where *PM_gdmy_*is daily average total PM_2.5_ concentrations in grid *g*, day *d*, month *m*, and year *y*; and 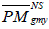 is the recent 3-year location– and month-specific median total PM_2.5_ concentrations on non-smoke days, which was

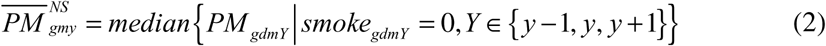

where *smoke_gdmy_* is a binary variable indicating whether grid *g* was covered by wildfire smoke in day *d*, month *m*, and year *y*, based on NOAA’s HMS product. Then, daily 1-km wildfire-related PM_2.5_ was obtained by using PM_2.5_ anomalies only for those grid-days with wildfire smoke overhead, shown in Equation (3) below ^7, 28^.

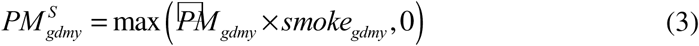

After getting the daily 1-km estimates of wildfire PM_2.5_, we further aggregated them into monthly and annual average estimates. The percentage of wildfire PM_2.5_ in total PM_2.5_ was also calculated for each annual average 1-km estimate to quantify the relative contribution of wildfire smoke in different areas across the CONUS.

To evaluate the reliability and spatial generalizability of our wildfire PM_2.5_ estimates, we used the out-of-sample total PM_2.5_ predictions generated through station-based spatial CV. At each held-out AQS station, wildfire PM_2.5_ was derived from the spatial-CV prediction and compared with an AQS-based wildfire PM_2.5_ estimate derived from the observed total PM_2.5_ concentration using the same wildfire-subtraction approach described in Equations (1)-(3). Agreement was assessed using R^2^, RMSE, MAE, MBE, and NMAE for wildfire PM_2.5_ concentrations at daily and annual scales, as well as for wildfire contributions to annual average total PM_2.5_.

### 2.4 Mortality burden attributable to PM_2.5_

Since long-term PM_2.5_ exposure can substantially contribute to burden of disease, this study estimated the mortality burden attributable to total and wildfire PM_2.5_ at the county level across the CONUS for each year between 2018 and 2024. The distributions of both population and PM_2.5_ concentrations within each county were taken into consideration for exposure assessment. We used an exposure-response relationship between total PM_2.5_ and all-cause mortality based on a nationwide cohort of U.S. adults, where the relative risk (RR) was 1.12 (95% confidence interval (CI): 1.08-1.15) per 10 μg/m^3^ increase in PM_2.5_^47^. Following the methods from Global Burden of Disease (GBD) study, we first calculated the population attributable fraction (PAF) of total PM_2.5_ as below ^5^.

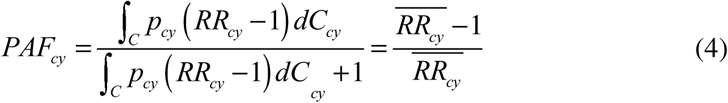

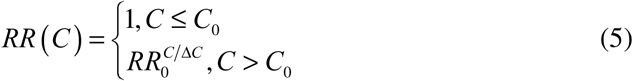

where *PAF_cy_*is PAF at county *c* in year *y*, *p* is the population fraction under *C* exposure, *RR*_0_ is 1.12 and approximately lognormally distributed, *C*_0_ is the theoretical minimum-risk exposure level (TMREL), yielding a uniform distribution between 2.4 and 5.9 μg/m^3^ ^5^, and △*C* is the unit increase in PM_2.5_, i.e., 10 μg/m^3^ here.

Finally, the attributable mortality burden of total PM_2.5_ at county *c* in year *y* can be estimated by

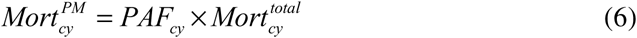

Where 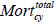 is the baseline all-cause mortality rate per 100,000 at county *c* in year *y*, which was obtained from the CDC WONDER (Wide-ranging Online Data for Epidemiologic Research) system ^48^. The mortality burden attributable to wildfire PM_2.5_ could be further estimated based on a proportional PAF approach used in GBD studies, as shown in Equation (7) below ^5^.

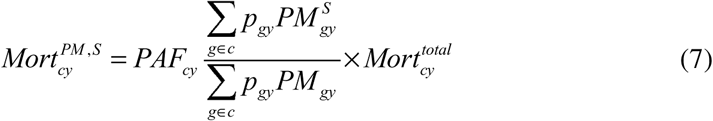

where the superscript *S* refers to wildfire smoke. A two-dimensional Monte Carlo simulation approach was used to estimate the uncertainty in attributable mortality burden ^49–51^. Details about uncertainty analysis can be found in **SI Section S2**.

All spatial analysis and statistical analyses described in the Methods section were performed with R V4.5.2.

## 3 Results

### 3.1 Model performance

**Figure 1** shows that the 4D-STET model accurately estimated the daily average total PM_2.5_ concentrations at most regulatory monitoring stations. Approximately 70% of regulatory monitoring stations had a sample-based CV R^2^ higher than 0.6 and NRMSE lower than 0.4. On average, the sample-based CV R^2^ and RMSE (NRMSE) of this model reached 0.72 and 2.76 μg/m^3^ (0.38) for daily estimates, respectively. The performance increased after aggregating daily estimates into monthly (CV R^2^ = 0.86, NRMSE = 0.16) and annual averages (CV R^2^ = 0.89, NRMSE = 0.10).

**Figure 1.**
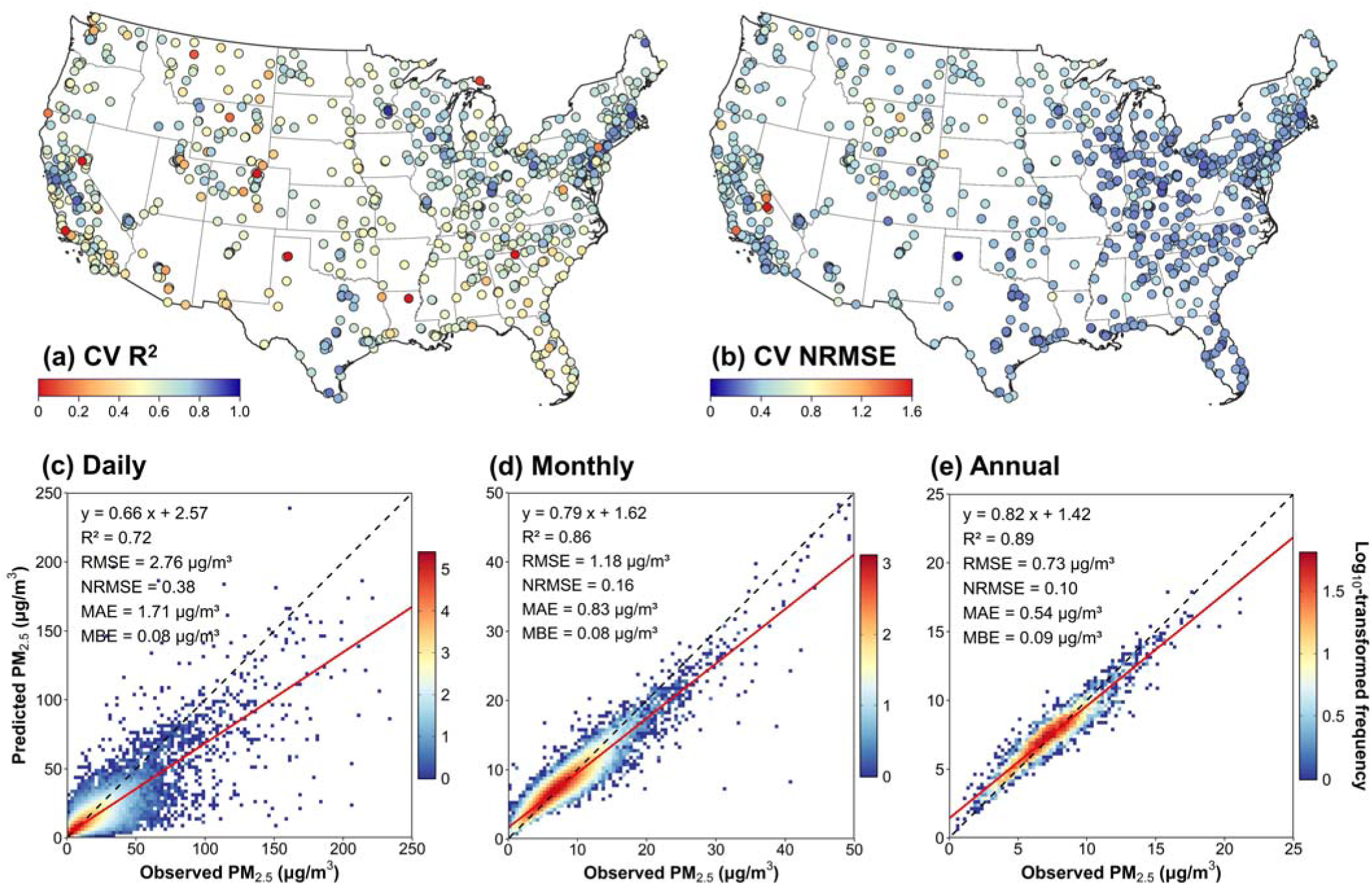
Model performance in the 10-fold sample-based cross-validation (CV): Spatial distribution of sample-based CV (*a*) R^2^ and (*b*) NRMSE of daily average total PM_2.5_ estimates in each regulatory monitoring station across the CONUS; and density scatterplots and related out-of-sample metrics of sample-based CV results for (*c*) daily, (*d*) monthly, and (*e*) annual average predictions.

Because sample-based CV did not account for spatiotemporal dependence within the data, station-based and day-based CV yielded slightly lower, but still acceptable, performance. Station-based CV obtained a CV R^2^ of 0.63 and an RMSE (NRMSE) of 3.22 μg/m^3^ (0.44) (**Figure S1**), while day-based CV had a CV R^2^ of 0.63 and an RMSE (NRMSE) of 3.20 μg/m^3^ (0.44). It suggests that this model had relatively high capability in predicting daily total PM_2.5_ concentrations in those areas and those days without any ground-level monitoring data. The CV performance of the ML model in this study was broadly consistent with and sometimes better than those reported in previous studies (details in Discussion). More detailed out-of-sample metrics in different temporal scales, divisions, and years are available in **Tables S3** and **S4**.

In addition to predictive performance, it is also essential to examine the feature contribution to the model predictions to understand the model behavior. The mean absolute SHAP values for the top 15 features are shown in **Figure S2**. The PM_2.5_ concentrations simulated by GEOS-CF model ranked first among all features and had a mean absolute SHAP value of 1.20 μg/m^3^, which was much higher than other features. Except for the spatial and temporal terms, temperature, formaldehyde concentration, wildfire smoke density, and BLH had moderate SHAP values, ranging from 0.48 to 0.60 μg/m^3^, followed by those with lower SHAP values, such as land cover, dewpoint temperature, wind speed, AOD, ammonia, and remaining features. We also examined the trend between PM_2.5_ prediction and these features through PDPs (**Figure S3**). We observed that predicted PM_2.5_ exhibited a monotonic increasing association with GEOS-CF PM_2.5_, formaldehyde, and ammonia concentrations, AOD, and wildfire smoke density, and a monotonic decreasing association with BLH. In contrast, the associations with temperature and dewpoint temperature followed a U-shaped pattern, suggesting more complicated underlying mechanisms.

**Figure S4** further presents an out-of-sample evaluation of the estimated wildfire PM_2.5_ contributions. During smoke days, predicted daily average wildfire PM_2.5_ concentrations showed moderate agreement with observation-based estimates at AQS sites (R^2^ = 0.65, NMAE = 0.48). The regression slope of 0.59 and an MBE of −0.65 μg/m^3^ indicated that the model tended to underestimate high daily wildfire PM_2.5_ concentrations. Agreement improved substantially after aggregation across all days of each year, for both annual wildfire PM_2.5_ concentrations (R^2^ = 0.83, NMAE = 0.23, MBE = –0.14 μg/m^3^) and annual wildfire contributions to total PM_2.5_ (R^2^ = 0.80, NMAE = 0.24, MBE = –0.02). These results support the reliability of the broad spatial and interannual patterns in the estimated wildfire contribution, while suggesting that daily extremes and exposure contrasts were attenuated, thus probably providing a lower-bound estimate of wildfire contributions in this study.

### 3.2 Spatiotemporal patterns of daily PM_2.5_ concentrations

Our daily average PM_2.5_ mapping can capture peak PM_2.5_ exposure patterns across the CONUS during extremely polluted days, which are often obscured in annual average maps due to temporal averaging and offsetting effects. **Figure 2** presents the 1-km daily average total PM_2.5_ concentration in six highly polluted days, most of which were driven by wildfire events. For instance, a series of record-breaking wildfire events happened along the U.S. west coast in early September 2020. Consequently, as shown in **Figure 2b**, dense wildfire smoke spanned across California, Oregon, and Washington on September 12, 2020, leading to extremely high daily average total PM_2.5_ exposure exceeding 100 μg/m^3^ across large areas of these three states. Another example on June 7, 2023 (**Figure 2e**) illustrated the long-range transport of wildfire smoke from Canada to the northeast U.S., leading to severely elevated daily total PM_2.5_ concentrations in New York, Pennsylvania, New Jersey, Connecticut, and Delaware. Although **Figure 2** shows the total PM_2.5_ concentrations, the above results can be also verified by our daily wildfire PM_2.5_ results.

**Figure 2.**
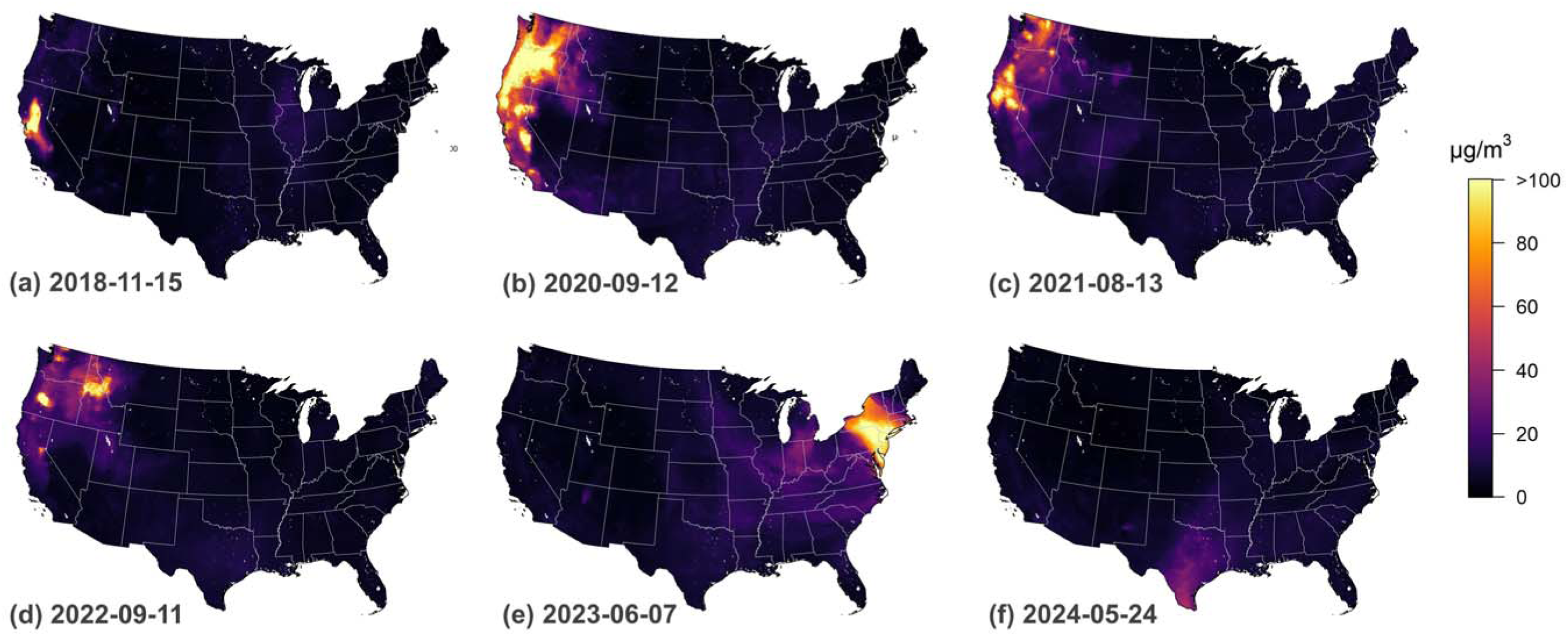
Examples of daily 1-km total PM_2.5_ concentration estimates across the CONUS on several days with relatively high PM_2.5_ peaks.

We also compared the daily averages with relevant daily limits in U.S. National Ambient Air Quality Standard (NAAQS) and World Health Organization (WHO) Air Quality Guidelines, in order to identify high-exposure regions. **Figure 3** shows the proportion of days from 2018 to 2024 with higher daily average total PM_2.5_ concentrations than 35 μg/m^3^ in NAAQS and 15 μg/m^3^ in WHO Air Quality Guidelines, respectively. As illustrated in **Figure 3a**, states along the U.S. west coast, especially north California, experienced extremely polluted conditions (daily PM_2.5_ > 35 μg/m^3^) on more than 2% of days. This pattern is consistent with the spatial distribution of wildfire events across the CONUS. In contrast, **Figure 3b** suggests more hotspots which went through highly polluted days (daily PM_2.5_ > 15 μg/m^3^). Besides the U.S. west coast, many metropolitan areas had relatively highly polluted conditions on more than 5% of days, such as Los Angeles, New York City, Chicago, Salt Lake City, Phoenix, Denver, and Houston, suggesting the impact of anthropogenic emissions on total PM_2.5_ levels. The south Texas and central valley of California also experienced considerable polluted days.

**Figure 3.**
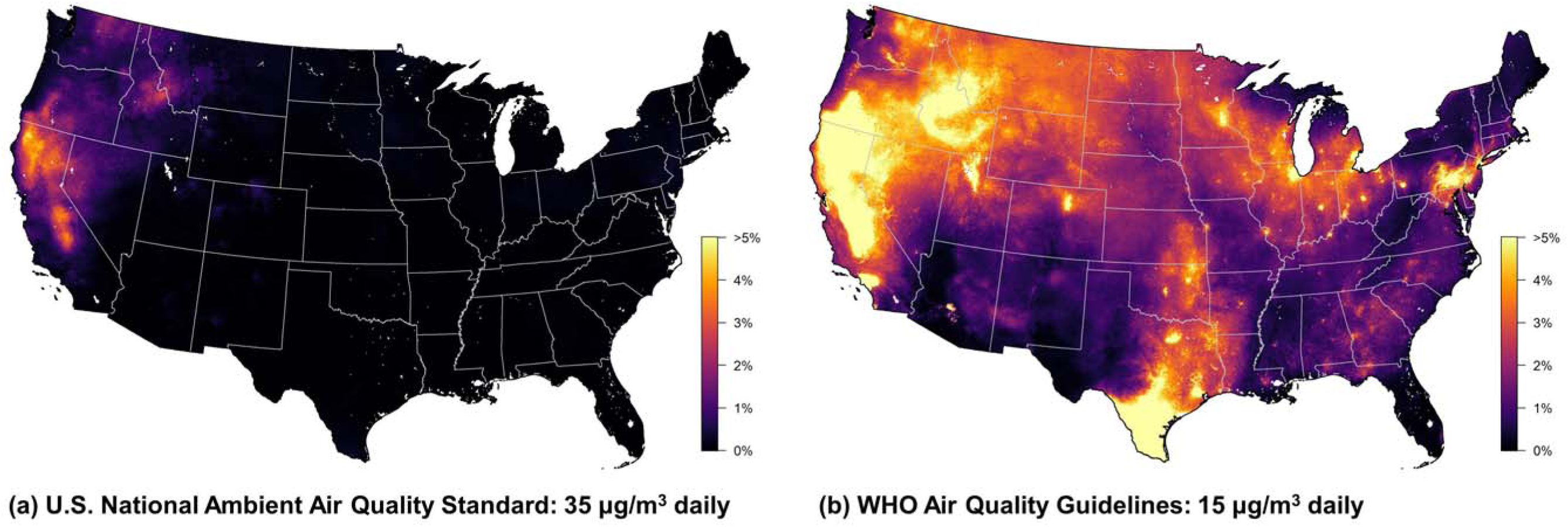
Percentage of days, between 2018 and 2024, with daily average total PM_2.5_ concentrations above daily limit in (*a*) U.S. National Ambient Air Quality Standard (NAAQS) and (*b*) WHO Air Quality Guidelines.

### 3.3 Spatiotemporal patterns of annual PM_2.5_ concentrations

Daily average PM_2.5_ concentrations were further aggregated into the annual average level for long-term exposure assessment. As shown in **Figure 4a**, higher annual average total PM_2.5_ concentrations were observed in both U.S. west coast and eastern U.S. Specifically, the multi-year average of annual average total PM_2.5_ reached 6.82 μg/m^3^ in Pacific Division and 7.72, 7.25, and 7.05 μg/m^3^ in West South Central, East South Central, and East North Central Divisions, respectively, which were much higher than 4.80 μg/m^3^ in Mountain Division and 4.69 μg/m^3^ in New England Division (**Table S5**; Divisions are defined in **Figure S5**).

**Figure 4.**
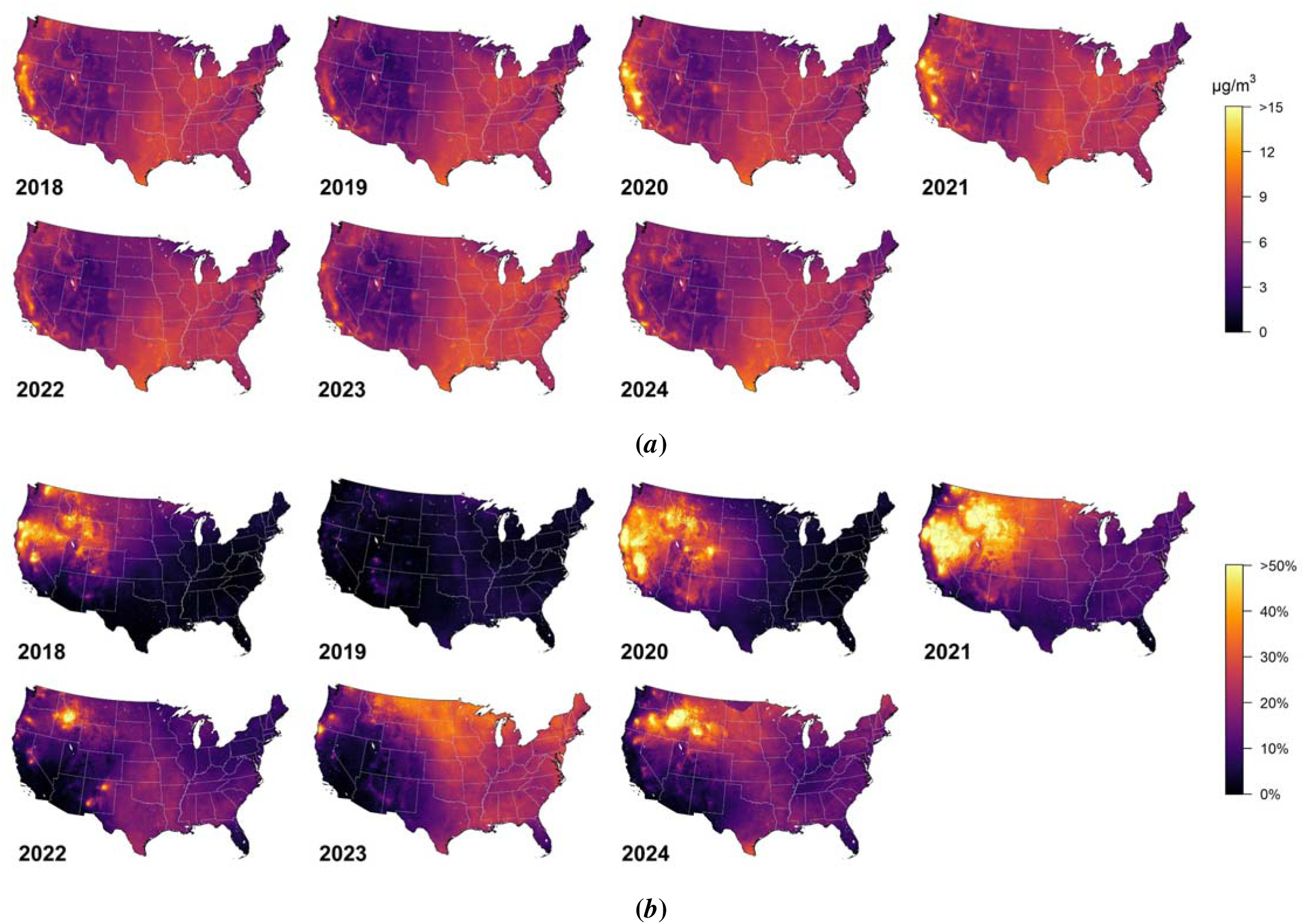
Annual average 1-km estimate of (*a*) total PM_2.5_ and (*b*) proportion of wildfire PM_2.5_ in total PM_2.5_ between 2018 and 2024.

However, high concentrations in western and eastern U.S. exhibit different temporal patterns. The relatively high total PM_2.5_ concentration in eastern U.S. kept stable between 2018 and 2024 with the standard deviation (SD) ranging from 0.22 to 0.47 μg/m^3^ among different divisions. In contrast, the annual average total PM_2.5_ concentrations along the west coast had a dramatically higher variance (SD = 0.91 μg/m^3^ in Pacific Division) across different years, showing much higher concentrations in 2018 (7.36 μg/m^3^), 2020 (8.27 μg/m^3^), and 2021 (7.83 μg/m^3^) than the remaining four years (5.69-6.27 μg/m^3^). The only exceptions along the west coast were the Central Valley of California and Los Angeles metropolitan area, which always had relatively high annual total PM_2.5_ concentrations between 2018 and 2024.

Wildfire smoke had non-negligible contributions to annual average total PM_2.5_ concentrations, as shown in **Figure 4b**. Detailed proportions and their mean and standard deviation by divisions are further provided in **Table S5**. In the Pacific Division along the west coast, wildfire smoke contributed 23.6%, 34.5%, and 29.5% of annual average total PM_2.5_ concentrations in 2018, 2020, and 2021, respectively. In some regions of western U.S., wildfire smoke accounted for over 50% in these three years, especially in 2021 with the largest wildfire impacted area. Despite not being the major driver of PM_2.5_ concentration elevation, wildfire events still accounted for over 10% of total PM_2.5_ in most of eastern U.S. between 2021 and 2024. In particular, the wildfire contribution increased to over 25% in the Midwest and Northeast U.S. in 2023 when these regions were strongly influenced by Canadian wildfire smoke.

### 3.4 PM_2.5_-related mortality burden

Between 2018 and 2024, the number of deaths attributable to total PM_2.5_ across the CONUS ranged from 110,000 (95% uncertainty interval (UI): 79,500 – 138,000) in 2019 to 152,000 (110,000 – 191,000) in 2021 (**Table S7**). Unlike the decline between 2000 and 2020, the PM_2.5_-attributable mortality did not exhibit a monotonic trend between 2018 and 2024. **Figure 5a** further shows the county-level mortality rate attributable to total PM_2.5_ across the CONUS (details in **Table S8**). The southeastern U.S. consistently experienced the highest mortality burden from ambient PM_2.5_ exposure during 2018-2024, whereas the U.S. west coast showed relatively high mortality burden only in 2020 and 2021, which were severely wildfire-influenced years. Taking 2021 as an example, the East South Central, West South Central, and Pacific Divisions had attributable mortality rates of 82.3 (59.7 – 103.7), 72.8 (52.9 – 91.6), and 63.4 (46.2 – 79.7) per 100,000, respectively. In contrast, the lowest attributable mortality rate was observed in the New England Division in northeastern U.S., which reached only 35.2 (25.5 – 44.5) per 100,000 (**Table S7**).

**Figure 5.**
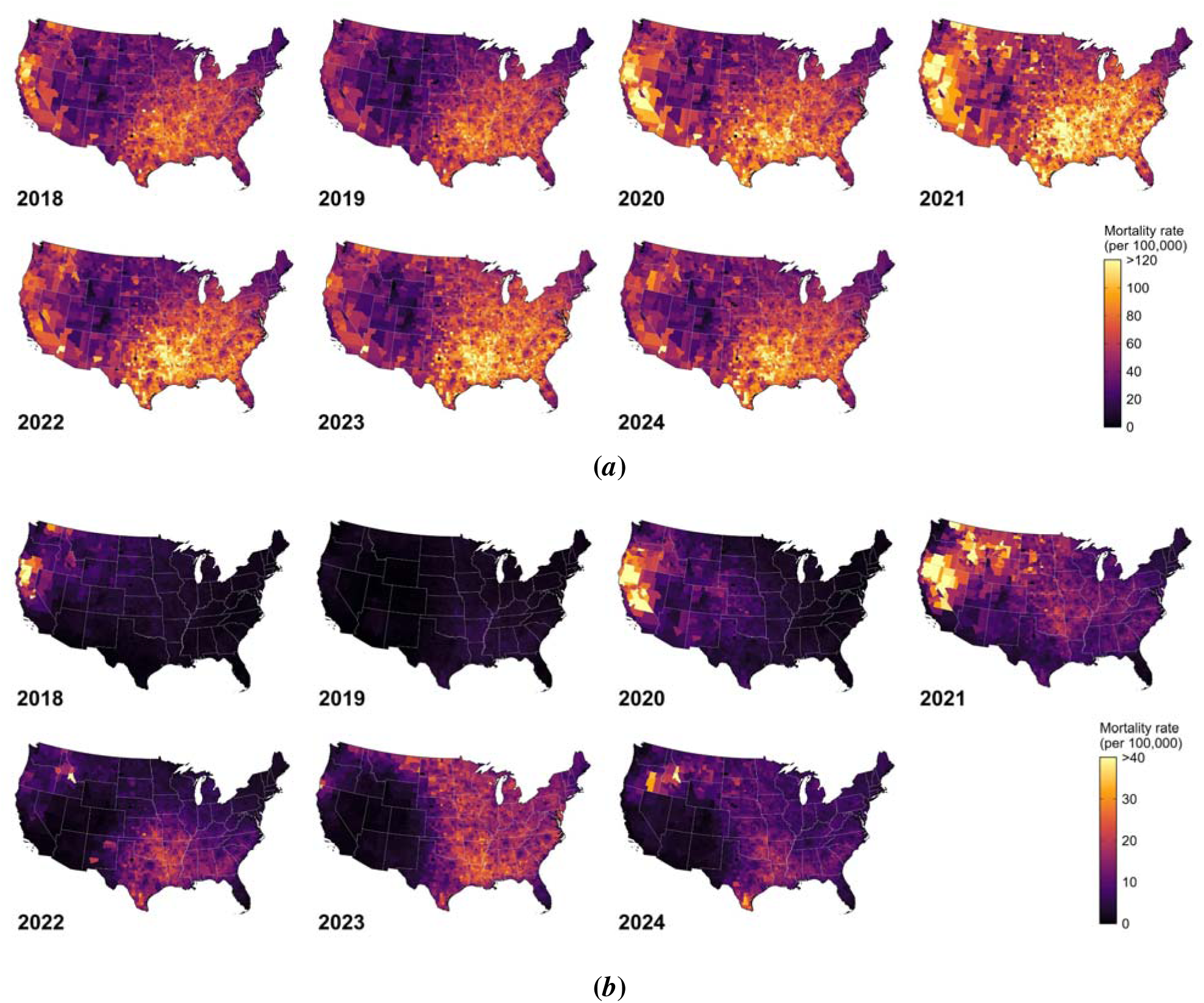
Annual mortality burden (per 100,000) attributable to (*a*) total PM_2.5_ and (*b*) wildfire PM_2.5_ between 2018 and 2024.

For wildfire-related PM_2.5_, the attributable number of deaths across the CONUS ranged from 2,870 (2,080 – 3,610) in 2019 to 25,100 (18,200 – 31,600) in 2023 (**Table S7**). **Figure 5b** provides the county-level mortality rate due to wildfire PM_2.5_ exposure (details in **Table S8**). It can be observed that the wildfire-related mortality burden was higher along the U.S. west coast in 2018, 2020, and 2021, whereas higher burdens were found in the eastern U.S. in 2022 and 2023. Compared with 2019 which experienced the lowest wildfire impact, the wildfire-related mortality burden increased nearly 30-fold in the Pacific Division in 2020 (0.48 to 13.83 per 100,000, accounting for 1.1% to 20.4%) and about 6-fold in the East South Central Division in 2023 (2.59 to 15.48 per 100,000, accounting for 4.1% to 20.6%). In contrast, the non-wildfire PM_2.5_ corresponded to relatively stable attributable deaths across the study period (**Table S7**), except slightly increase in 2020 and 2021 due to the COVID-19 pandemic. Therefore, the variations in attributable deaths of total PM_2.5_ during the study period were dominated by wildfire smoke events.

## 4 Discussion

This study provides high-resolution estimates of total and wildfire-related PM_2.5_ concentrations across the CONUS during recent wildfire-influenced years and highlights the growing role of wildfire smoke in shaping PM_2.5_ exposure patterns. Our results suggest that while the U.S. west coast experienced the most severe episodic pollution events, elevated total PM_2.5_ concentrations were also observed in several metropolitan areas across both western and eastern regions. The contribution of wildfire smoke to total PM_2.5_ was over 30% during major wildfire years in the west coast and over 20% during long-range smoke transport events in the east coast, highlighting the increasing influence of climate-driven wildfire activity on air quality. Consistent with these exposure patterns, the mortality burden attributable to wildfire-related PM_2.5_ also showed considerable interannual variability, underscoring the growing public health importance of wildfire smoke in the U.S.

### 4.1 Comparison of ML performance with previous studies

The CV performance of the ML model in this study was broadly consistent with, and in some cases slightly better than, those reported in previous studies focusing on the prediction of daily average total PM_2.5_ (**Table S3**). Since exposure and health risk assessments primarily depend on the accuracy of absolute exposure estimates, RMSE was considered as the primary CV metric. However, as different studies covered different time periods with quite different total PM_2.5_ concentrations, NRMSE provides a more appropriate basis for comparison.

The sample-based CV NRMSE at the daily scale in this study was 0.38 across the CONUS, which was similar to values reported by Wei et al. (0.40; CONUS, 2000-2020) ^15^ and Kayastha et al. (0.37; CONUS, 2018-2022). ^11^ The sample-based CV NRMSEs in the Mountain and Pacific Divisions reached 0.45 and 0.47, respectively, which were lower than that reported by Reid et al. (0.67; western U.S., 2008-2018). ^12^ Similar patterns were observed for station-based CV. Notably, the comparable or improved performance in this study was achieved under more challenging modeling conditions. For example, Wei et al. and Reid et al. covered the early 2000s with higher total PM_2.5_ concentrations which may be easier to predict due to stronger signal-to-noise ratios; ^12, 15^ and Kayastha et al. developed predictions at a coarser 4-km spatial resolution instead of 1-km resolution. ^11^

Many previous studies did not report NRMSE values, but instead reported CV R^2^, which we considered as our secondary evaluation metric. The sample-based CV R^2^ in this study (0.72) falls within the range reported in previous studies in U.S. and North America (0.62-0.83). ^9, 11, 12, 14–16, 18^ Similarly, the station-based CV R^2^ in this study (0.63) is comparable to previously reported values (0.59-0.86). ^8, 9, 11–13, 15, 16^ The highest station-based CV R^2^ (0.86) was reported in Di et al. ^8^ However, many of these studies were conducted under substantially different pollution regimes. Di et al. developed daily PM_2.5_ estimates from 2000 to 2015, when anthropogenic emissions dominated the ambient PM_2.5_ levels across the CONUS. ^8^ In contrast, the 2018-2024 period in this study was characterized by generally low background PM_2.5_ concentrations on non-wildfire days but episodic extreme concentrations on wildfire days. Because R^2^ depends on the total variance of the observations, it can be lower in settings with limited background variability but occasional extreme events, even when absolute prediction errors remain small. Therefore, NRMSE may provide more informative measures of model performance for exposure assessment in recent wildfire-influenced periods.

In addition, a limited number of previous studies reported CV performance on aggregated monthly and annual average concentrations. The station-based CV NRMSE of annual averages was 0.19 in this study, lower than 0.26 in Wei et al. ^15^ and 0.25 in Shen et al. ^17^

### 4.2 Interpretation of important features in the ML model

Section 3.1 identified the most important features in the 4D-STET model and further examined their trends with ground-level total PM_2.5_ predictions using PDPs (**Figure S3**), including GEOS-CF simulated PM_2.5_, formaldehyde, ammonia, as well as AOD, temperature, dewpoint temperature, wildfire smoke density, BLH, and northward wind speed. Some trends are easily understandable, such as the positive associations with GEOS-CF simulated PM_2.5_, AOD, and wildfire smoke density, whereas other trends require more explanations.

GEOS-CF simulated PM_2.5_ was the most influential feature based on its mean absolute SHAP value and exhibited a positive marginal association with predicted ground-level total PM_2.5_ (**Figure S3 (*a*)**). As a global chemical transport modeling system, GEOS-CF provides spatially and temporally complete, process-based estimates that integrate information on emissions, atmospheric chemistry, transport, deposition, and meteorological conditions. It therefore served as a baseline estimate of regional and temporal PM_2.5_ variation for the ML model. A conceptually similar modeling strategy has been used in previous studies, in which spatially interpolated regulatory monitoring observations provided an observation-based baseline estimate for subsequent ML prediction. ^52^ Compared with interpolation based primarily on the geographic configuration and AQS monitoring data, the GEOS-CF field can represent pollution transport and chemical processes across areas with sparse monitoring coverage.

Positive marginal associations were observed between total PM_2.5_ predictions and GEOS-CF simulated ammonia and formaldehyde from PDPs in **Figure S3 (*i*)** and **(*c*)**, respectively. This can be interpreted by secondary aerosol formation mechanisms. Ammonia is one of the most important precursors to PM_2.5_ and plays a vital role in the formation of secondary inorganic aerosols (SIAs), such as ammonium nitrate (NH_4_NO_3_) and ammonium sulfate ((NH_4_)_2_SO_4_). ^53–55^ Several studies also suggest that reducing ammonia emissions can efficiently reduce ambient PM_2.5_ pollution. ^56, 57^ One source apportionment study across the CONUS shows that secondary nitrate and secondary sulfate contributed 30.8%-74.6% of total PM_2.5_ mass across different regions in the CONUS, which are the dominant components of PM_2.5_. ^58^ This can possibly explain why ammonia was identified as an important feature in our model. In contrast, although formaldehyde is not a direct precursor of PM_2.5_, it is typically used as a proxy of volatile organic compound (VOC) reactivity as formaldehyde is the oxidation product of many anthropogenic and biogenic VOCs. ^59–62^ As VOCs play critical roles in the formation of secondary organic aerosols (SOAs), formaldehyde may function as an indicator of SOAs. ^59–62^ The source apportionment results also show that SOAs accounted for 1.2%-18.2% across the CONUS. ^58^ That is possibly why formaldehyde was selected as one of the most important features in this study.

Besides the above precursors or proxy compounds, some essential meteorological features were also identified. **Figure S3 (*e*)** shows a decreasing trend between BLH and total PM_2.5_ predictions. The BLH can depress the diffusion of ground-level PM_2.5_ to the upper atmosphere, so PM_2.5_ usually accumulates under the BLH and a lower BLH will then exacerbate the ground-level PM_2.5_ pollution. ^63, 64^ Many field papers have observed their negative associations. ^63–65^ The associations with temperature and dewpoint temperature are more complicated. **Figure S3 (*b*)** exhibits a U-shaped trend between total PM_2.5_ predictions and temperature. Higher temperatures over 20LJ are associated with higher PM_2.5_ concentrations, probably because higher temperatures can promote atmospheric photochemical reactions to form SOAs ^54, 66, 67^ and higher temperatures are often correlated with more wildfire events and more biogenic VOC emissions. ^68^ By contrast, lower temperatures below 5LJ are also associated with higher PM_2.5_ concentrations. This is because wintertime lower temperatures are typically accompanied by a shallow boundary layer and frequent near-surface temperature inversions, which reduce atmospheric mixing and trap PM_2.5_ near the ground surface. ^65, 69–71^ Relatively low temperatures may be also correlated with more residential heating emissions, such as oil and wood combustion. Besides, low temperature favors ammonium nitrate formation, which is a major component of PM_2.5_. ^72, 73^ The above complicated underlying mechanisms shape the nonlinear patterns of the trend with temperature. A similar U-shaped pattern was found for dewpoint temperature in **Figure S3 (*f*)**. Lower dewpoint temperatures, i.e., low humidity, are associated with higher total PM_2.5_ predictions, probably because low humidity corresponds to weaker wet removal of PM_2.5_, ^74, 75^ and is also typically related to wintertime with shallow BLH and stagnant air. As the dewpoint temperature increases over 10LJ, the PM_2.5_ predictions also slightly increase. One of the possible underlying mechanisms is the hygroscopic growth of particles, whereby high relative humidity promotes water uptake by aerosols, increasing particle size and mass concentrations. ^64, 76, 77^ In addition, higher humidity may promote aqueous-phase secondary aerosol formation as well as the gas-to-particle partitioning, especially ammonium nitrate. ^64, 78, 79^

### 4.3 Interpretation of spatiotemporal variations in PM_2.5_

The spatiotemporal variations in total PM_2.5_ can be influenced by multiple factors, including primary emission sources, secondary aerosol formation, and topography effects. The U.S. west coast shows a relatively higher annual average total PM_2.5_ concentrations than other CONUS regions, especially in north California, central valley of California, and Los Angeles Basin. Specifically, north California, as well as south Oregon and east Washington, show substantially higher total PM_2.5_ concentrations in severe wildfire years, such as 2018, 2020, and 2021. Additionally, in these areas, **Figure S6** supports that wildfire-related PM_2.5_ was higher than other regions, and **Figure S7** demonstrates that average total PM_2.5_ was higher in summer and fall than the other two cold seasons. Since these three areas do not have very populated metropolitan areas, anthropogenic emissions cannot be the major driver of the elevated PM_2.5_ level. All the above evidence validates that the high total PM_2.5_ concentrations in these areas are dominated by severe wildfire smoke. In contrast, as the second largest metropolitan area in the U.S., the poor air quality in the Los Angeles Basin is dominantly impacted by anthropogenic emission sources and topography effects. Previous source apportionment analysis reveals that fresh traffic emissions, residual oil combustion from ships and oil refineries, and biomass burning possibly due to residential heating accounted for 13.4%, 13.5%, and 4.1% of total PM_2.5_ mass, respectively. ^80^ The strong solar radiation and warm temperature in the Los Angeles Basin can also promote photochemical secondary aerosol formation. Besides, due to proximity to the Pacific Ocean, sea salt contributed 20.6% of total PM_2.5_, which is a unique but very important source and may act as a critical role for secondary aerosols near the sea-land interface. ^80, 81^ Last but not least, the Los Angeles Basin is surrounded by several mountains and often suffers from marine inversions, so the air pollutants are typically trapped in the cooler air near the ground surface. ^80^ All the abovementioned factors exacerbate the PM_2.5_ pollution in the Los Angeles Basin. The central valley of California combines the adverse characteristics of both north California and Los Angeles Basin, which makes it one of the worst PM_2.5_ polluted region across the CONUS. The central valley is impacted by both severe wildfire smoke in summertime and anthropogenic emissions from traffic and agricultural burning sources. ^82^ Additionally, the topographical features of this valley as well as the strong temperature inversion and shallow BLH in wintertime further aggravate the PM_2.5_ pollution in this area (**Figure S7 (*d*)**). ^82^

In addition to the U.S. west coast, vast areas in eastern U.S. also show moderately elevated total PM_2.5_ levels, where some hotspots can be observed around those metropolitan areas. Overall, SIA formation, including secondary sulfate and nitrate, had a higher contribution to PM_2.5_ in eastern U.S. (48.6%-74.6%) than western U.S. (30.8%-44.2%) and came from slightly different sources. The Ohio River Valley showed relatively high total PM_2.5_ concentrations, especially in summer and winter (**Figure S7**). In summer, secondary sulfate dominated the PM_2.5_ mass concentrations, which was likely contributed from industrial emissions, such as vehicle manufacturing, steel production, petroleum refining, and coal power plants. ^58, 83^ More sunlight, higher temperature, and higher humidity in summer can promote photochemical reactions for secondary sulfate formation. By contrast, in wintertime, secondary nitrate became the major contributor of PM_2.5_ concentrations, which possibly originated from vehicle exhaust, agricultural emissions, and power plants, ^58, 83^ because lower temperature favors ammonium nitrate formation and lower BLH and more stagnant air accumulates these particles near the ground surface. ^72, 73^ Compared with the Ohio River Valley, the South Division, especially Texas, demonstrated a quite different seasonal pattern, in which total PM_2.5_ concentrations were higher in spring and summer. One previous study near U.S.-Mexico border in Texas reveals that biomass burning and crustal dust accounted for 12.8% and 12.7% of PM_2.5_ mass, respectively. ^84^ The agricultural burning and wildfire smoke, as well as dust were transported from Mexico and southwest U.S. to Texas during spring and summer. Besides, SOA concentrations were much higher in South Division than other areas in eastern U.S., because there were higher VOC emissions in these areas from industry, transportation, and biogenic sources. ^58, 85^ The photochemical reactions with these VOCs are the major pathways to form SOAs in summer. Different from Ohio River Valley and south U.S., the total PM_2.5_ hotspot between Washington D.C. and New York City was mostly contributed by urban anthropogenic emissions, including road traffic, biomass burning (e.g. wood combustion for heating), and residual oil (e.g. ships and industries). ^86, 87^

### 4.4 Implications for population exposure

The high-resolution mapping of PM_2.5_ concentrations is critical for reducing possible exposure misclassification. Many EPA regulatory monitoring stations are located far away from the local sources, so using the nearest EPA regulatory monitoring data or their spatial interpolations, such as inverse distance weighting and kriging, may not adequately capture the impact of emission sources on PM_2.5_ concentrations in those areas without regulatory monitoring. Machine learning models could address this limitation by incorporating many source-related predictors, such as road density and emission inventories. Some widely used PM_2.5_ products are available only at relatively coarse spatial resolutions (e.g., 10 km and 12 km). ^88, 89^ To evaluate the impact of spatial resolution, we conducted a sensitivity analysis by aggregating our 1-km estimates into a 10-km resolution. **Figure S8** compares annual average total PM_2.5_ concentrations derived from 1-km and 10-km products in eight metropolitan areas across the CONUS in 2021. The 10-km total PM_2.5_ estimates were generally lower than our 1-km total PM_2.5_ estimates, particularly those in core urban areas. This difference likely reflects the loss of fine-scale spatial variability, because the substantial heterogeneity in total PM_2.5_ within the core urban areas is smoothed by the coarser-resolution estimates. Therefore, the finer spatial-resolution estimates can offer more valuable insights for prioritizing regions for emission control and exposure mitigation.

In addition to high spatial resolution, this study also predicted PM_2.5_ at a high temporal resolution, i.e., daily averages, which can further reduce exposure misclassification associated with temporal averaging. Although annual average estimates are very useful for long-term health risk assessment, annual averages may obscure substantial day-to-day variations in PM_2.5_ concentrations, particularly in those regions influenced by mask short-term episodic pollution such as wildfire smoke. In contrast, daily exposure estimates can better characterize exposure dynamics, capture peak exposure metrics in addition to long-term cumulative exposure, and facilitate future epidemiological studies about short-term health effects.

Furthermore, the daily estimates can enable the separation of wildfire and non-wildfire PM_2.5_ contributions. Although many PM_2.5_ models have been developed in previous studies, relatively few distinguish between wildfire and non-wildfire contributions. This separation is essential because the two sources typically exhibitdistinct temporal patterns and potentially different exposure-response relationships, thus possibly contributing disproportionately to short-term health risks. Previous studies have suggested that short-term exposure to wildfire-related PM_2.5_ may have much stronger respiratory health effects than non-wildfire PM_2.5_ exposure. ^90, 91^ Future epidemiological studies can also rely on this separation to examine the associations between wildfire PM_2.5_ and more health outcomes. Therefore, the finer temporal-resolution estimates can further improve the precision in exposure and health risk assessment.

Understanding population exposure to PM_2.5_ differs from simply mapping PM_2.5_ concentrations, since highly polluted areas may have relatively low population densities. **Table S6** show the population-weighted annual average total PM_2.5_ exposure in nine divisions, which can be compared with unweighted annual average total concentrations in **Table S5**. Taking Pacific Division as an example, the population-weighted average exposure of total PM_2.5_ (9.46 μg/m^3^) was higher than the average concentration (6.82 μg/m^3^), whereas the population-weighted average exposure of wildfire-related PM_2.5_ (0.75 μg/m^3^) was lower than the average concentration (1.31 μg/m^3^). The higher value of total PM_2.5_ exposure was possibly because a large fraction of population resides in the metropolitan areas, such as Los Angeles and the Bay Area, where total PM_2.5_ mass is dominated by urban anthropogenic emissions. The opposite finding for wildfire-related PM_2.5_ is also plausible, since wildfires often occur in mountainous and forested regions, where smoke concentration can be relatively high, but population density is low. Taking Middle Atlantic Division as another example, the finding about total PM_2.5_ was similar to that observed in the Pacific Division. The population-weighted average exposure of total PM_2.5_ (7.52 μg/m^3^) was higher than the average concentration (6.22 μg/m^3^). Nevertheless, the population-weighted average exposure of wildfire-related PM_2.5_ (0.71 μg/m^3^) was slightly higher than the average concentration (0.64 μg/m^3^). This pattern is likely attributable to the long-range transport of wildfire smoke from Canada, which often affects densely populated urban corridors in this region. Hence, the above findings highlight the importance of considering population distribution when interpreting PM_2.5_ concentration patterns in exposure assessment.

### 4.5 Implications for public health

Unlike the declining temporal trend observed in the first two decades of the 21st Century across the CONUS, ^15^ the mortality burden attributable to total PM_2.5_ did not show a clear decline between 2018 and 2024. One important reason is that recent increases in wildfire smoke offset the improvements in anthropogenic PM_2.5_ reduction. As shown in **Figure S9**, non-wildfire PM_2.5_ exposure driven by anthropogenic emissions continued to show a gradual decline in most divisions from 2018 to 2024. However, wildfire-related PM_2.5_ exposure was the primary driver of the year-to-year variation in total PM_2.5_ exposure. Besides, population aging and increase in baseline mortality due to COVID-19 pandemic may also have contributed to the stagnation in attributable mortality burden.

Since wildfire was the major driver of the variations in total PM_2.5_ exposure and associated mortality risks, we compared our wildfire PM_2.5_ related mortality burden with those reported in previous studies. Ma et al. estimated around 11,400 deaths per year attributable to wildfire PM_2.5_ during 2007-2020, ^92^ while Zhang et al. reported approximately 24,100 deaths per year during 2006-2020 and Qiu et al. reported about 24,200 deaths per year during 2011-2020. ^93, 94^ Our estimate of approximately 12,300 deaths per year during 2018-2024 was therefore comparable to that of Ma et al. but approximately half the estimates reported by Zhang et al. and Qiu et al. ^92–94^ These differences may largely result from different exposure-response functions. The three previous studies applied their own developed wildfire-specific exposure-response functions, which generally indicated substantially higher health effects than those functions for all-source PM_2.5_. In a sensitivity analysis, Qiu et al. estimated roughly 6,000-9,000 deaths per year using several all-source PM_2.5_ exposure-response functions, substantially lower than 24,200 deaths estimated using wildfire-specific functions. ^93^ In the present study, applying the same all-source PM_2.5_ exposure–response function to wildfire and non-wildfire PM_2.5_ enabled a consistent comparison of the mortality burdens attributable to the two components. Nevertheless, this approach assumes equivalent toxicity and may yield conservative estimates of wildfire-related mortality if wildfire PM_2.5_ is more harmful than average total PM_2.5_. ^95^ Differences in study periods, wildfire PM_2.5_ exposure estimates, population, and baseline mortality data may also contribute to variation among the reported mortality burdens.

We then examined the spatial variability of wildfire-related PM_2.5_-related mortality. The county-level results in **Figure 5b** reveal that western U.S. counties suffered from a dramatically higher mortality burden due to wildfire PM_2.5_ than eastern U.S. counties in 2018, 2020, and 2021, whereas eastern U.S. counties experienced moderately higher mortality burdens attributable to wildfire PM_2.5_ in 2022 and 2023. While county-level patterns generally follow fire geography, spatially aggregated division-level estimates exhibited different patterns after considering population weighting.

Since severe wildfire smoke hit both western and eastern U.S. in 2021, we use this year as an illustrative example. The average concentration of wildfire PM_2.5_ in Pacific Division (2.31 μg/m^3^) was higher than that in Middle Atlantic (0.77 μg/m^3^) and West North Central (1.55 μg/m^3^) Divisions. However, the population-weighted average exposure of wildfire PM_2.5_ in Pacific Division (0.82 μg/m^3^) was comparable to that in Middle Atlantic Division (0.84 μg/m^3^) and substantially lower than that in West North Central Division (1.43 μg/m^3^). Consistent with these exposure patterns, the mortality rate due to wildfire PM_2.5_ in Pacific Division in 2021 (5.98 deaths per 100,000) was similar to that in Middle Atlantic Division (5.85) and much lower than West North Central Division (10.77). When averaged over 2018-2024, the highest wildfire-attributable mortality rates were observed in West North Central (6.80 deaths per 100,000) and East South Central (6.69) Divisions, rather than in Pacific Division (4.68). These findings highlight that the health impacts of wildfire smoke in recent years were comparable between eastern and western U.S. regions when evaluated from a population exposure perspective. This suggests that wildfire smoke should be considered a nationwide public health concern rather than solely a western U.S. environmental issue, and that public awareness and preparedness for wildfire smoke events should be strengthened across the entire country.

The separation of wildfire and non-wildfire related mortality burden (**Tables S7** and **S8**) is critical for public health policies, because different sources require fundamentally different regulatory and public health responses. Emission control strategies are highly effective for reducing anthropogenic PM_2.5_, while preventive measures to mitigate exposure should be emphasized for wildfire PM_2.5_. During severe wildfire smoke events, individuals are advised to stay indoors and limit outdoor activities. Keeping windows closed and using portable air cleaners with HEPA filters can further reduce indoor exposure. ^96^ When outdoor activities are unavoidable, wearing properly fitted N95 or equivalent respirators is strongly recommended. In addition, smoke forecasting and early warning systems should be strengthened by local governments to improve risk communication and enhance public health preparedness.

### 4.6 Strengths and limitations of this study

This study has several strengths. First, to the best of our knowledge, this is the first study to predict 1-km daily average concentrations of both total and wildfire-related PM_2.5_ across the CONUS with a focus on recent wildfire-influenced years. Many previous studies have covered long timeframes beginning in the early 2000s, during which anthropogenic emissions, rather than wildfire events, were the primary drivers of total PM_2.5_ variability. By contrast, our study focused on the 2018-2024 period, when wildfire activity increasingly dominated interannual variability in total PM_2.5_ concentrations.

Second, our machine learning framework incorporated predictors specifically relevant to wildfire and atmospheric chemistry. We included wildfire smoke, which many previous studies have not done. This focus allows for a clearer characterization of the growing influence of wildfire smoke on air quality under substantially reduced anthropogenic emissions. We also incorporated multiple chemically relevant precursor species from the GEOS-CF model, including key gaseous pollutants associated with secondary PM_2.5_ formation, which helped improve model performance and better capture the chemical processes influencing PM_2.5_ variability.

Third, few studies have simultaneously estimated the total and wildfire-related PM_2.5_ concentrations during recent wildfire-influenced years, which is essential for quantifying the contribution of wildfire smoke to total PM_2.5_ exposure. Future epidemiological studies can use this distinction between wildfire and non-wildfire PM_2.5_ to examine source-specific health effects.

Fourth, this study provided a comprehensive evaluation of model performance, including sample-, station-, and day-based CV metrics (i.e., R^2^, RMSE, MAE, MBE, and NRMSE) for daily, monthly, and annual average estimates, whereas previous studies only reported a more limited set of these metrics, making direct inter-study comparison challenging. Finally, our framework relied entirely on publicly accessible data sources, facilitating future updates and extensions, and we released the resulting prediction datasets to support further research on total and wildfire PM_2.5_ exposure and its health impacts.

However, several limitations should also be noted. First, the cross-validation results suggest that our model slightly underestimated extremely high total PM_2.5_ concentrations during wildfire episodes, a limitation also reported in previous ML-based PM_2.5_ prediction studies. Possible reasons include the relatively low frequency of episodic wildfire events, the use of mean squared error as the loss function, and the leaf-averaging effect in tree-based models. In addition, uncertainties in the HMS smoke product may affect the model’s ability to characterize wildfire-related PM_2.5_. Because HMS smoke polygons rely largely on daytime visible satellite imagery, they may fail to detect smaller or lower-intensity smoke plumes, including those from some prescribed fires. Furthermore, the two-dimensional smoke polygons provide limited information on plume altitude and therefore cannot reliably distinguish elevated smoke plumes from smoke that reaches the surface and contributes to ground-level PM_2.5_ concentrations.

Second, the proportional PAF approach was used in this study to separate the health impacts from wildfire-related and non-wildfire PM_2.5_. Although it ensures that the sum of source-specific health burdens equals the total attributable burden, it assumes that wildfire-related and non-wildfire PM_2.5_ share the same exposure-response relationships with all-cause mortality. While some studies have suggested potentially differentiated health effects of wildfire-related PM_2.5_, ^95^ more epidemiological evidence is required to better characterize source-specific health risks.

Third, this study only focused on the long-term health impacts of total and wildfire-related PM_2.5_. Future studies could further investigate short-term health impacts, especially those associated with episodic wildfire-related PM_2.5_ peak exposures.

## 5 Conclusion

This study developed a high-resolution machine learning framework to estimate daily 1-km concentrations of total and wildfire-related PM_2.5_ across the CONUS from 2018 to 2024. The model had a good predictive performance and revealed substantial spatiotemporal variability in total PM_2.5_ pollution during recent wildfire-influenced years. The U.S. west coast experienced the most severe episodic pollution events, while many metropolitan areas across both western and eastern regions had considerable numbers of highly polluted days. Wildfire smoke contributed substantially to total PM_2.5_ concentrations during major wildfire years, accounting for approximately 30-35% of total PM_2.5_ along the west coast during 2020-2021, and over 25% in the eastern U.S. in 2023. In severe wildfire years, wildfire smoke also contributed over 20% of population exposure in both western and eastern regions, highlighting its growing influence on air quality nationwide.

For long-term health impacts, the mortality burden attributable to total PM_2.5_ peaked in 2021 (62.7 per 100,000), while wildfire-related PM_2.5_ mortality was highest in 2023 (10.23 per 100,000), accounting for 18.4% of total PM_2.5_-related deaths. Notably, when evaluated from a population exposure perspective, the health impacts of wildfire smoke were comparable between eastern and western U.S. regions, underscoring the importance of recognizing wildfire smoke as a nationwide public health concern rather than a regionally confined issue.

## CRediT authorship contribution statement

**Ningrui Liu:** Conceptualization, Methodology, Formal analysis, Software, Visualization, Writing – original draft. **Yeonseung Cheong:** Formal analysis, Writing – original draft. **Timothy Larson:** Supervision, Writing – review & editing. **Elena Austin:** Conceptualization, Methodology, Supervision, Writing – review & editing. **Edmund Seto:** Conceptualization, Methodology, Supervision, Writing – review & editing.

## Acknowledgements

We thank Prof. Marissa Childs of the Department of Environmental & Occupational Health Sciences at the University of Washington for her helpful suggestions on this project.

## Data availability

Datasets of daily, monthly, and annual average total and wildfire PM_2.5_ concentrations at 1-km spatial resolution are available at https://zenodo.org/records/24094871.

