## Supplementary Material 1 for "High-resolution spatiotemporal estimates of total and wildfire PM_2.5_ in the contiguous United States, 2018-2024: Implications for population exposure and mortality burden"

**Note:** **Tables S7 and S8** are available in the ***Supporting Information* 2**.

**Section S1 Machine learning model details**

Compared with the classical extra-trees model, the “four-dimensional space-time” framework additionally incorporates the Euclidean spherical spatial coordinates and triangular spiral temporal representations to account for spatiotemporal heterogeneity. Specifically, the spatial terms *Ps*=[*S*_1_, *S*_2_, *S*_3_] are defined as ^1^

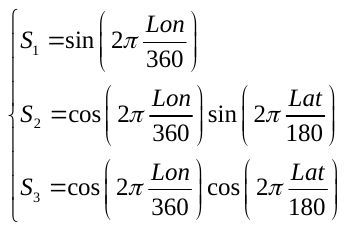
(S1)

where *Lon* and *Lat* refer to the longitude and latitude of the 1-km grid centroid, respectively. The temporal terms *Pt*=[*T*_1_, *T*_2_, *T*_3_] are defined as ^1^

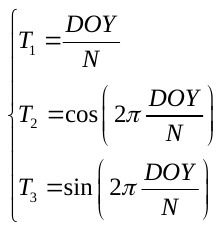
(S2)

where *DOY* is the day of year (i.e., 1, 2, 3, …, 365(, 366)), and *N* is the total number of days in this year.

To address the missingness of satellite AOD data, we adopted a two-stage modeling strategy ^1^. The first stage fit the 4D-STET model to fill the AOD gaps using simulated AOD and meteorological data, as shown in Equation (S3) below.

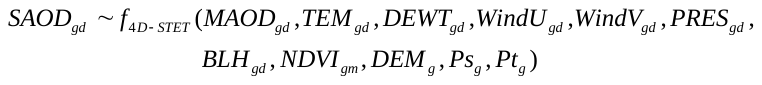
(S3)

where *SAOD* is the satellite AOD, *MAOD* is the modeled AOD from GEOS-CF, *TEM* is temperature, *DEWT* is dewpoint temperature, *WindU* and *WindV* is eastward and northward component of wind speed, *PRES* is surface pressure, *BLH* is boundary layer height, *NDVI* is normalized difference vegetation index, *DEM* is elevation, and *Ps* and *Pt* are spatial and temporal terms; the subscripts *g* and *d* refer to grid and day, respectively. Since the sample size of grids with available SAOD data was too huge and SAOD values in adjacent grids and days are highly correlated, it is unnecessary to include all available data to fit the 4D-STET model. Specifically, 200 grids with SAOD data per day were selected using the “regular” method (i.e., sample points are placed at evenly spaced intervals across the spatial extent of the object) to form the final training dataset, which included 502,668 data points in total.

Then the second stage applied the 4D-STET model with the gap-filled AOD, all meteorological data, population, nighttime lights, all land-use variables, and wildfire smoke density to predict ground-level daily average PM_2.5_ concentrations.

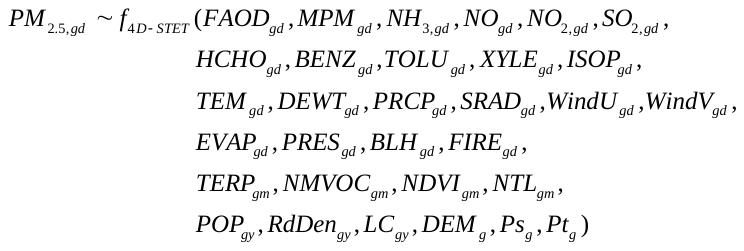
(S4)

where *FAOD* is the gap-filled AOD predicted from the step-1 model above, *MPM* is the GEOS-CF simulated PM_2.5_ concentration, *NH*_3_ is the GEOS-CF simulated ammonia concentration, *NO* is the GEOS-CF simulated nitrogen oxide concentration, *NO*_2_ is the GEOS-CF simulated nitrogen dioxide concentration, *SO*_2_ is the GEOS-CF simulated sulfur dioxide concentration, *HCHO* is the GEOS-CF simulated formaldehyde concentration, *BENZ* is the GEOS-CF simulated benzene concentration, *TOLU* is the GEOS-CF simulated toluene concentration, *XYLE* is the GEOS-CF simulated xylene concentration, *ISOP* is the GEOS-CF simulated isoprene concentration, *PRCP* is the total precipitation, *SRAD* is the total surface solar radiation downwards, *EVAP* is the total evaporation, *FIRE* is the wildfire smoke density, *TERP* is the anthropogenic emission amount of monoterpenes, *NMVOC* is the anthropogenic emission of non-methane VOCs, *NTL* is the nighttime light radiance, *POP* is the population counts, *RdDen* is the road density of S1100, S1200, and S1400 categories, and *LC* is the sum of area of 16 land cover types separately; the subscripts *g*, *m*, *d*, and *y* refer to grid, day, month, and year, respectively. More details of these predictors are available in **Table S1**. A total of 1,564,740 data points were included in this step-2 model.

There are three major hyperparameters to be tuned in the extra-trees model using ranger package in R: mtry (number of variables to possibly split at in each node), min.node.size (minimal node size to split at), and sample.fraction (fraction of observations to sample). Another hyperparameter, i.e., num.trees (number of trees), was set as 500 and not tuned here, because we found that further increasing the number of trees from 500 to 1000 did not meaningfully improve model performance. For each step of the model, we generated 100 hyperparameter vectors at random using a maxmin Latin-hypercube sampling approach, and fitted the 4D-STET model using each hyperparameter vector separately. Finally, we selected the optimal hyperparameters based on the minimum out-of-bag (OOB) error. The optimal hyperparameters in the two steps are shown in **Table S2**.

With the optimal hyperparameters, the final 4D-STET model was evaluated by 10-fold sample-based, station-based, and day-based cross-validation (CV) as introduced in the manuscript. ^1^^,^ ^2^ Here we show the formulas of the evaluation metrics used in these CV approaches:

(1) Coefficient of determination (R^2^)

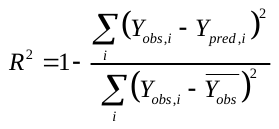
(S5)

where *Y_obs_* and *Y_pred_* are observed values and model predictions, respectively;
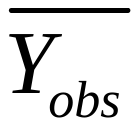
 is the overall mean of all observations.

(2) Root mean squared error (RMSE) and normalized RMSE (NRMSE)

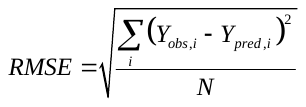
(S6)

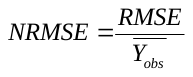
(S7)

(3) Mean absolute error (MAE)

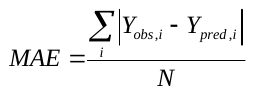
(S8)

(4) Mean bias error (MBE)

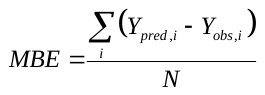
(S9)

**Section S2 Uncertainty analysis**

This section provides more details for the two-dimensional Monte Carlo simulation used in the uncertainty analysis. ^3-5^ The first dimension reflects the exposure concentration distribution due to intrapopulation variability within each county. In this dimension, 1000 concentrations were randomly sampled from all 1-km grids within each county weighted by grid-level population, i.e., *C*_1_, ……, *C*_1000_. The second dimension shows the uncertainty in the exposure-response relationships. In this dimension, 1000 relative risks per unit increase of PM_2.5_ exposure (*RR*_0_ = 1.12 (1.08 – 1.15)) were randomly sampled from the log-normal distribution of *RR*_0_ (log *RR*_0_ is approximately normally distributed when sample size is large), i.e., *RR*_0,1_, ……, *RR*_0,1000_. According to the exposure-response relationship in equation (5) in the main text, a total of 1,000,000 relative risks can be obtained (*RR_i_*_,_*_j_* for the *i*^th^ *RR*_0_ and the *j*^th^ concentration).

***Schematic of the process of a two-stage Monte Carlo simulation.***

|  | *C*_1_ | *C*_2_ | …… | *C*_1000_ | 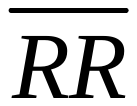 | 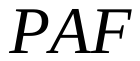 |
| --- | --- | --- | --- | --- | --- | --- |
| ***RR*_0,1_** | *RR*_1,1_ | *RR*_1,2_ | …… | *RR*_1,1000_ | 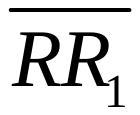 | 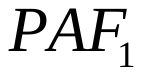 |
| ***RR*_0,2_** | *RR*_2,1_ | *RR*_2,2_ | …… | *RR*_2,1000_ | 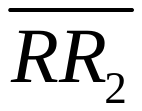 | 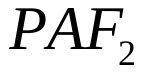 |
| **……** | …… | …… | …… | …… | …… | …… |
| ***RR*_0,1000_** | *RR*_1000,1_ | *RR*_1000,2_ | …… | *RR*_1000,1000_ | 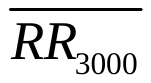 | 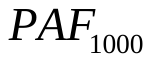 |

Then, we averaged each row of *RR_i_*_,_*_j_* to obtain 1000 average relative risks, so as to average the variability and retain the uncertainty in the exposure-response relationships. Then the PAF can be estimated for each row, which gives a total of 1000 PAFs. Using these 1000 PAFs, the 1000 attributable mortality burden can be estimated, which formed the distribution of itself. Finally, the mean and 2.5^th^ to 97.5^th^ percentiles of the 1000 simulated mortality burden are reported as the mean and 95% UI.

All spatial analysis and statistical analyses described in the machine learning and uncertainty analysis section were performed with R V4.5.2 using the sf, ^6^^,^ ^7^ terra, ^8^ ranger, ^9^ dplyr, ^10^ stringr, ^11^ reshape2, ^12^ lhs, ^13^ fastshap, ^14^ pdp, ^15^ tigris, ^16^ and tidycensus packages. ^17^

**Table S1.** Summary of datasets and sources used in the machine learning models.

| Variable | Content | Unit | Spatial coverage | Spatial resolution | Temporal coverage | Temporal resolution | Data source |
| --- | --- | --- | --- | --- | --- | --- | --- |
| PM_2.5_ data | | | | | | | |
| PM2.5 | Ground-level PM_2.5_ measurements | μg/m^3^ | U.S. | Site | 1980-2025 | Hourly | US EPA ^18^ |
| MPM2.5 | Model PM_2.5_ | μg/m^3^ | Global | 0.25°🞨0.25° | 2018-2025 | Hourly | GEOS-CF ^19^ |
| AOD data | | | | | | | |
| SAOD | Satellite AOD (550 nm) | - | Global | 1km🞨1km | 2000-2025 | Daily | MCD19A2 ^20^ |
| MAOD | Model AOD from CTM | - | Global | 0.25°🞨0.25° | 2018-2025 | Hourly | GEOS-CF ^19^ |
| Precursor data from CTMs | | | | | | | |
| NH3 | Model ammonia | mol frac | Global | 0.25°🞨0.25° | 2018-2025 | Hourly | GEOS-CF ^19^ |
| NO | Model nitrogen oxide | mol frac | Global | 0.25°🞨0.25° | 2018-2025 | Hourly | GEOS-CF ^19^ |
| NO2 | Model nitrogen dioxide | mol frac | Global | 0.25°🞨0.25° | 2018-2025 | Hourly | GEOS-CF ^19^ |
| SO2 | Model sulfur dioxide | mol frac | Global | 0.25°🞨0.25° | 2018-2025 | Hourly | GEOS-CF ^19^ |
| HCHO | Model formaldehyde | mol frac | Global | 0.25°🞨0.25° | 2018-2025 | Hourly | GEOS-CF ^19^ |
| BENZ | Model benzene | mol frac | Global | 0.25°🞨0.25° | 2018-2025 | Hourly | GEOS-CF ^19^ |
| TOLU | Model toluene | mol frac | Global | 0.25°🞨0.25° | 2018-2025 | Hourly | GEOS-CF ^19^ |
| XYLE | Model xylene | mol frac | Global | 0.25°🞨0.25° | 2018-2025 | Hourly | GEOS-CF ^19^ |
| ISOP | Model isoprene | mol frac | Global | 0.25°🞨0.25° | 2018-2025 | Hourly | GEOS-CF ^19^ |
| TERP | Anthropogenic emission of monoterpenes | kg/m2/s | Global | 0.1°🞨0.1° | 2000-2025 | Monthly | CAMS global emission inventories ^21^ |
| NMVOC | Anthropogenic emission of non-methane VOCs | kg/m2/s | Global | 0.1°🞨0.1° | 2000-2025 | Monthly | CAMS global emission inventories ^21^ |
| Meteorological data | | | | | | | |
| TEM | Temperature at 2m | K | Global | 0.1°🞨0.1° | 1950-2025 | Hourly | ERA5-Land ^22^ |
| DEWT | Dew point temperature at 2m | K | Global | 0.1°🞨0.1° | 1950-2025 | Hourly | ERA5-Land ^22^ |
| PRCP | Total sum of precipitation | mm | Global | 0.1°🞨0.1° | 1950-2025 | Hourly | ERA5-Land ^22^ |
| SRAD | Total surface solar radiation downwards | J/m2 | Global | 0.1°🞨0.1° | 1950-2025 | Hourly | ERA5-Land ^22^ |
| WindU | u-component of wind speed | m/s | Global | 0.1°🞨0.1° | 1950-2025 | Hourly | ERA5-Land ^22^ |
| WindV | v-component of wind speed | m/s | Global | 0.1°🞨0.1° | 1950-2025 | Hourly | ERA5-Land ^22^ |
| EVAP | Total sum of evaporation | mm | Global | 0.1°🞨0.1° | 1950-2025 | Hourly | ERA5-Land ^22^ |
| PRES | Surface pressure | Pa | Global | 0.1°🞨0.1° | 1950-2025 | Hourly | ERA5-Land ^22^ |
| BLH | Boundary layer height | m | Global | 0.25°🞨0.25° | 1940-2025 | Hourly | ERA5 ^23^ |
| Sociodemographic data | | | | | | | |
| POP | Population | Count | Global | 1km🞨1km | 2015-2030 | Annual | WorldPop ^24^ |
| NTL | Nighttime lights | nW/sr/cm^2^ | Global | 500m🞨500m | 2012-2025 | Monthly | VIIRS ^25^ |
| Land-use data | | | | | | | |
| NDVI | Normalized difference vegetation index | - | Global | 1km🞨1km | 2000-2025 | Monthly | MOD13A3 ^26^ |
| LC (multiple) | Area of different land cover types in the grid | m2 | CONUS | 30m🞨30m | 1985-2024 | Annual | NLCD ^27^ |
| RdDen | Road density (S1100, S1200, S1400) | km/km^2^ | US | Vector | 2012-2025 | Annual | TIGER ^28^ |
| DEM | Mean of surface elevation | m | Global | 90m🞨90m | - | - | SRTM ^29^ |
| Wildfire data | | | | | | | |
| FIRE | HMS wildfire smoke density | Categorical | CONUS | Polygon | 2003-2025 | Daily | HMS NOAA ^30^ |

**Notes:** The links of the above data sources are listed below.

1. EPA AQS data: <https://aqs.epa.gov/aqsweb/airdata/download_files.html>
2. MCD19A2 (Terra & Aqua MAIAC Land Aerosol Optical Depth): <https://developers.google.com/earth-engine/datasets/catalog/MODIS_061_MCD19A2_GRANULES?hl=en>
3. GEOS-CF (GEOS Composition Forecasting): <https://developers.google.com/earth-engine/datasets/catalog/NASA_GEOS-CF_v1_rpl_tavg1hr?hl=en>
4. CAMS (Copernicus Atmosphere Monitoring Service) global emission inventories: <https://ads.atmosphere.copernicus.eu/datasets/cams-global-emission-inventories?tab=overview>
5. ERA5 reanalysis data: <https://developers.google.com/earth-engine/datasets/catalog/ECMWF_ERA5_HOURLY?hl=en>
6. ERA5-land reanalysis data: <https://developers.google.com/earth-engine/datasets/catalog/ECMWF_ERA5_LAND_DAILY_AGGR?hl=en>
7. WorldPop data: <https://hub.worldpop.org/project/categories?id=3>
8. VIIRS (Visible Infrared Imaging Radiometer Suite): <https://developers.google.com/earth-engine/datasets/catalog/NOAA_VIIRS_DNB_MONTHLY_V1_VCMCFG?hl=en#bands>
9. MOD13A3 (MODIS/Terra Vegetation Indices): <https://developers.google.com/earth-engine/datasets/catalog/MODIS_061_MOD13A3?hl=en>
10. NLCD (National Land Cover Database): <https://www.sciencebase.gov/catalog/item/6810c1a4d4be022940554075>
11. TIGER (Topologically Integrated Geographic Encoding and Referencing): <https://gee-community-catalog.org/projects/tiger_roads/>
12. SRTM (Shuttle Radar Topography Mission): <https://developers.google.com/earth-engine/datasets/catalog/CGIAR_SRTM90_V4?hl=en>
13. NOAA Hazard Mapping System (HMS) Fire and Smoke Product: <https://www.ospo.noaa.gov/products/land/hms.html#data>

**Table S2.** Hyperparameters used in the 4D-STET model.

| Hyperparameter | Range | Optimal value in Step 1 | Optimal value in Step 2 |
| --- | --- | --- | --- |
| mtry | 3~(*P*-1) | 5 | 41 |
| min.node.size | 1~20 | 2 | 3 |
| sample.fraction | 0.5~1.0 | 0.74 | 0.90 |

Note: *P* is the number of predictors in the machine learning model.

**Table S3.** Model performance from cross-validation in this study and previous studies. The R^2^ and NRMSE are dimensionless, and the units of RMSE, MAE, and MBE are μg/m^3^.

| Study | Time | Location | CV type | Daily | | | | | Monthly | | | | | Annual | | | | |
| --- | --- | --- | --- | --- | --- | --- | --- | --- | --- | --- | --- | --- | --- | --- | --- | --- | --- | --- |
|  |  |  |  | R^2^ | RMSE | MAE | MBE | NRMSE | R^2^ | RMSE | MAE | MBE | NRMSE | R^2^ | RMSE | MAE | MBE | NRMSE |
| This study | **2018-2024** | **CONUS** | **Sample** | **0.72** | **2.76** | **1.71** | **0.08** | **0.38** | **0.86** | **1.18** | **0.83** | **0.08** | **0.16** | **0.89** | **0.73** | **0.54** | **0.09** | **0.10** |
|  |  |  | **Station** | **0.63** | **3.22** | **2.02** | **0.15** | **0.44** | **0.65** | **1.87** | **1.27** | **0.15** | **0.26** | **0.62** | **1.38** | **0.98** | **0.15** | **0.19** |
|  |  |  | **Day** | **0.63** | **3.20** | **1.98** | **0.10** | **0.44** | **0.84** | **1.27** | **0.90** | **0.10** | **0.17** | **0.88** | **0.77** | **0.58** | **0.10** | **0.11** |
| Wei et al. ^1^ | 2017-2022 | North America | Station | 0.76 | 4.62 | 2.53 |  |  |  |  |  |  |  |  |  |  |  |  |
|  |  | Global | Sample | 0.91 | 9.20 | 4.09 |  | 0.37 | 0.97 | 4.15 | 2.21 |  | 0.17 | 0.98 | 2.77 | 1.62 |  | 0.11 |
|  |  |  | Station | 0.87 | 10.94 |  |  | 0.44 |  |  |  |  |  |  |  |  |  |  |
|  |  |  | Day | 0.81 | 13.49 |  |  | 0.54 |  |  |  |  |  |  |  |  |  |  |
| Wei et al. ^2^ | 2000-2020 | CONUS | Sample | 0.82 |  |  |  | 0.40 | 0.97 |  |  |  | 0.11 | 0.99 |  |  |  | 0.07 |
|  |  |  | Station | 0.72 |  |  |  | 0.50 | 0.81 |  |  |  | 0.28 | 0.80 |  |  |  | 0.26 |
|  |  |  | Day | 0.74 |  |  |  | 0.48 | 0.96 |  |  |  | 0.13 | 0.98 |  |  |  | 0.08 |
| Di et al. ^31^ | 2000-2015 | CONUS | Station | 0.86 | 2.79 |  |  |  |  |  |  |  |  | 0.89 |  |  |  |  |
| Just et al. ^32^ | 2003-2023 | CONUS | Station |  |  | 2.09 | -0.21 |  |  |  |  |  |  |  |  |  |  |  |
| Wang et al. ^33^ | 2005-2021 | CONUS | Sample | 0.74 | 3.67 |  | -0.09 |  |  |  |  |  |  |  |  |  |  |  |
| Kayastha et al. ^34^ | 2018-2022 | CONUS | Sample | 0.83 | 3.00 | 1.61 |  | 0.37 |  |  |  |  |  |  |  |  |  |  |
|  |  |  | Station | 0.72 | 3.81 | 1.97 |  | 0.47 |  |  |  |  |  |  |  |  |  |  |
| Shen et al. ^35^ | 2000-2019 | North America | Station |  |  |  |  |  | 0.50-0.61 |  |  |  | 0.29-0.35 | 0.57 | 2.00 |  |  | 0.25 |
| van Donkelaar et al.^36^ | 2000-2022 | North America | Sample | 0.62 | 4.20 |  |  | 0.44 |  |  |  |  |  |  |  |  |  |  |
| Hu et al. ^37^ | 2011 | CONUS | Sample | 0.80 | 2.83 | 1.78 |  |  |  |  |  |  |  |  |  |  |  |  |
|  |  |  | Station | 0.70 | 3.43 | 2.24 |  |  |  |  |  |  |  |  |  |  |  |  |
|  |  |  | Day | 0.79 | 2.88 | 1.81 |  |  |  |  |  |  |  |  |  |  |  |  |
| Reid et al. ^38^ | 2008-2018 | 11 states in west U.S. | Sample | 0.72 | 5.48 |  |  | 0.67 |  |  |  |  |  |  |  |  |  |  |
|  |  |  | Station | 0.60 | 6.58 |  |  | 0.80 |  |  |  |  |  |  |  |  |  |  |
| Swanson et al. ^39^ | 2003-2021 | West U.S. | Station | 0.65 | 5.84 | 2.72 |  |  |  |  |  |  |  |  |  |  |  |  |
| Zhang et al. ^40^ | 2007-2018 | CONUS | Sample | 0.75 | 4.59 |  |  |  | 0.84 | 2.66 |  |  |  | 0.90 |  |  |  |  |
|  |  |  | Spatial | 0.59 | 5.88 |  |  |  | 0.67 | 3.82 |  |  |  |  |  |  |  |  |
|  |  |  | Temporal | 0.67 | 5.18 |  |  |  | 0.79 | 3.00 |  |  |  |  |  |  |  |  |

Note:

(1) Kayastha et al. used 4-km spatial resolution instead of 1-km resolution. ^34^
(2) van Donkelaar et al. used biweekly average instead of daily average estimates ^36^.
(3) Reid et al. predicted daily PM_2.5_ at the census tract level. ^38^
(4) Swanson et al. used leave-one-out cross-validation (LOOCV) instead of 10-fold CV. ^39^
(5) Zhang et al. used a 20-fold CV instead of a 10-fold CV. ^40^

**Table S4.** Division- and year-specific sample-based CV metrics of the daily average predictions.

| Division/Year | R^2^ | RMSE | MAE | MBE | NRMSE |
| --- | --- | --- | --- | --- | --- |
| New England | 0.74 | 2.10 | 1.44 | 0.11 | 0.35 |
| Middle Atlantic | 0.74 | 2.33 | 1.56 | -0.01 | 0.31 |
| South Atlantic | 0.68 | 2.32 | 1.55 | 0.13 | 0.32 |
| East South Central | 0.64 | 2.24 | 1.59 | 0.20 | 0.30 |
| West South Central | 0.66 | 3.18 | 2.00 | 0.01 | 0.36 |
| East North Central | 0.72 | 2.39 | 1.62 | 0.02 | 0.30 |
| West North Central | 0.66 | 2.66 | 1.71 | 0.15 | 0.41 |
| Mountain | 0.71 | 2.71 | 1.59 | 0.10 | 0.45 |
| Pacific | 0.75 | 3.72 | 2.14 | 0.09 | 0.47 |
| 2018 | 0.75 | 3.04 | 1.92 | -0.26 | 0.39 |
| 2019 | 0.71 | 2.51 | 1.68 | 0.07 | 0.35 |
| 2020 | 0.73 | 3.15 | 1.77 | 0.12 | 0.43 |
| 2021 | 0.72 | 2.91 | 1.77 | -0.04 | 0.37 |
| 2022 | 0.71 | 2.50 | 1.62 | 0.13 | 0.35 |
| 2023 | 0.72 | 2.76 | 1.71 | 0.10 | 0.38 |
| 2024 | 0.70 | 2.54 | 1.61 | 0.32 | 0.37 |

Note: The units of RMSE, MAE, and MBE are μg/m^3^, and R^2^ and NRMSE are dimensionless.

**Table S5.** Annual average total PM_2.5_ concentrations (unweighted) in nine divisions across the CONUS.

| Division | Mean (SD) | 2018 | 2019 | 2020 | 2021 | 2022 | 2023 | 2024 |
| --- | --- | --- | --- | --- | --- | --- | --- | --- |
| Annual average total PM_2.5_ concentrations (μg/m^3^) | | | | | | | | |
| New England | 4.69 (0.32) | 4.51 | 4.34 | 4.55 | 4.82 | 4.51 | 5.41 | 4.73 |
| Middle Atlantic | 6.22 (0.47) | 6.02 | 6.01 | 5.80 | 6.50 | 5.95 | 7.26 | 5.97 |
| East North Central | 7.05 (0.43) | 7.01 | 6.97 | 6.67 | 7.49 | 6.69 | 7.85 | 6.64 |
| West North Central | 6.20 (0.47) | 6.07 | 5.70 | 5.86 | 6.83 | 5.73 | 6.95 | 6.28 |
| South Atlantic | 6.93 (0.32) | 6.61 | 6.98 | 6.53 | 7.02 | 6.83 | 7.60 | 6.96 |
| East South Central | 7.25 (0.30) | 7.11 | 7.27 | 6.81 | 7.28 | 7.17 | 7.88 | 7.23 |
| West South Central | 7.72 (0.22) | 7.54 | 7.39 | 7.59 | 7.68 | 7.82 | 8.00 | 8.02 |
| Mountain | 4.80 (0.57) | 4.92 | 3.98 | 5.21 | 5.87 | 4.43 | 4.40 | 4.79 |
| Pacific | 6.82 (0.91) | 7.36 | 5.69 | 8.27 | 7.83 | 6.22 | 6.12 | 6.27 |
| Annual average wildfire PM_2.5_ concentrations (μg/m^3^) | | | | | | | | |
| New England | 0.53 (0.43) | 0.20 | 0.17 | 0.13 | 0.61 | 0.36 | 1.40 | 0.85 |
| Middle Atlantic | 0.64 (0.59) | 0.19 | 0.16 | 0.11 | 0.77 | 0.44 | 1.90 | 0.91 |
| East North Central | 0.77 (0.56) | 0.35 | 0.23 | 0.26 | 1.07 | 0.68 | 1.92 | 0.90 |
| West North Central | 1.00 (0.55) | 0.61 | 0.26 | 0.61 | 1.55 | 0.82 | 1.92 | 1.25 |
| South Atlantic | 0.64 (0.49) | 0.11 | 0.23 | 0.13 | 0.74 | 0.81 | 1.58 | 0.87 |
| East South Central | 0.73 (0.52) | 0.15 | 0.34 | 0.18 | 0.79 | 1.04 | 1.71 | 0.89 |
| West South Central | 0.81 (0.47) | 0.17 | 0.29 | 0.48 | 0.76 | 1.35 | 1.41 | 1.17 |
| Mountain | 0.86 (0.53) | 0.86 | 0.10 | 1.25 | 1.85 | 0.54 | 0.52 | 0.87 |
| Pacific | 1.31 (0.93) | 1.74 | 0.15 | 2.85 | 2.31 | 0.64 | 0.64 | 0.85 |
| Annual average wildfire contribution (%) | | | | | | | | |
| New England | 10.80 (7.95) | 4.36 | 3.89 | 2.86 | 12.69 | 7.95 | 25.87 | 18.01 |
| Middle Atlantic | 9.76 (8.15) | 3.14 | 2.60 | 1.95 | 11.78 | 7.41 | 26.18 | 15.25 |
| East North Central | 10.66 (7.02) | 4.99 | 3.24 | 3.91 | 14.26 | 10.17 | 24.47 | 13.58 |
| West North Central | 15.65 (7.51) | 10.03 | 4.57 | 10.42 | 22.77 | 14.35 | 27.60 | 19.85 |
| South Atlantic | 8.95 (6.51) | 1.71 | 3.32 | 1.93 | 10.61 | 11.83 | 20.74 | 12.54 |
| East South Central | 9.82 (6.63) | 2.10 | 4.64 | 2.68 | 10.78 | 14.52 | 21.68 | 12.37 |
| West South Central | 10.28 (5.86) | 2.30 | 3.99 | 6.32 | 9.85 | 17.25 | 17.60 | 14.64 |
| Mountain | 16.82 (8.64) | 17.57 | 2.47 | 23.93 | 31.54 | 12.09 | 11.88 | 18.24 |
| Pacific | 17.80 (10.7) | 23.63 | 2.64 | 34.48 | 29.50 | 10.32 | 10.48 | 13.58 |

**Note:** The annual average wildfire contribution here is obtained from dividing annual average wildfire PM_2.5_ by annual average total PM_2.5_ for each division separately, instead of the average of 1-km ratios.

**Table S6.** Population-weighted annual average total PM_2.5_ exposures in nine divisions across the CONUS.

| Division | Mean (SD) | 2018 | 2019 | 2020 | 2021 | 2022 | 2023 | 2024 |
| --- | --- | --- | --- | --- | --- | --- | --- | --- |
| Population-weighted annual average total PM_2.5_ exposures (μg/m^3^) | | | | | | | | |
| New England | 6.10 (0.33) | 6.09 | 5.88 | 5.99 | 6.26 | 5.84 | 6.82 | 5.84 |
| Middle Atlantic | 7.52 (0.54) | 7.51 | 7.39 | 7.06 | 7.76 | 7.11 | 8.71 | 7.11 |
| East North Central | 8.13 (0.42) | 8.18 | 8.21 | 7.73 | 8.55 | 7.77 | 8.85 | 7.62 |
| West North Central | 7.38 (0.47) | 7.34 | 7.09 | 7.00 | 7.94 | 6.88 | 8.24 | 7.20 |
| South Atlantic | 7.37 (0.30) | 7.18 | 7.46 | 6.99 | 7.46 | 7.19 | 7.99 | 7.33 |
| East South Central | 7.65 (0.30) | 7.54 | 7.66 | 7.20 | 7.79 | 7.51 | 8.26 | 7.57 |
| West South Central | 8.82 (0.29) | 8.63 | 8.53 | 8.59 | 8.69 | 8.75 | 9.26 | 9.27 |
| Mountain | 6.83 (0.50) | 6.91 | 6.09 | 7.44 | 7.58 | 6.50 | 6.46 | 6.83 |
| Pacific | 9.46 (0.78) | 10.31 | 8.64 | 10.85 | 9.59 | 9.17 | 8.67 | 8.95 |
| Population-weighted annual average wildfire PM_2.5_ exposures (μg/m^3^) | | | | | | | | |
| New England | 0.57 (0.48) | 0.23 | 0.18 | 0.10 | 0.65 | 0.39 | 1.58 | 0.83 |
| Middle Atlantic | 0.71 (0.67) | 0.22 | 0.16 | 0.11 | 0.84 | 0.48 | 2.17 | 0.97 |
| East North Central | 0.78 (0.58) | 0.30 | 0.20 | 0.26 | 1.02 | 0.71 | 1.95 | 1.02 |
| West North Central | 0.99 (0.61) | 0.48 | 0.28 | 0.46 | 1.43 | 0.91 | 2.13 | 1.21 |
| South Atlantic | 0.57 (0.47) | 0.09 | 0.18 | 0.10 | 0.65 | 0.68 | 1.50 | 0.80 |
| East South Central | 0.72 (0.51) | 0.16 | 0.31 | 0.17 | 0.85 | 1.00 | 1.69 | 0.84 |
| West South Central | 0.91 (0.60) | 0.16 | 0.32 | 0.44 | 0.75 | 1.43 | 1.78 | 1.47 |
| Mountain | 0.62 (0.44) | 0.59 | 0.05 | 1.20 | 1.32 | 0.29 | 0.31 | 0.55 |
| Pacific | 0.75 (0.67) | 1.18 | 0.09 | 2.17 | 0.82 | 0.35 | 0.36 | 0.28 |
| Population-weighted annual average wildfire contribution (%) | | | | | | | | |
| New England | 9.00 (7.07) | 3.82 | 3.09 | 1.75 | 10.39 | 6.62 | 23.19 | 14.13 |
| Middle Atlantic | 8.99 (7.78) | 2.98 | 2.17 | 1.54 | 10.89 | 6.75 | 24.96 | 13.63 |
| East North Central | 9.42 (6.54) | 3.72 | 2.39 | 3.31 | 11.96 | 9.17 | 22.01 | 13.34 |
| West North Central | 13.00 (7.26) | 6.48 | 3.99 | 6.61 | 18.05 | 13.26 | 25.84 | 16.75 |
| South Atlantic | 7.60 (5.92) | 1.30 | 2.43 | 1.50 | 8.76 | 9.50 | 18.82 | 10.88 |
| East South Central | 9.20 (6.26) | 2.09 | 4.10 | 2.36 | 10.98 | 13.35 | 20.48 | 11.05 |
| West South Central | 10.10 (6.43) | 1.88 | 3.76 | 5.06 | 8.68 | 16.28 | 19.20 | 15.88 |
| Mountain | 8.60 (5.67) | 8.58 | 0.82 | 16.08 | 17.37 | 4.51 | 4.82 | 8.04 |
| Pacific | 7.45 (6.08) | 11.45 | 1.08 | 20.03 | 8.55 | 3.83 | 4.11 | 3.13 |

**Note:** The population-weighted annual average wildfire contribution here is obtained from dividing population-weighted annual average wildfire PM_2.5_ by population-weighted annual average total PM_2.5_ for each division separately, instead of the population-weighted average of 1-km ratios.

**Figure S1.** **Model performance in the 10-fold station-based cross-validation (CV):** Density scatterplots and related out-of-sample metrics of station-based CV results for (*a*) daily, (*b*) monthly, and (*c*) annual average predictions.

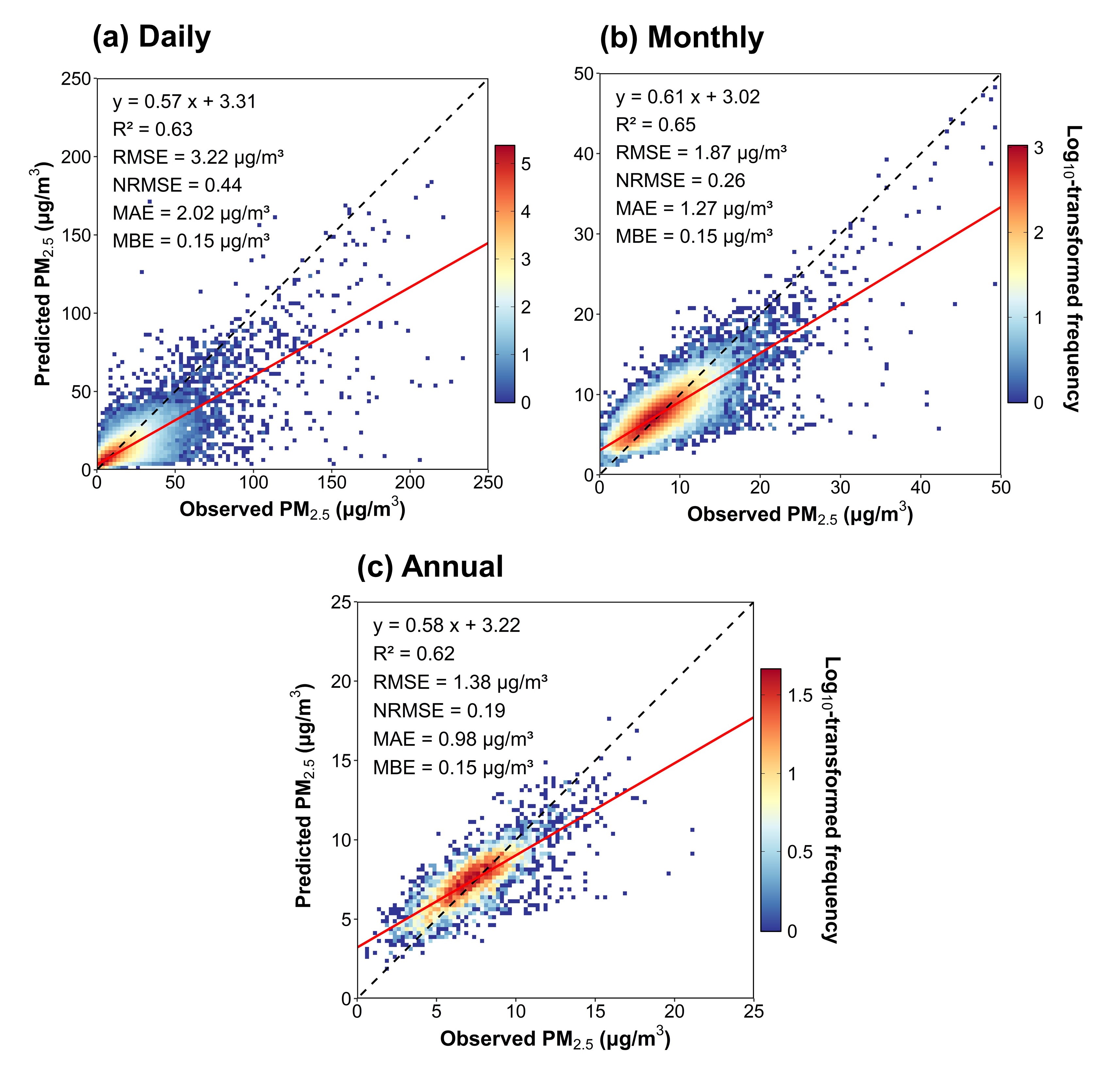

**Figure S2.** Mean absolute SHAP values for the top 15 features in the 4D-STET model.

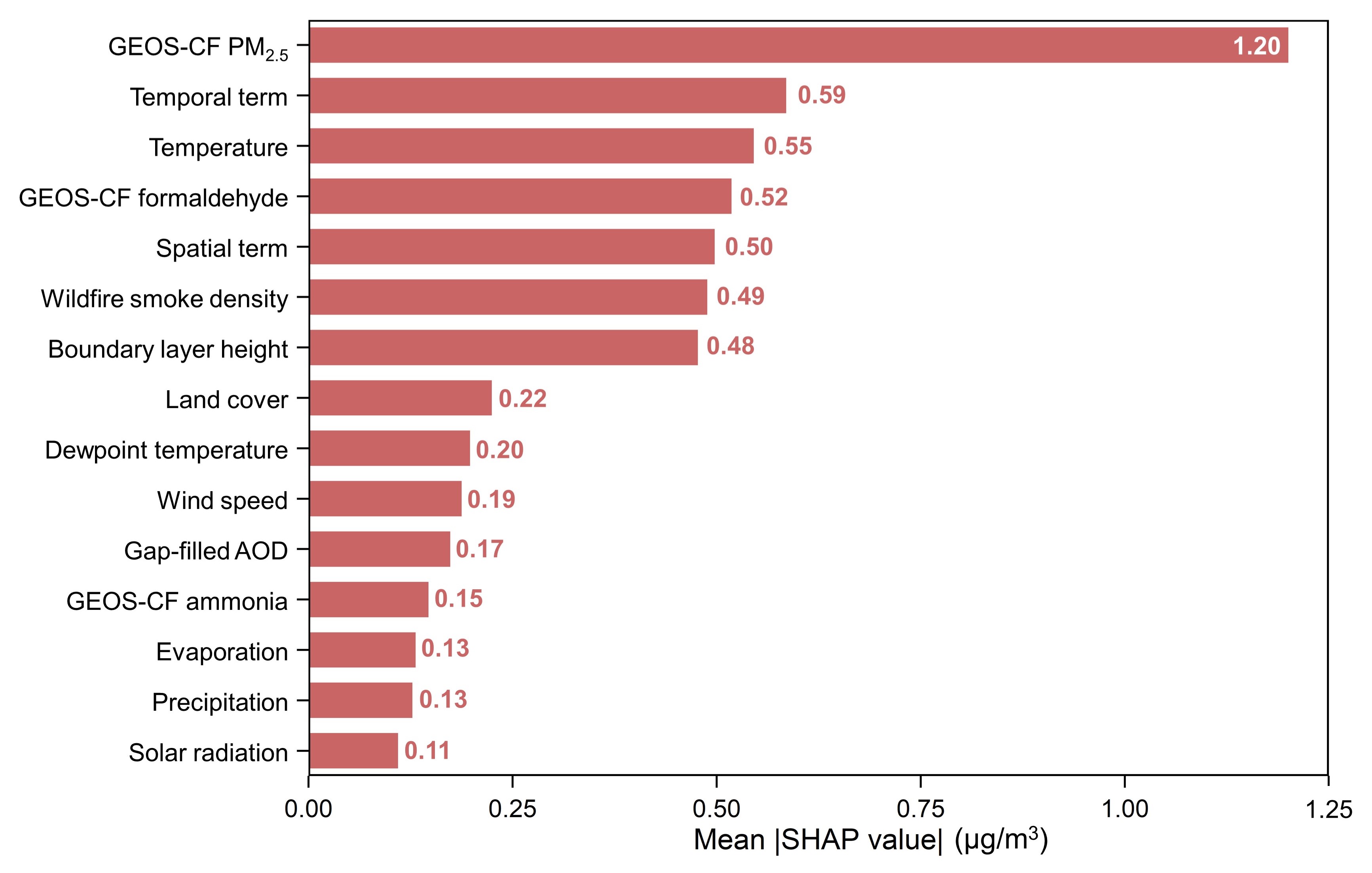

**Note:** Temporal term includes three temporal features;

Spatial term includes three spatial features;

Land cover includes 16 land cover features;

Wind speed includes 2 features in both directions.

**Figure S3.** Partial dependence plots (PDPs) of selected features ranking among the highest according to their mean absolute SHAP values.

| 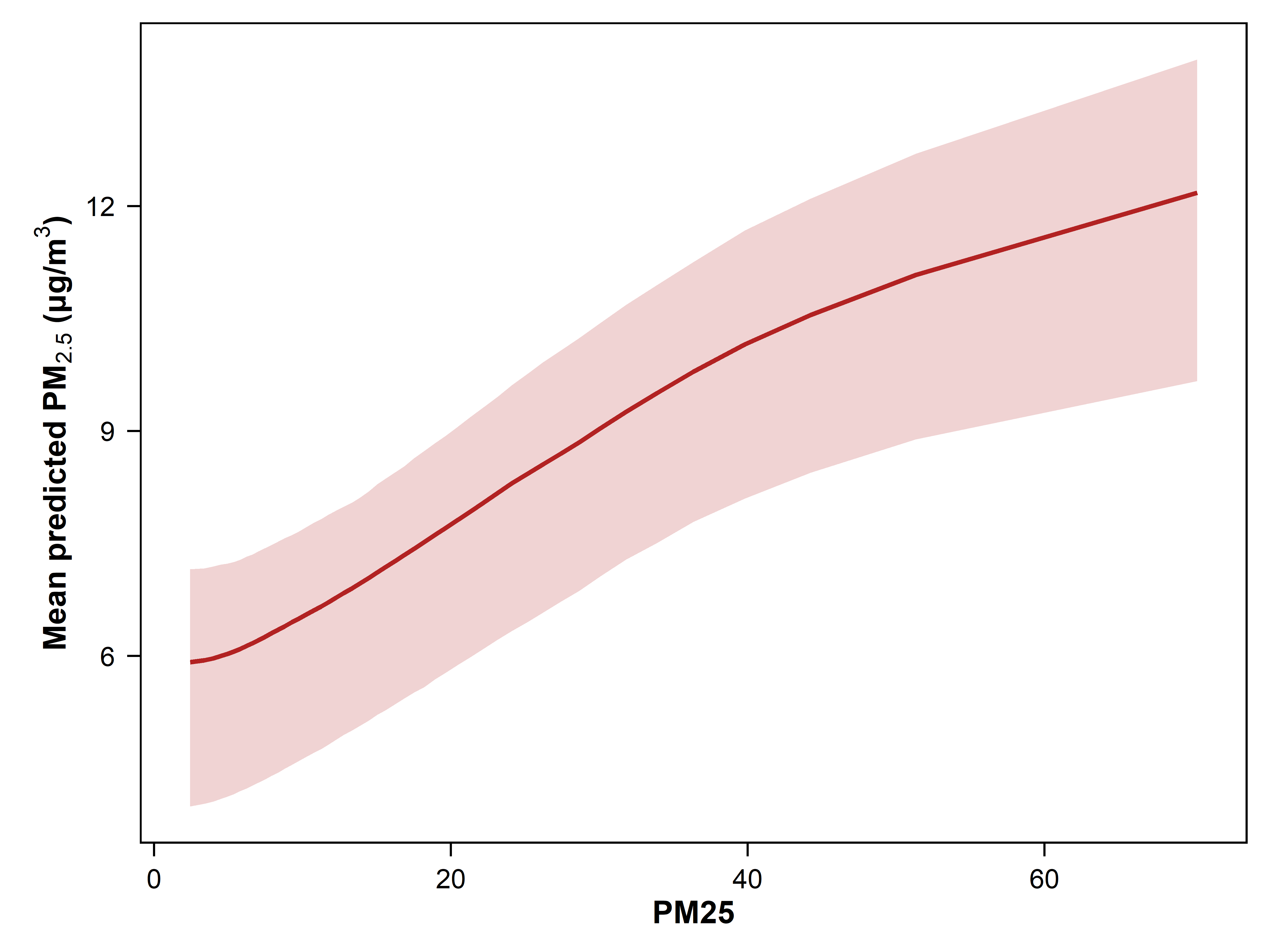 | 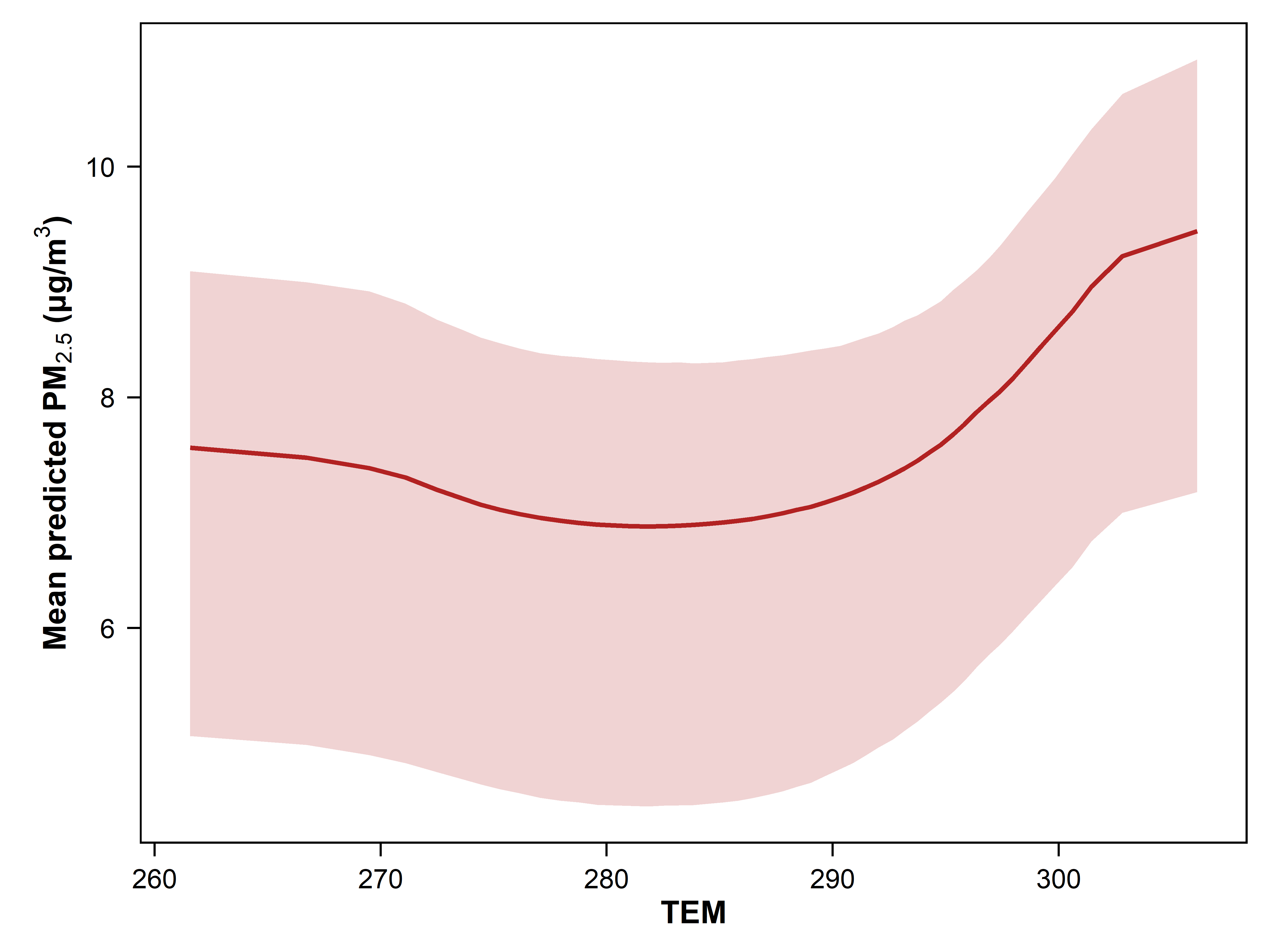 |
| --- | --- |
| **(*a*)** GEOS-CF simulated PM_2.5_ | **(*b*)** Temperature |
| 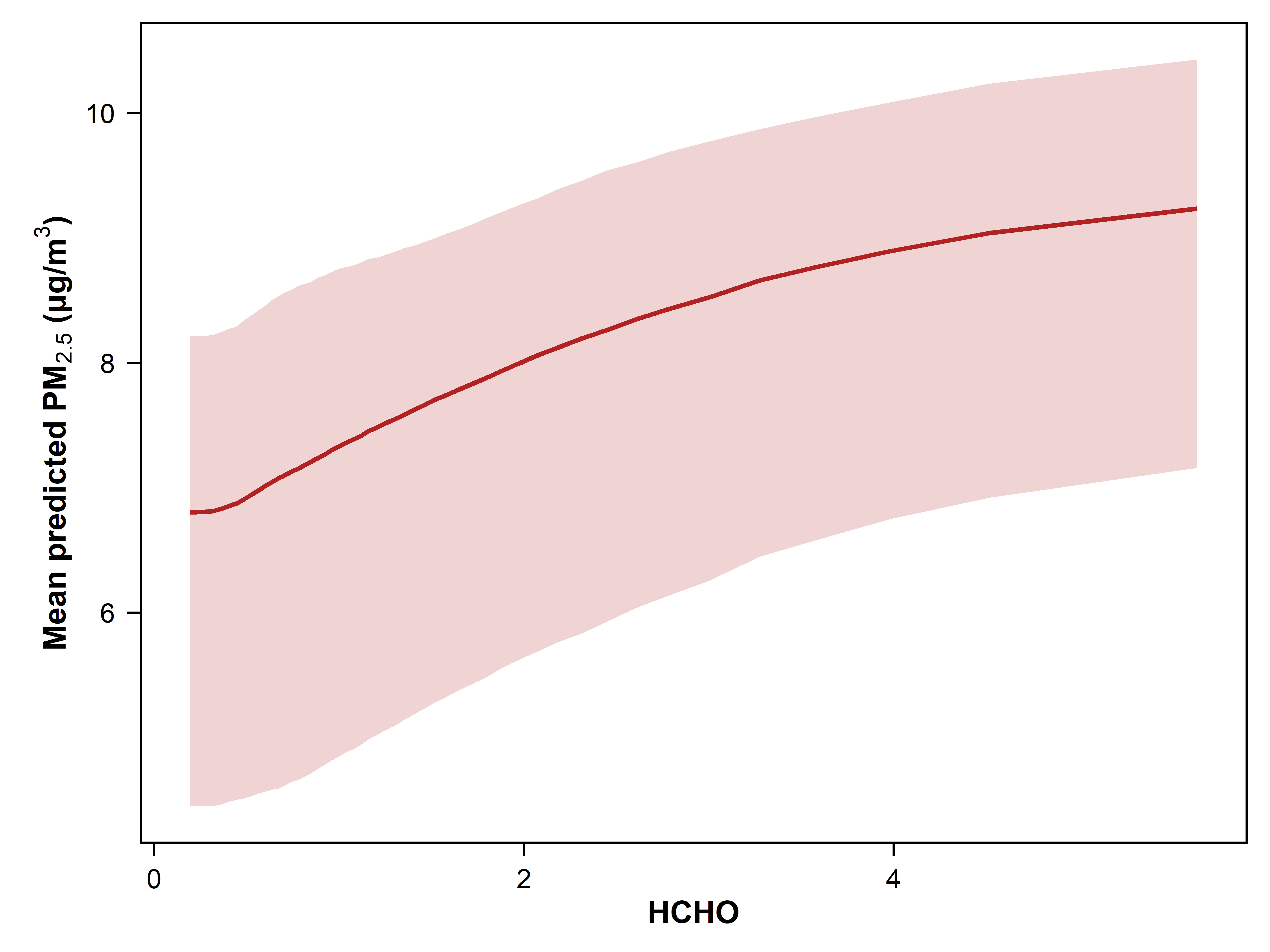 | 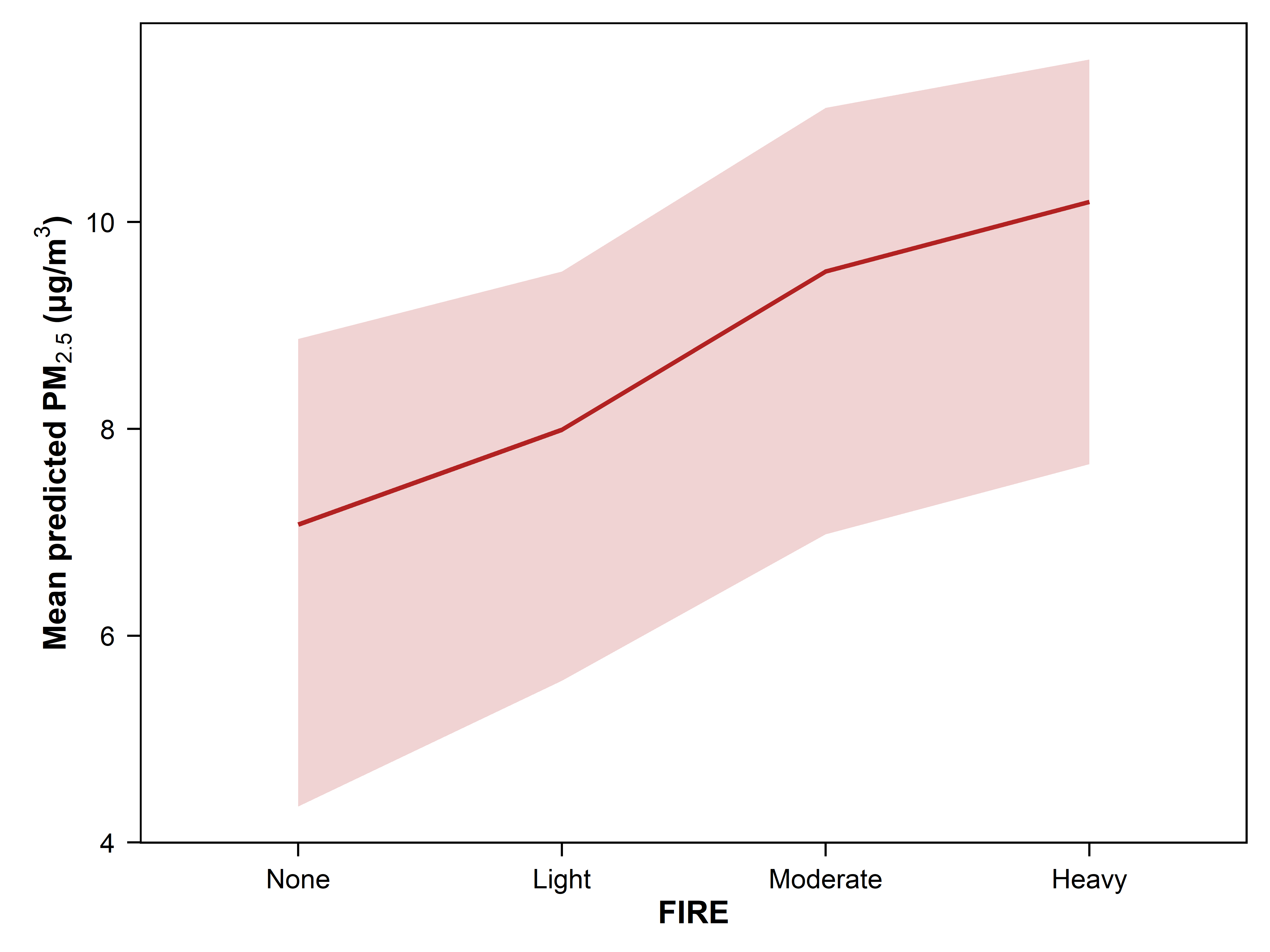 |
| **(*c*)** GEOS-CF simulated formaldehyde | **(*d*)** Wildfire smoke density |
| 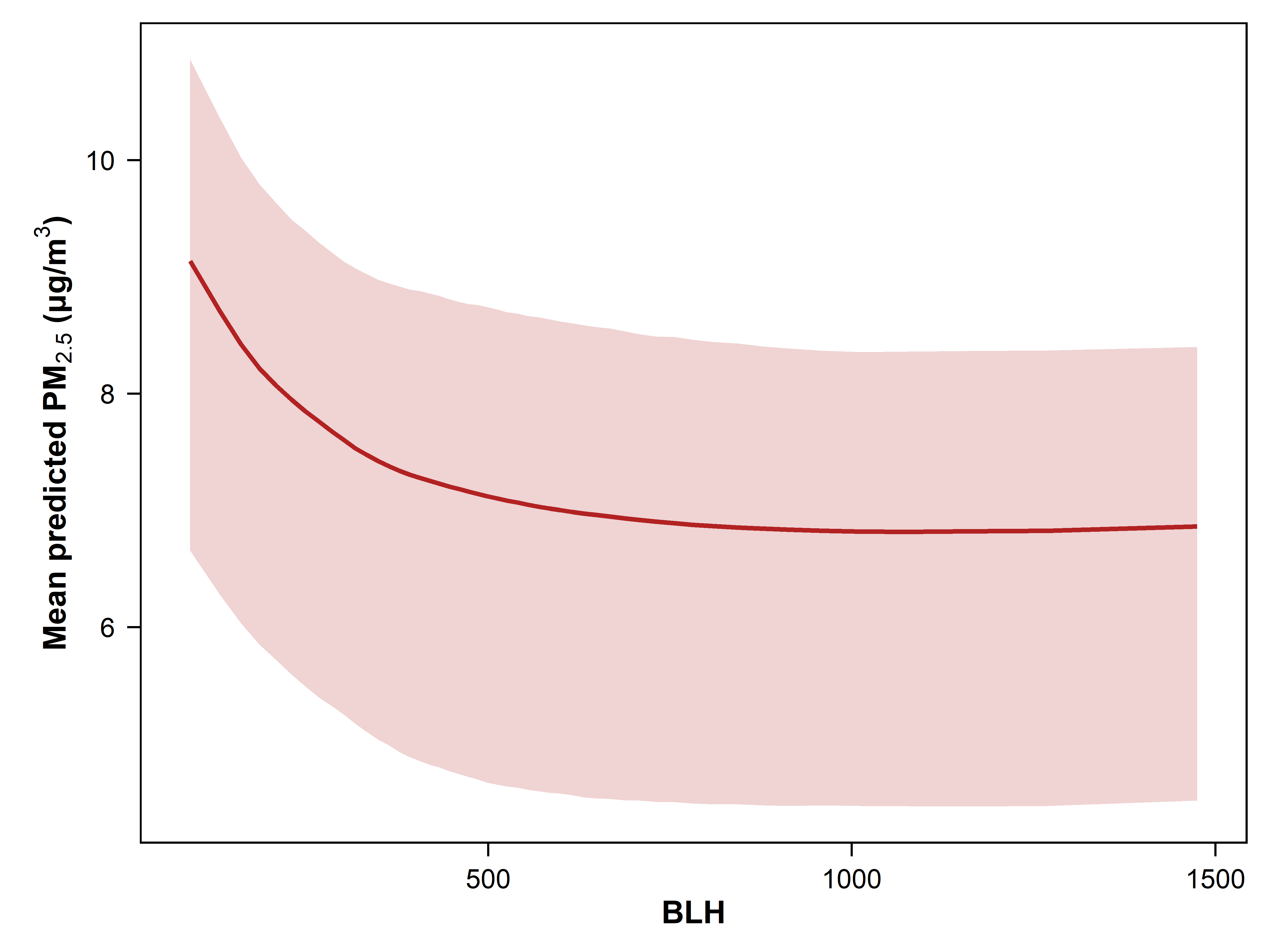 | 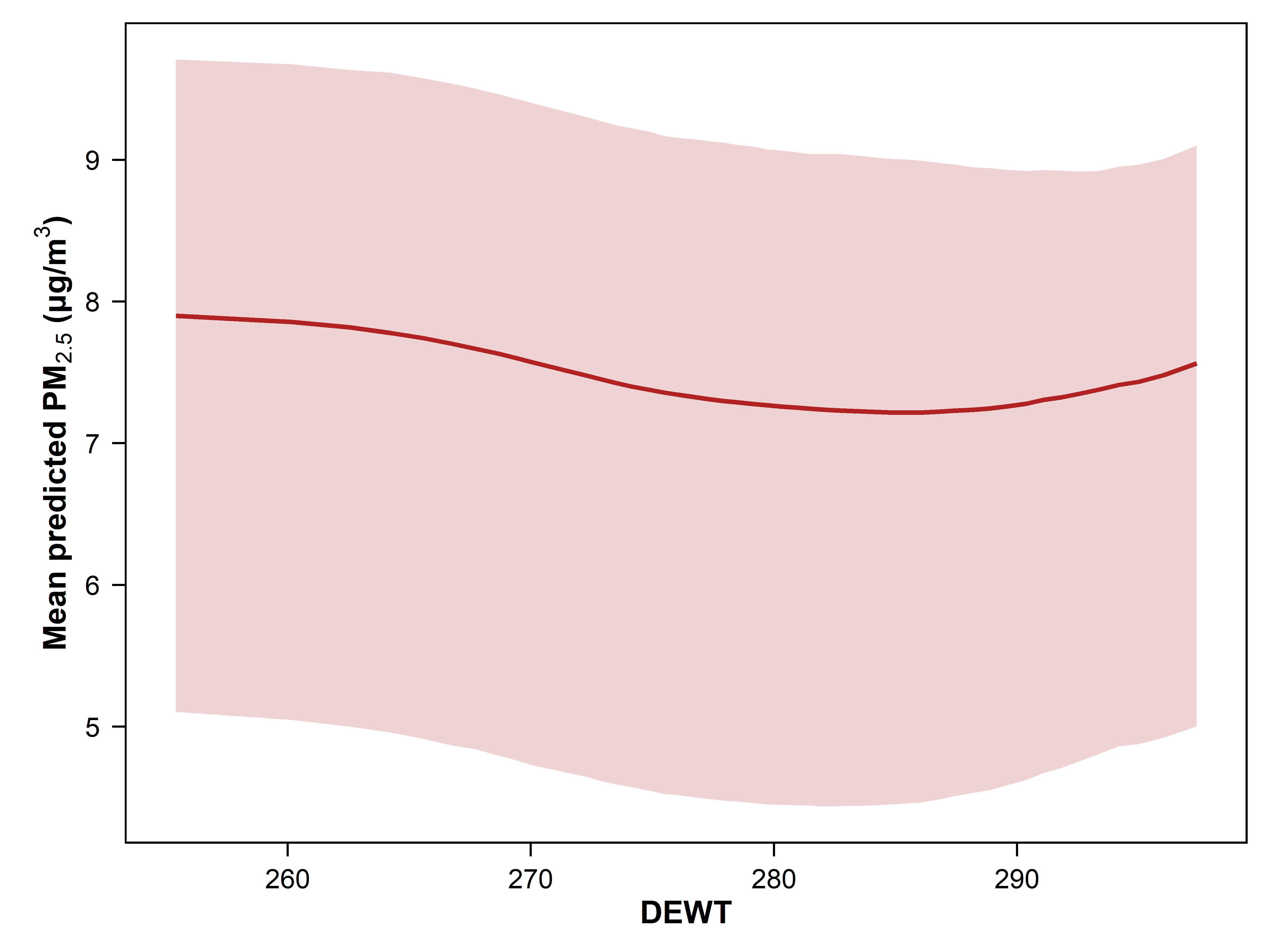 |
| **(*e*)** Boundary layer height | **(*f*)** Dewpoint temperature |
| 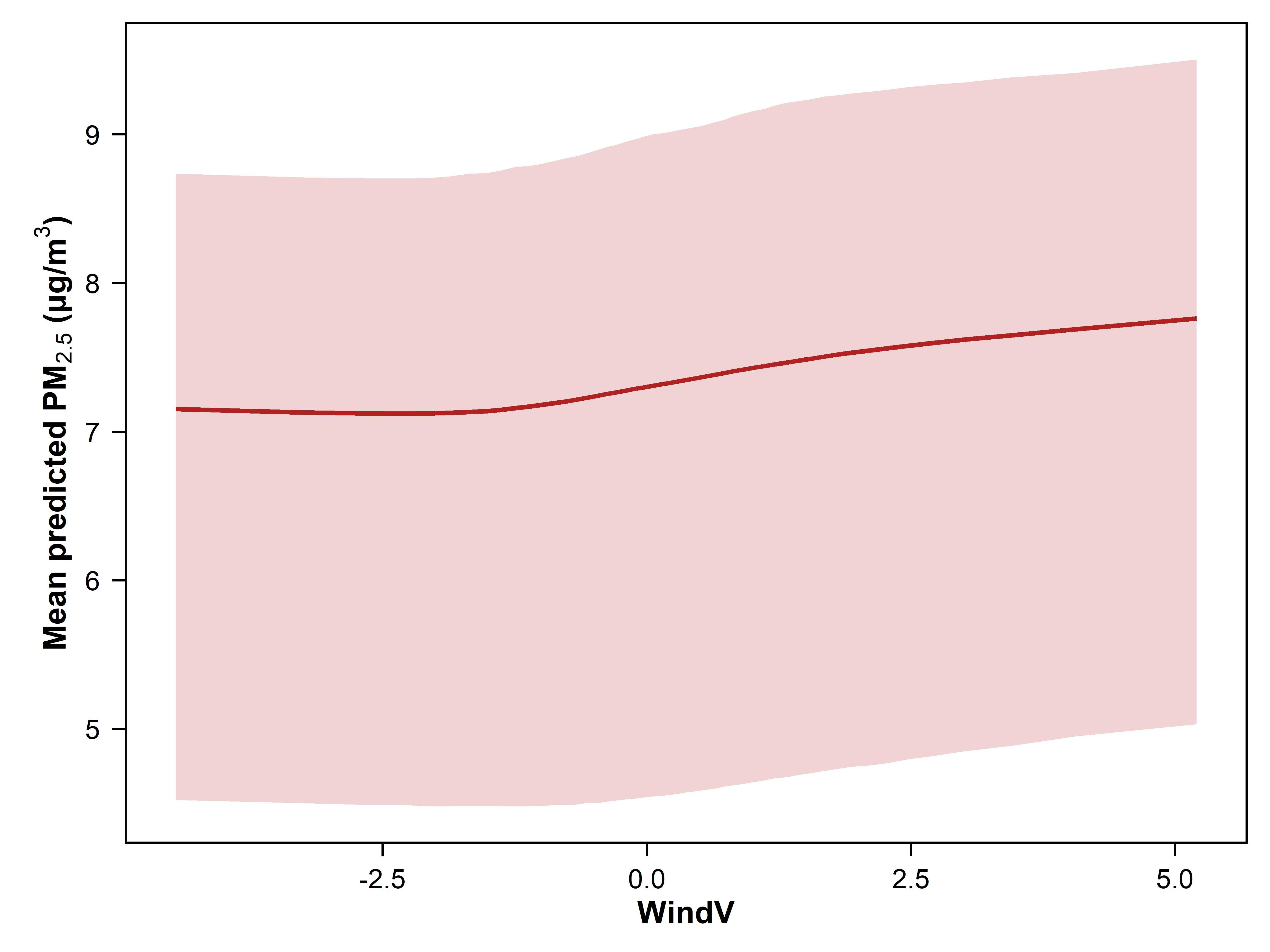 | 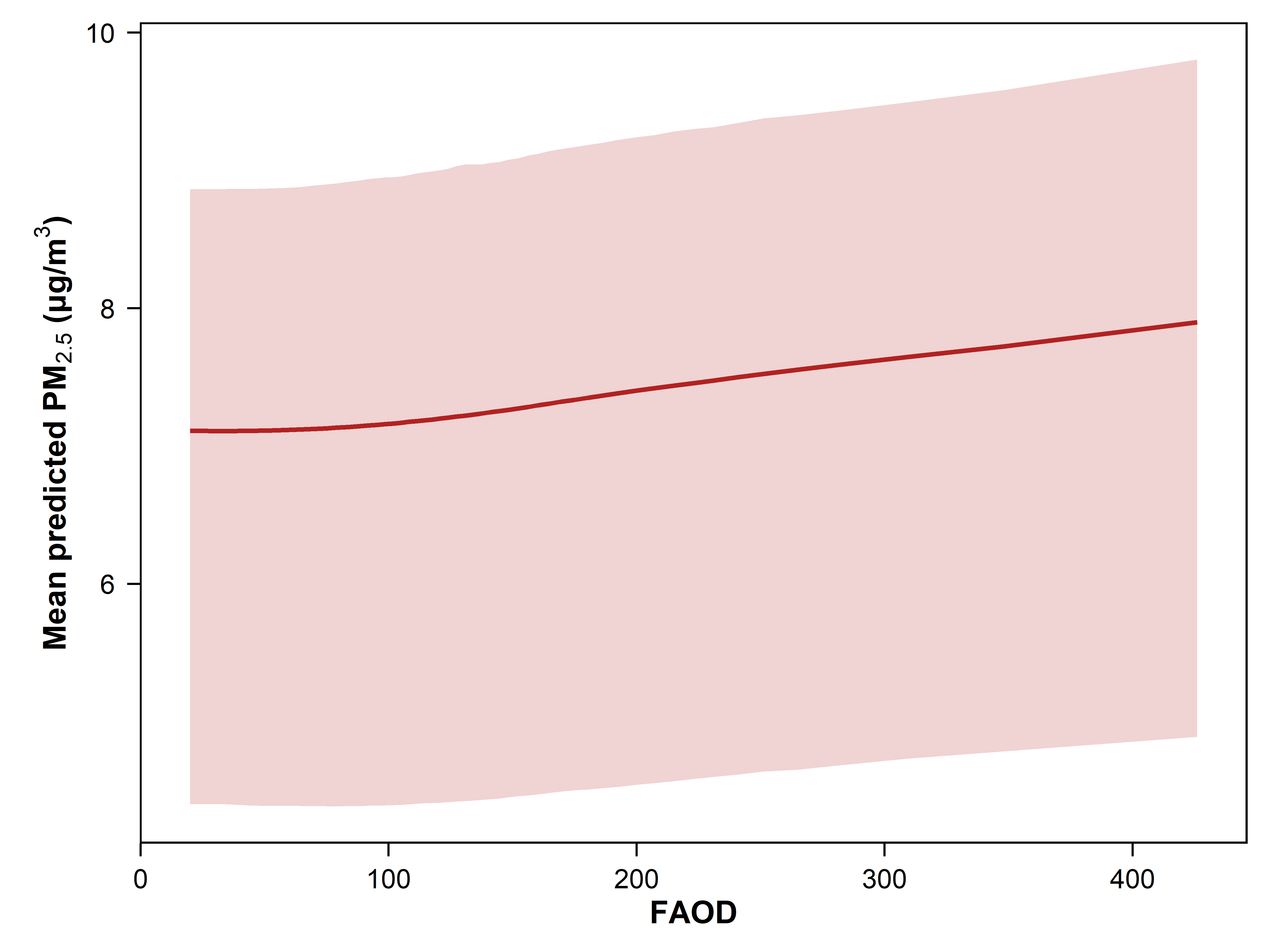 |
| **(*g*)** Northward wind speed | **(*h*)** Aerosol optical depth (AOD) |
| 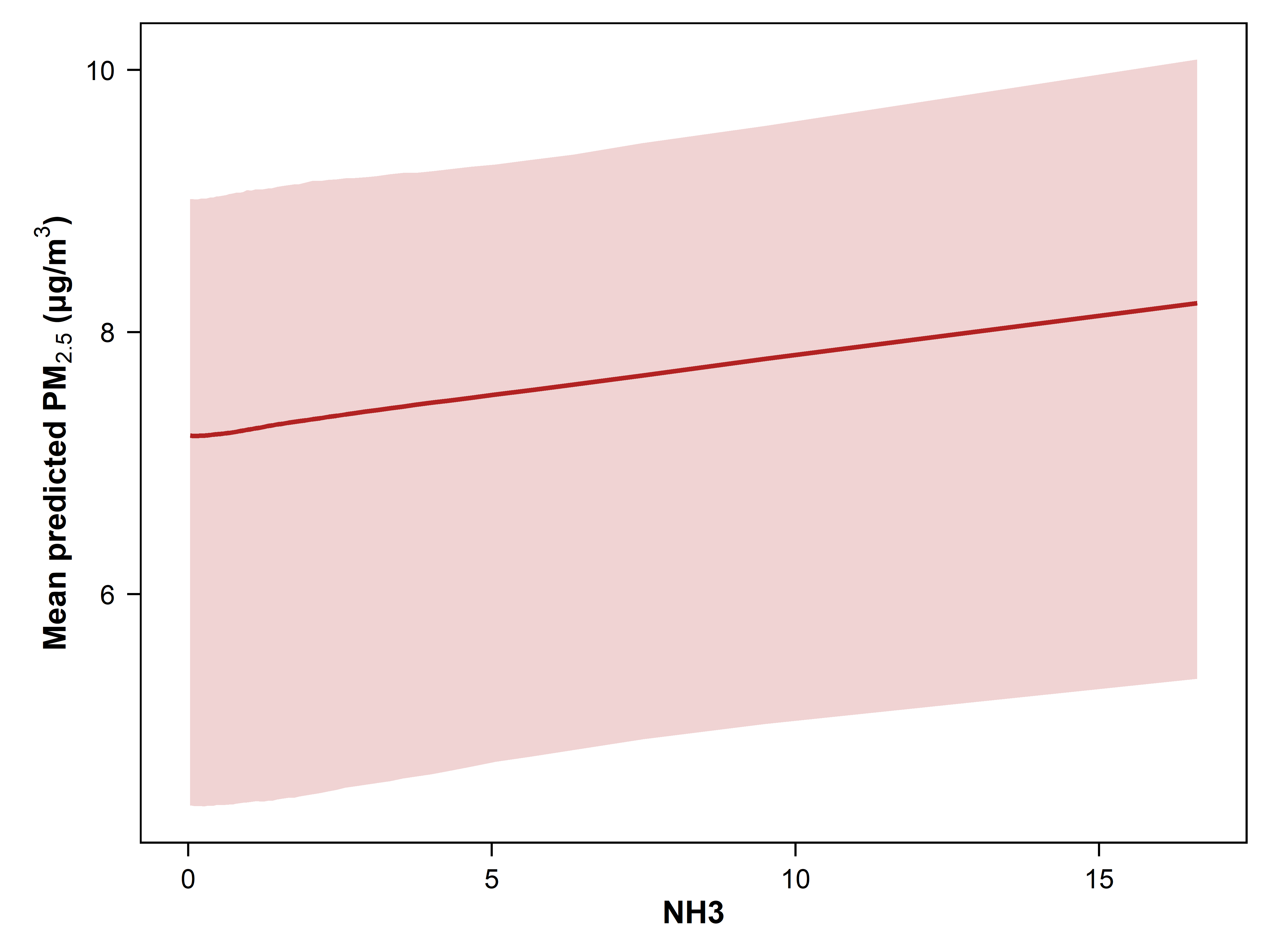 |  |
| **(*i*)** GEOS-CF simulated ammonia |  |

**Figure S4.** **Model performance of wildfire PM_2.5_ estimates in the 10-fold station-based cross-validation (CV):** (*a*) Daily average wildfire PM_2.5_ concentrations during smoke days; (*b*) Annual average wildfire PM_2.5_ concentrations across all days; (*c*) Annual average percentages of wildfire smoke to total PM_2.5_ across all days.

| 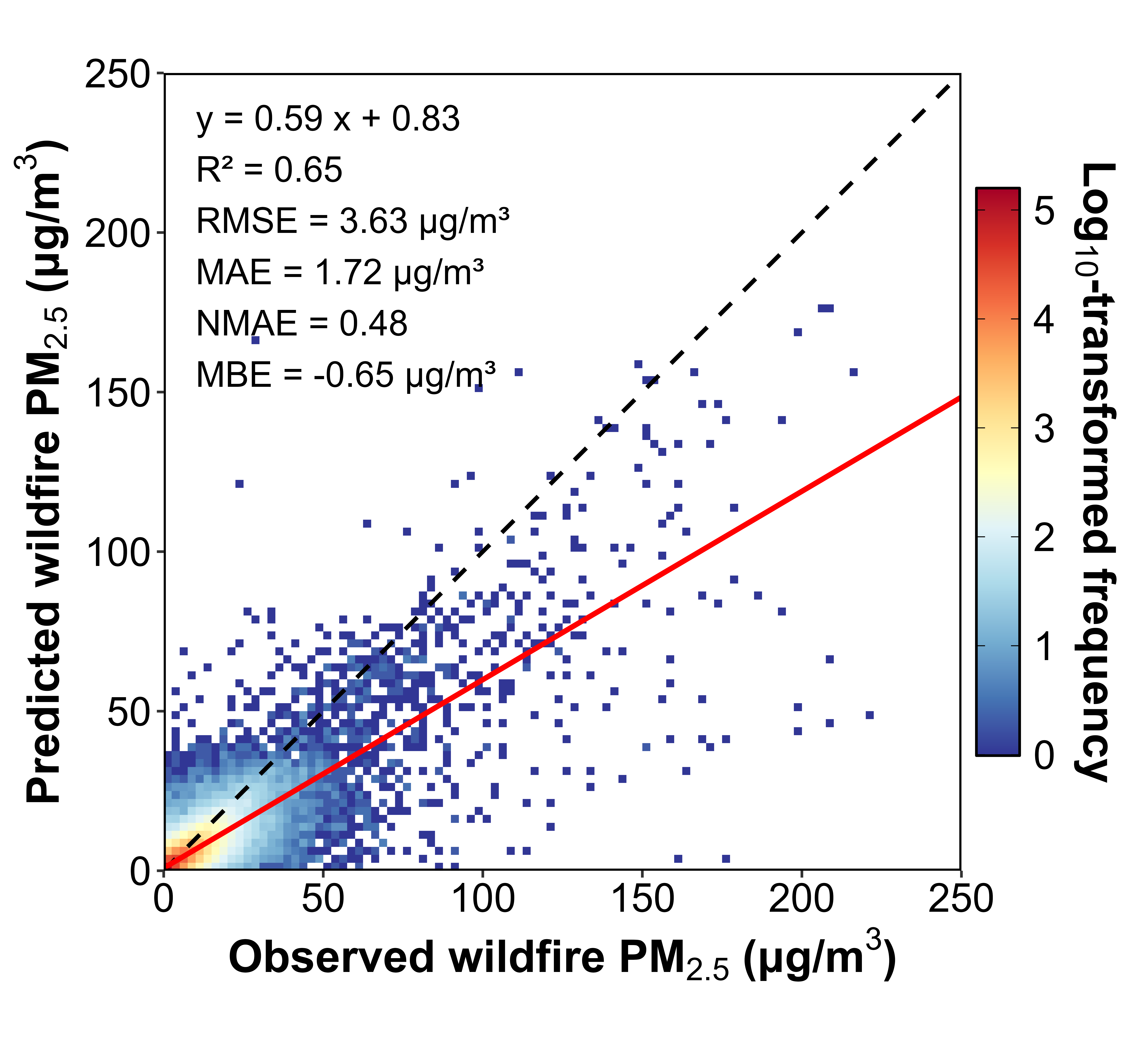 | |
| --- | --- |
| (*a*) | |
| 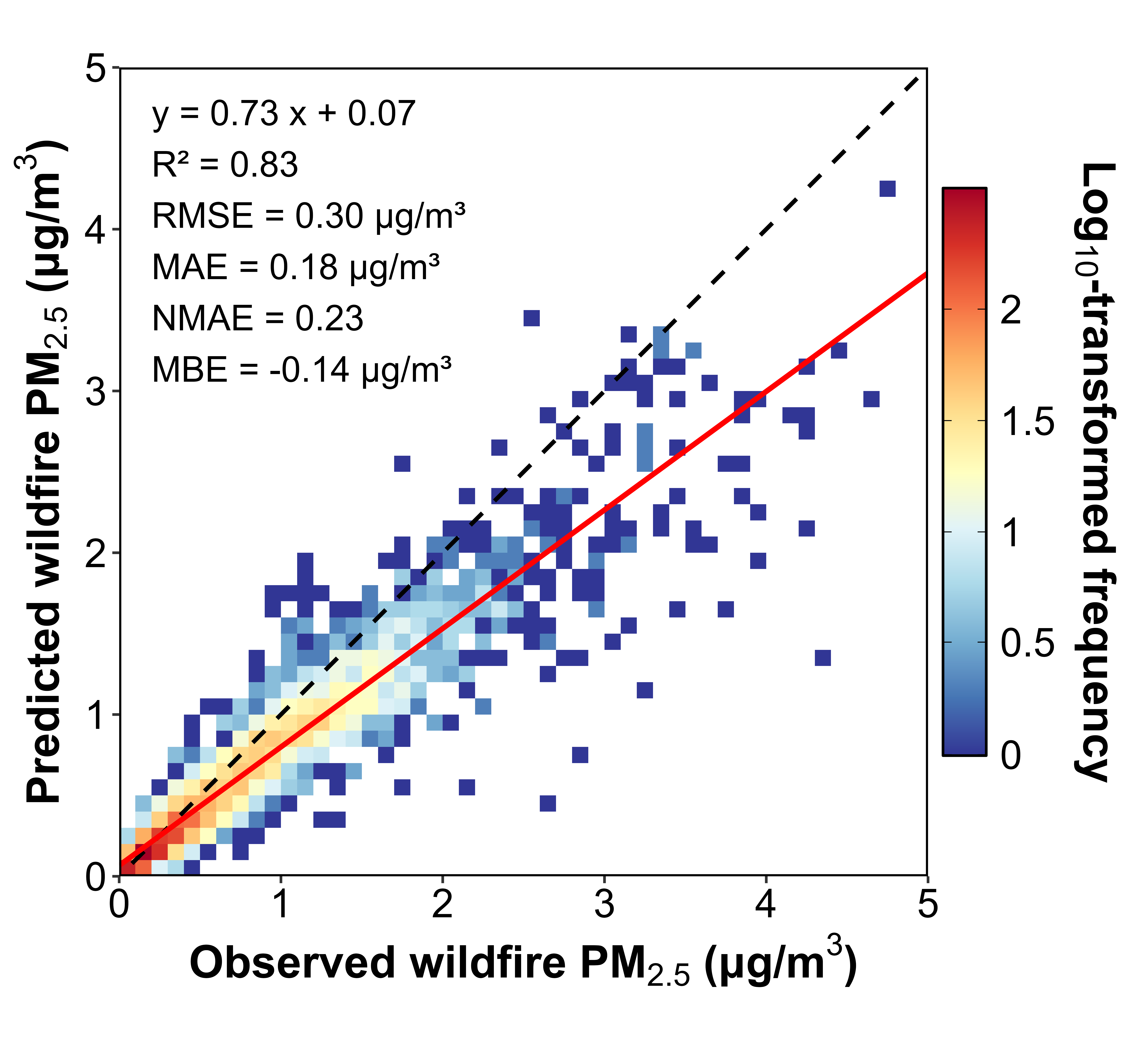 | 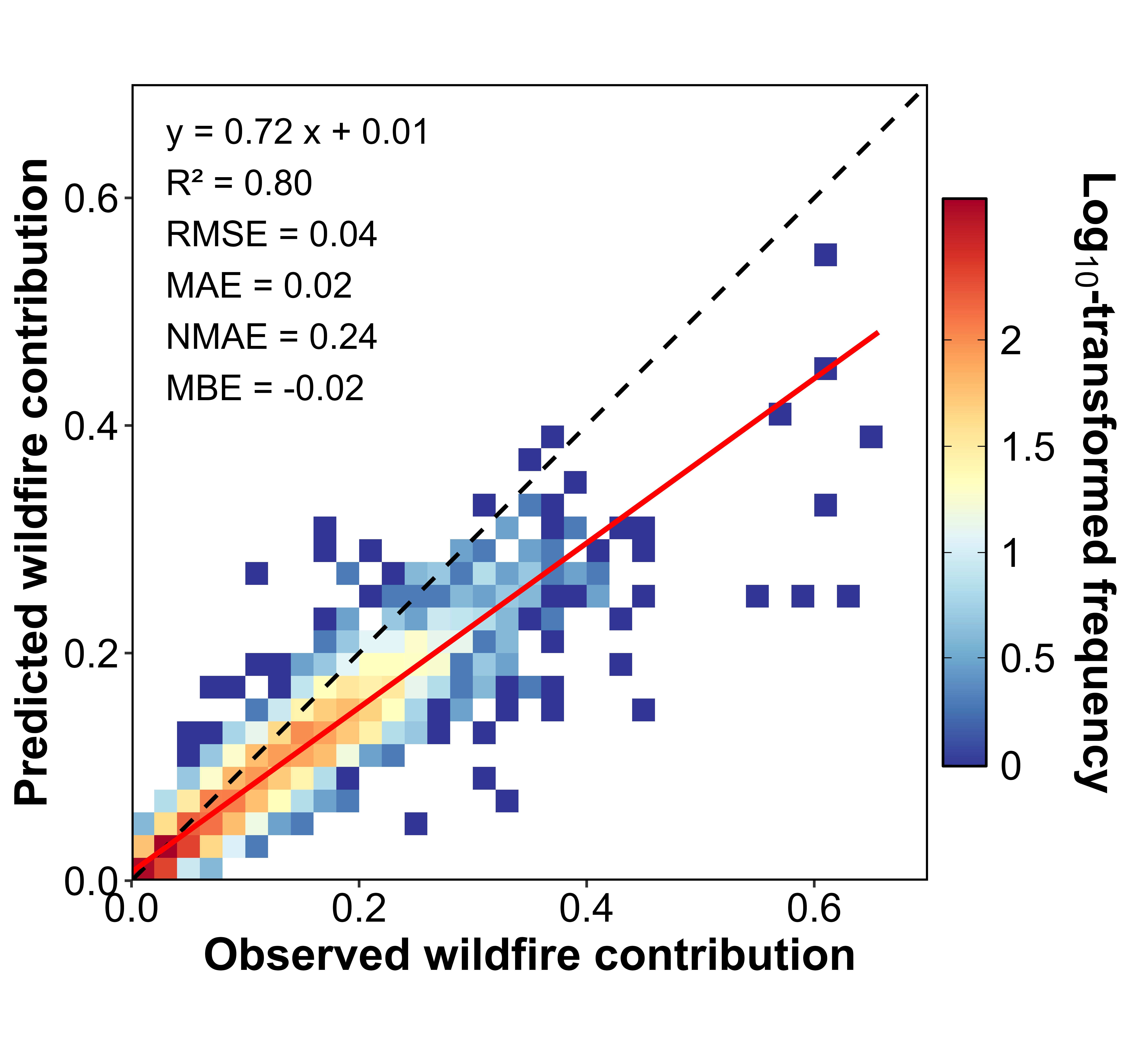 |
| (*b*) | (*c*) |

**Figure S5.** Nine geographic divisions across the Unites States.

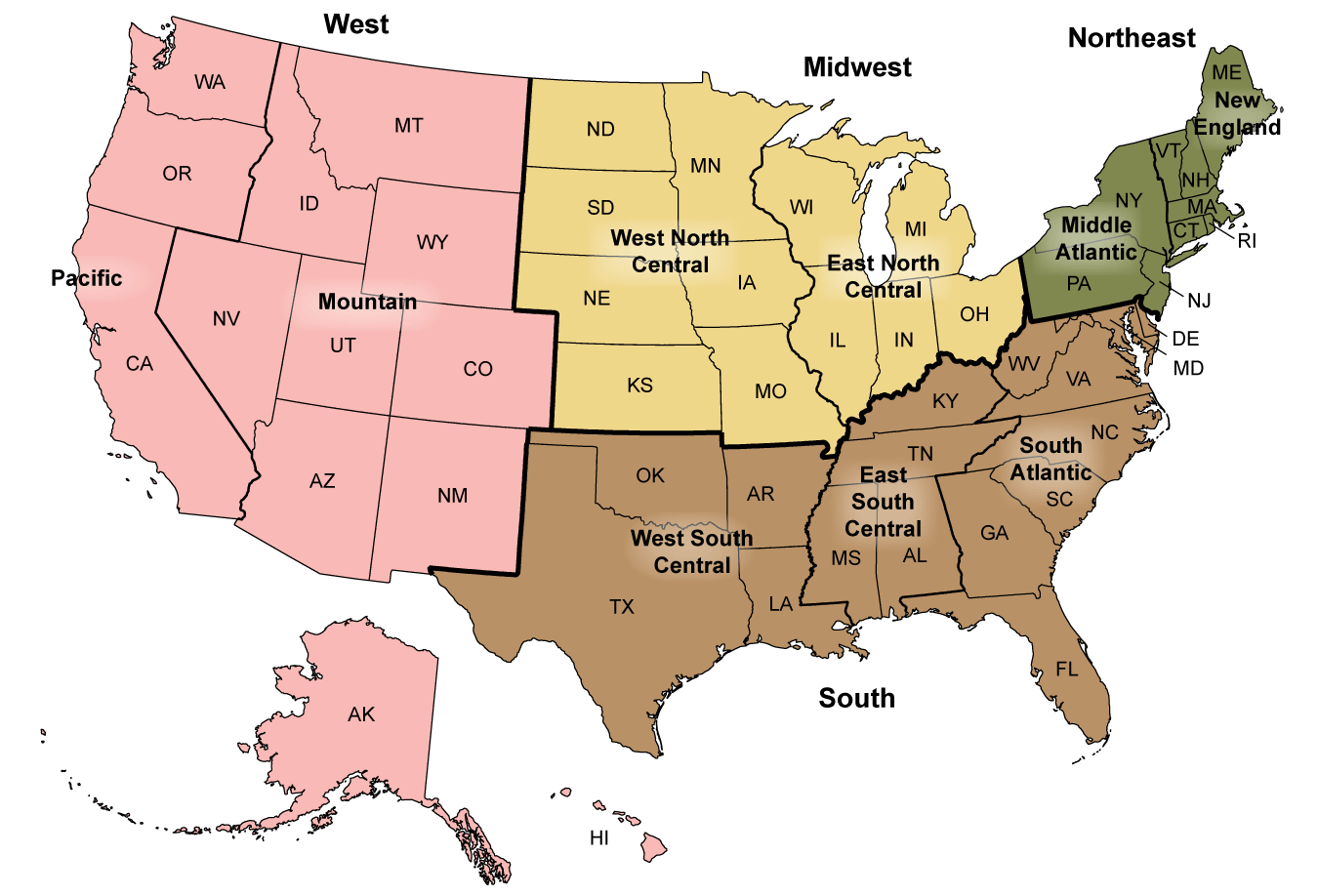

Source: <https://www.cdc.gov/nchs/hus/sources-definitions/geographic-region.htm>

**Figure S6.** Annual average 1-km estimate of wildfire PM_2.5_ between 2018 and 2024.

| 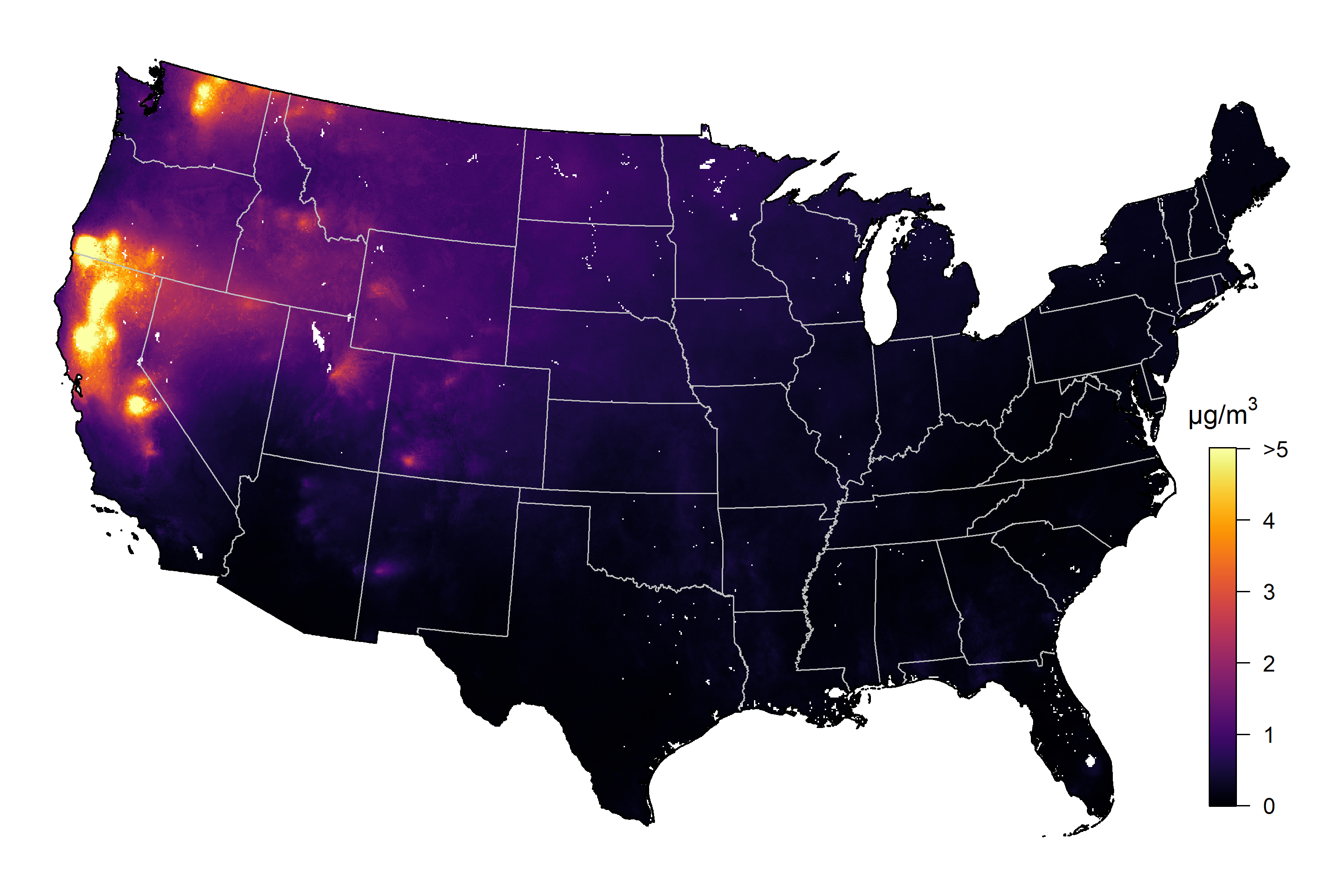 | 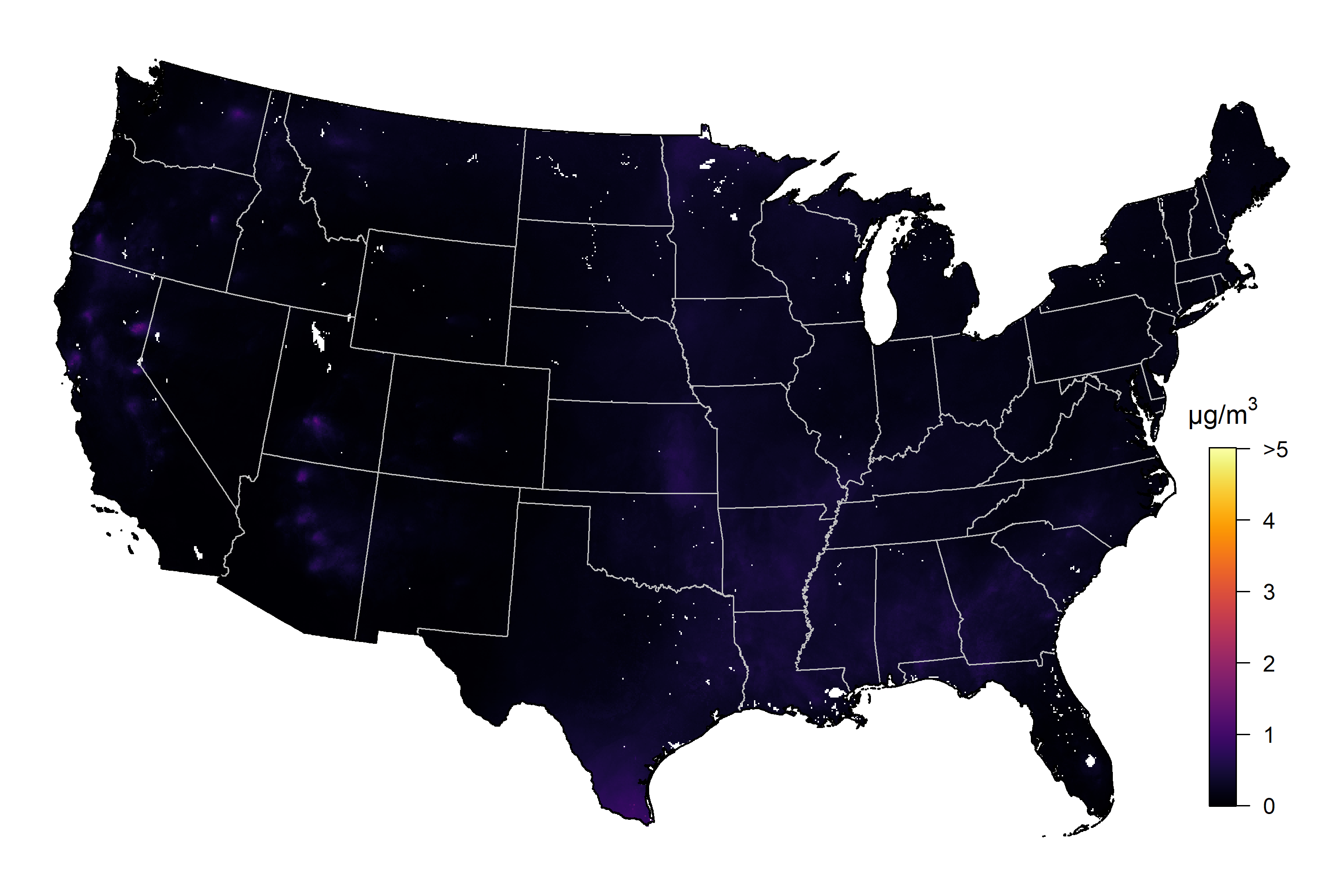 |
| --- | --- |
| (*a*) 2018 | (*b*) 2019 |
| 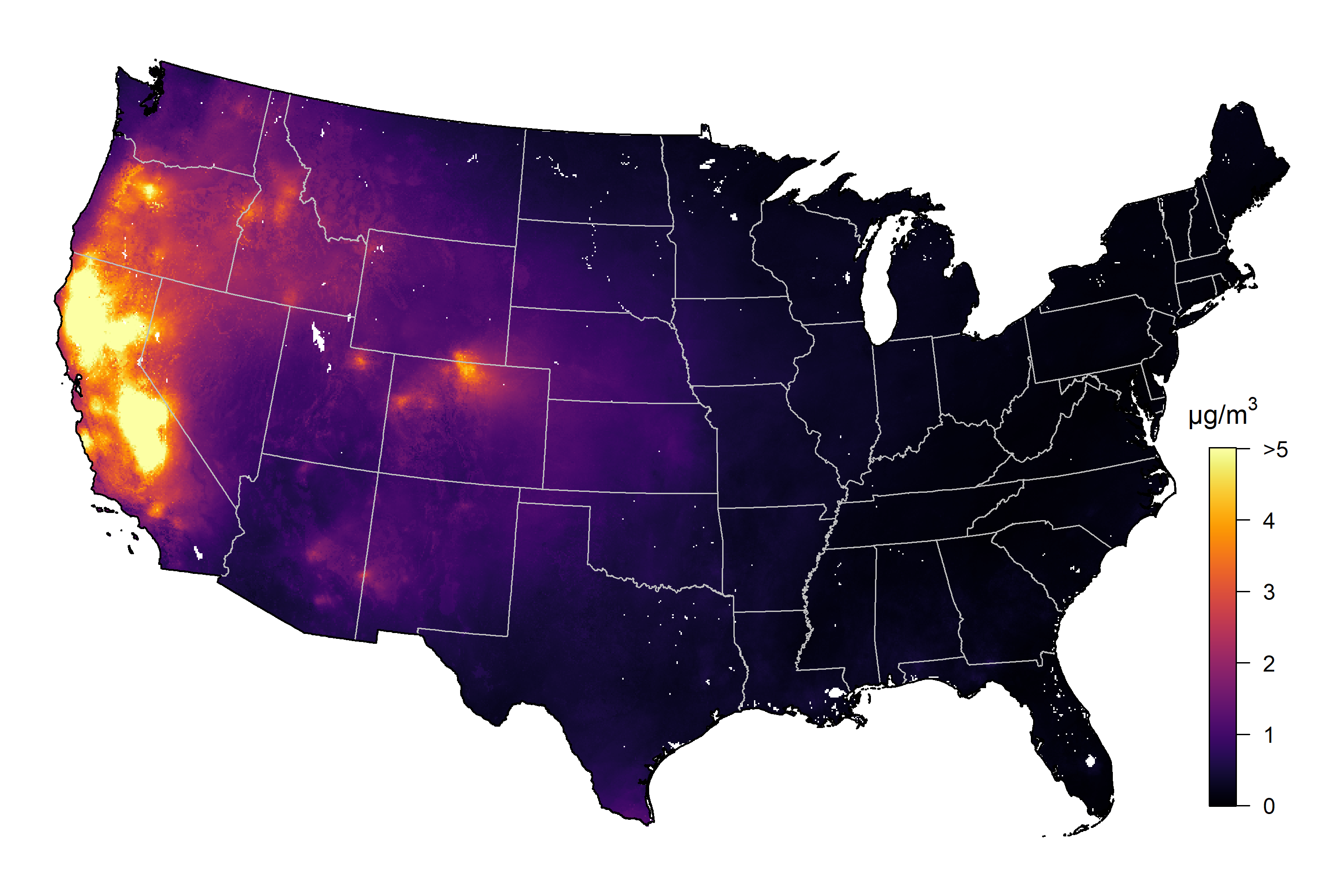 | 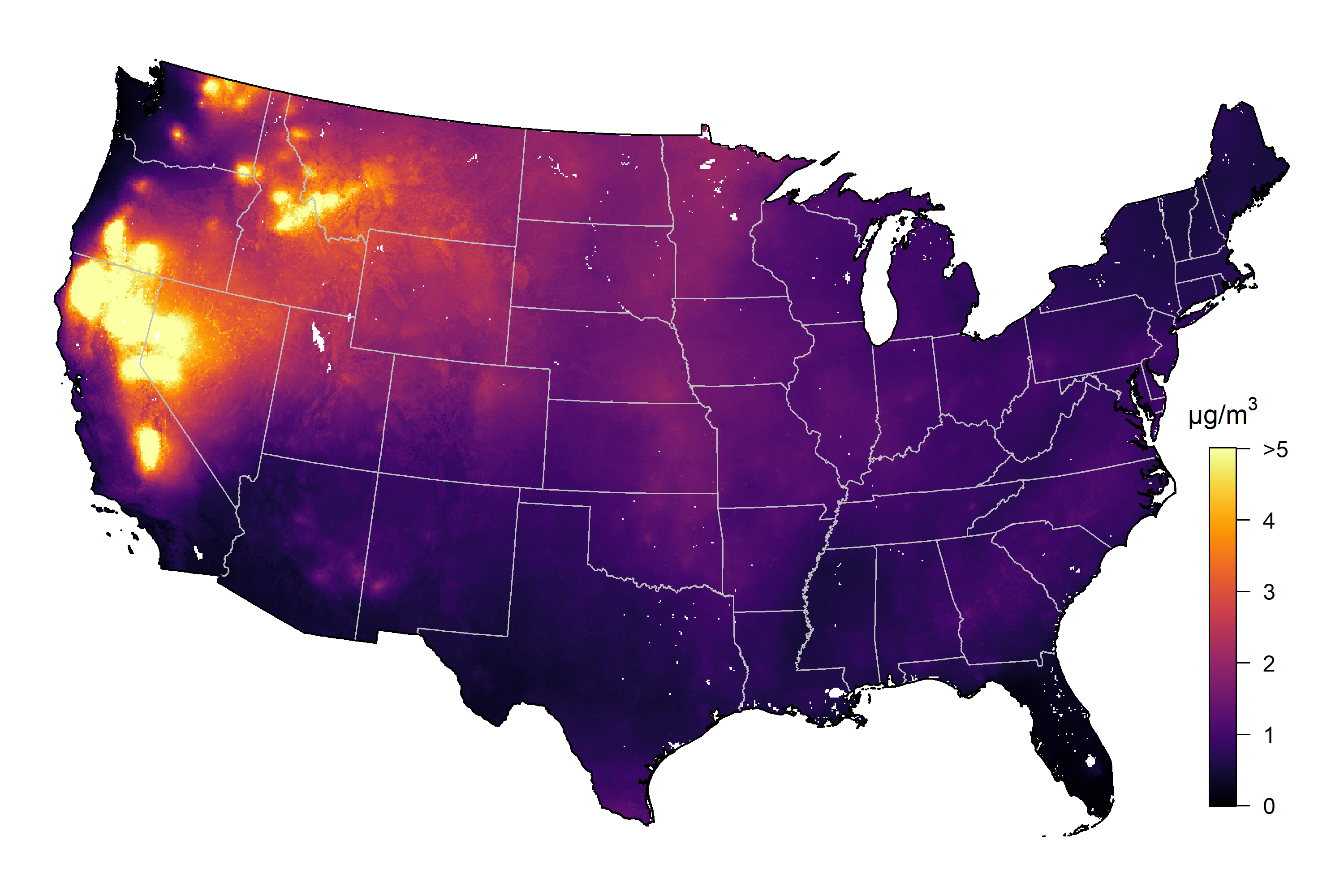 |
| (*c*) 2020 | (*d*) 2021 |
| 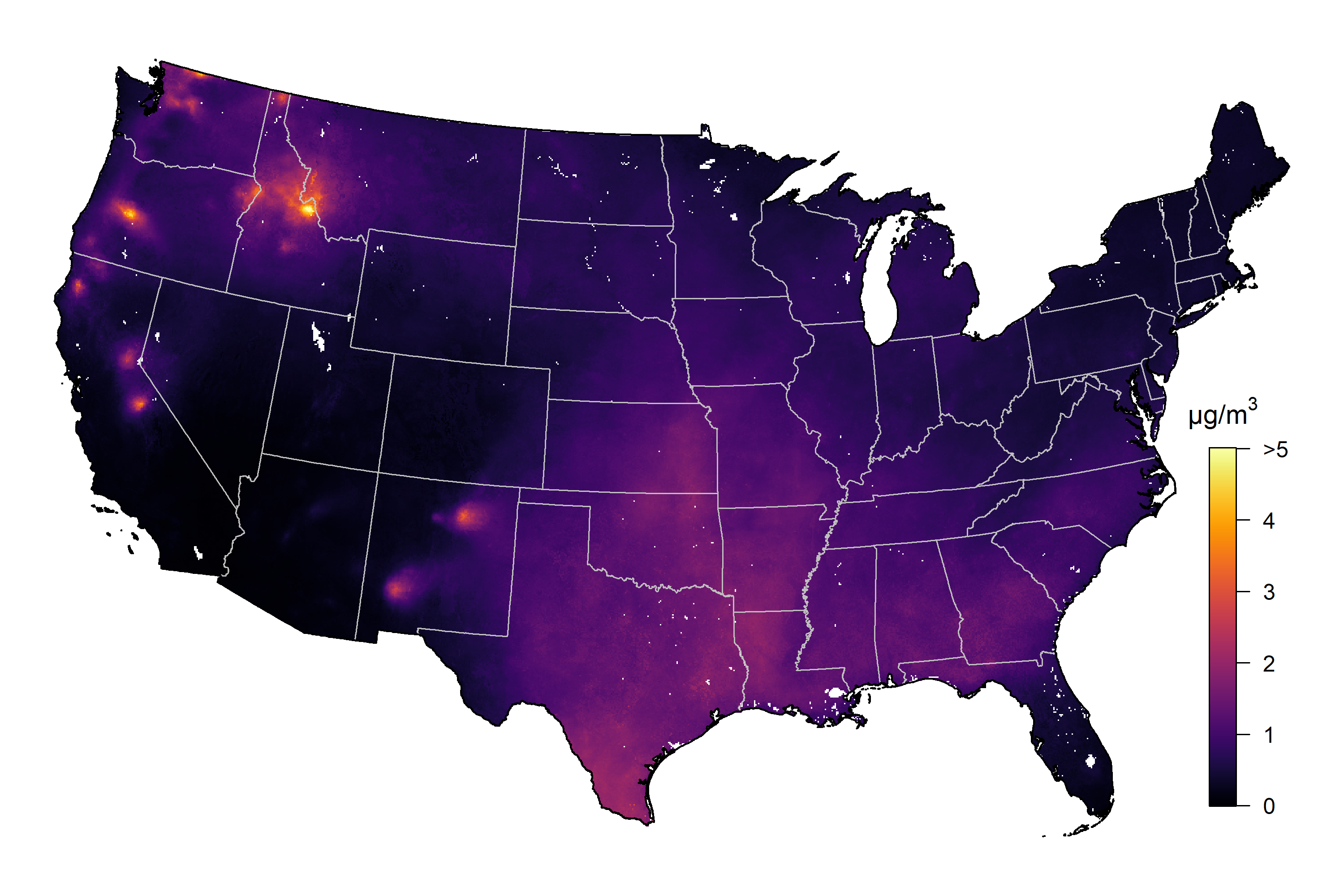 | 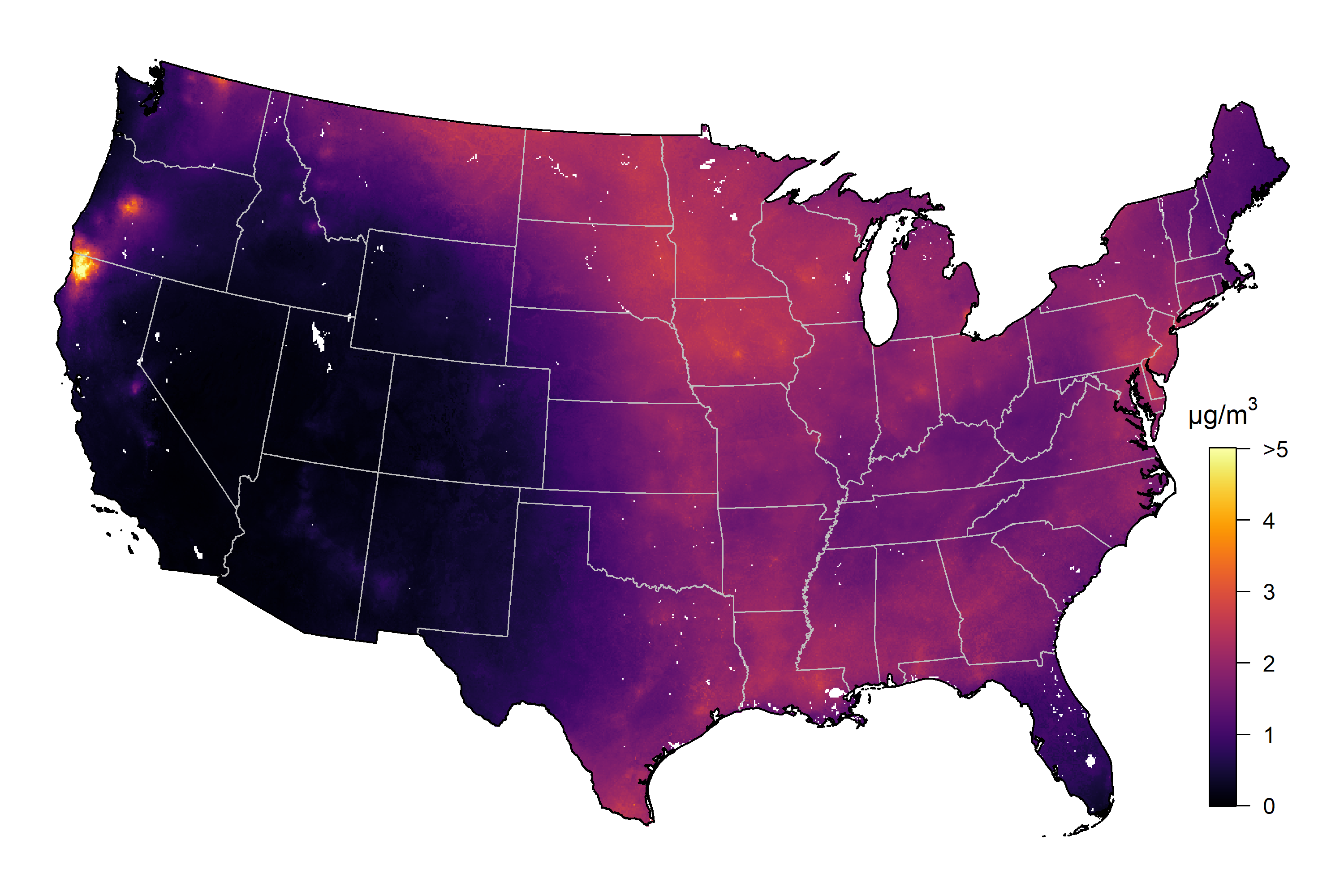 |
| (*e*) 2022 | (*f*) 2023 |
| 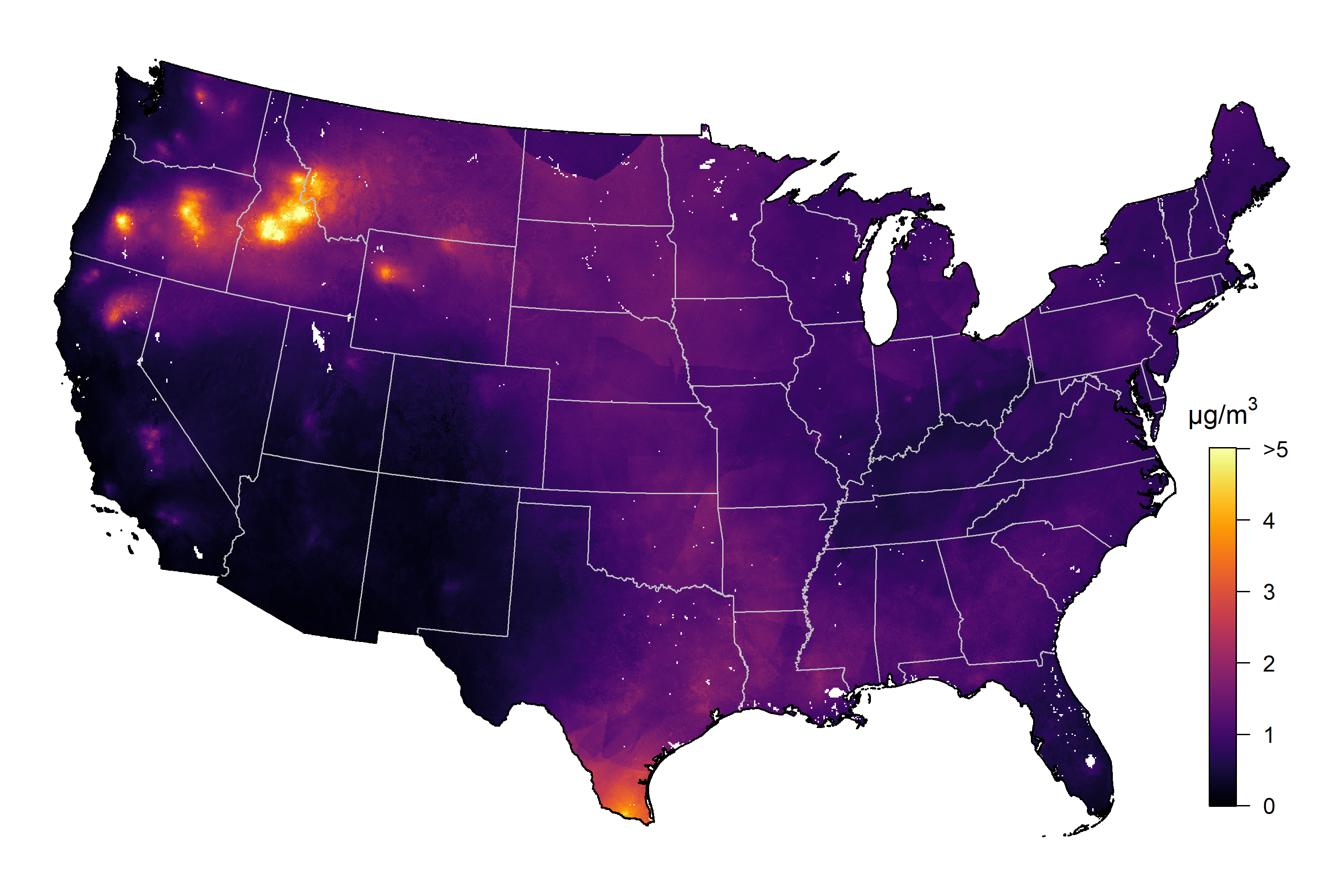 |  |
| (*g*) 2024 |  |

**Figure S7.** Seasonal average 1-km estimate of total PM_2.5_ between 2018 and 2024.

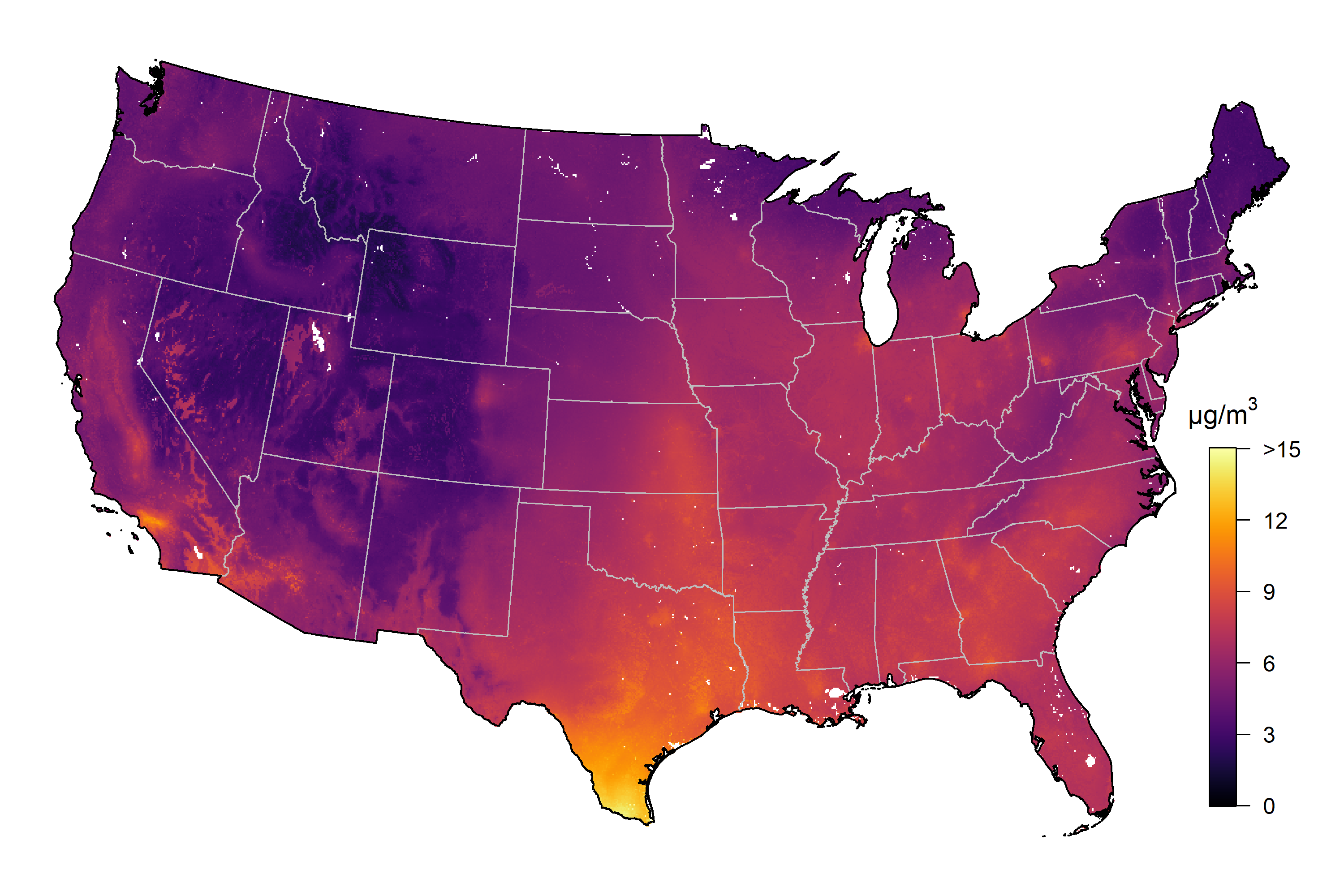

(a) Spring (March, April, May)

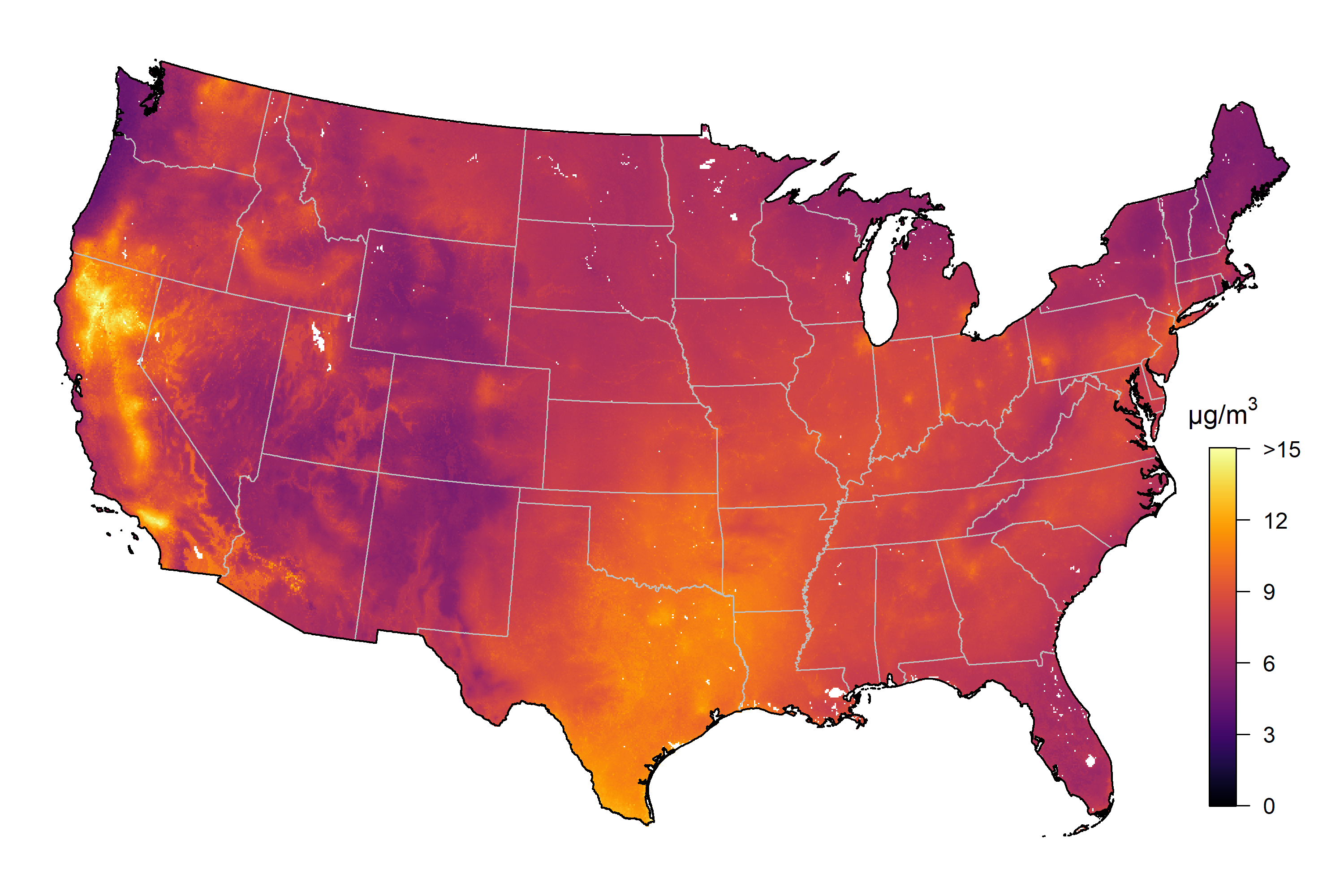

(b) Summer (June, July, August)

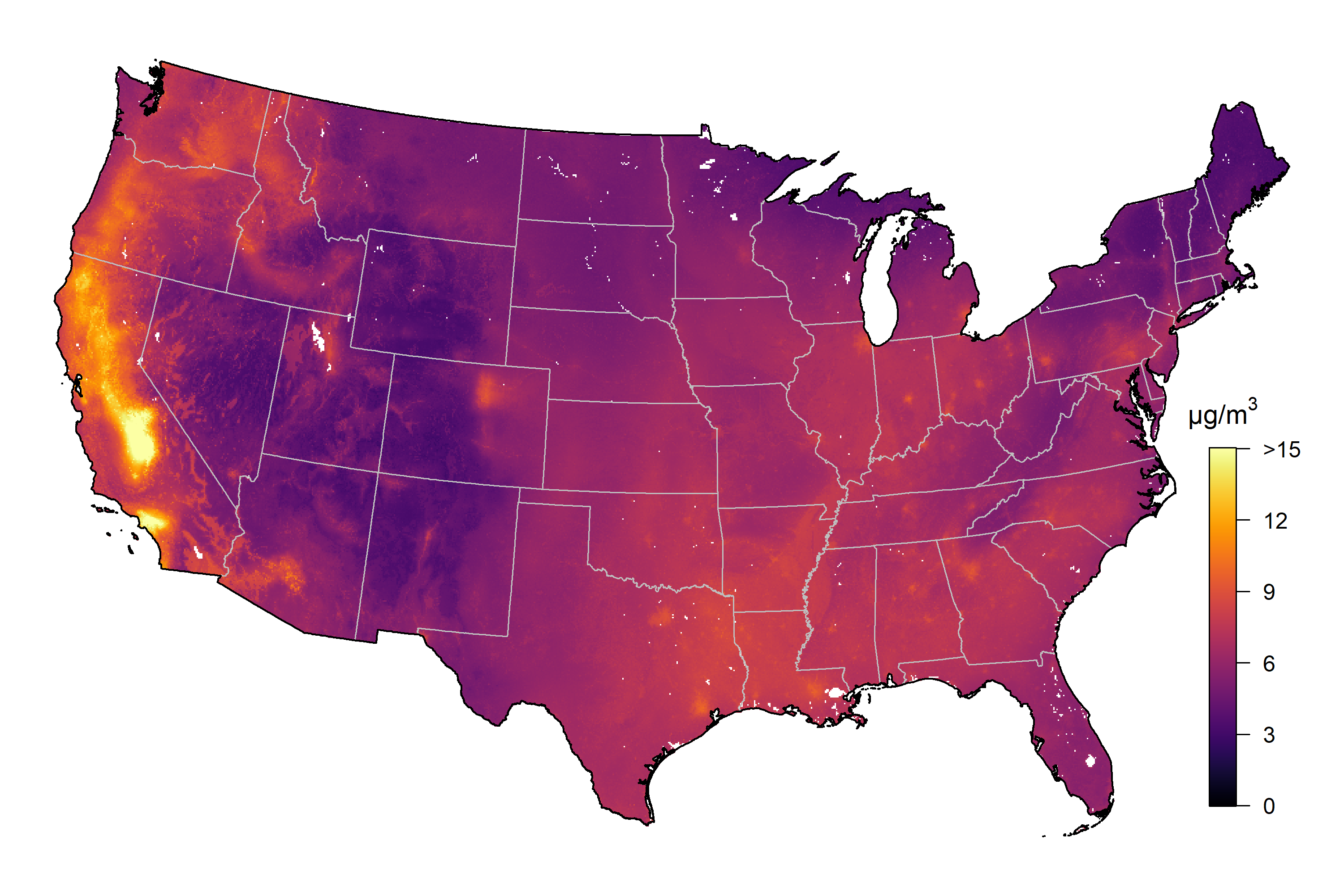

(c) Fall (September, October, November)

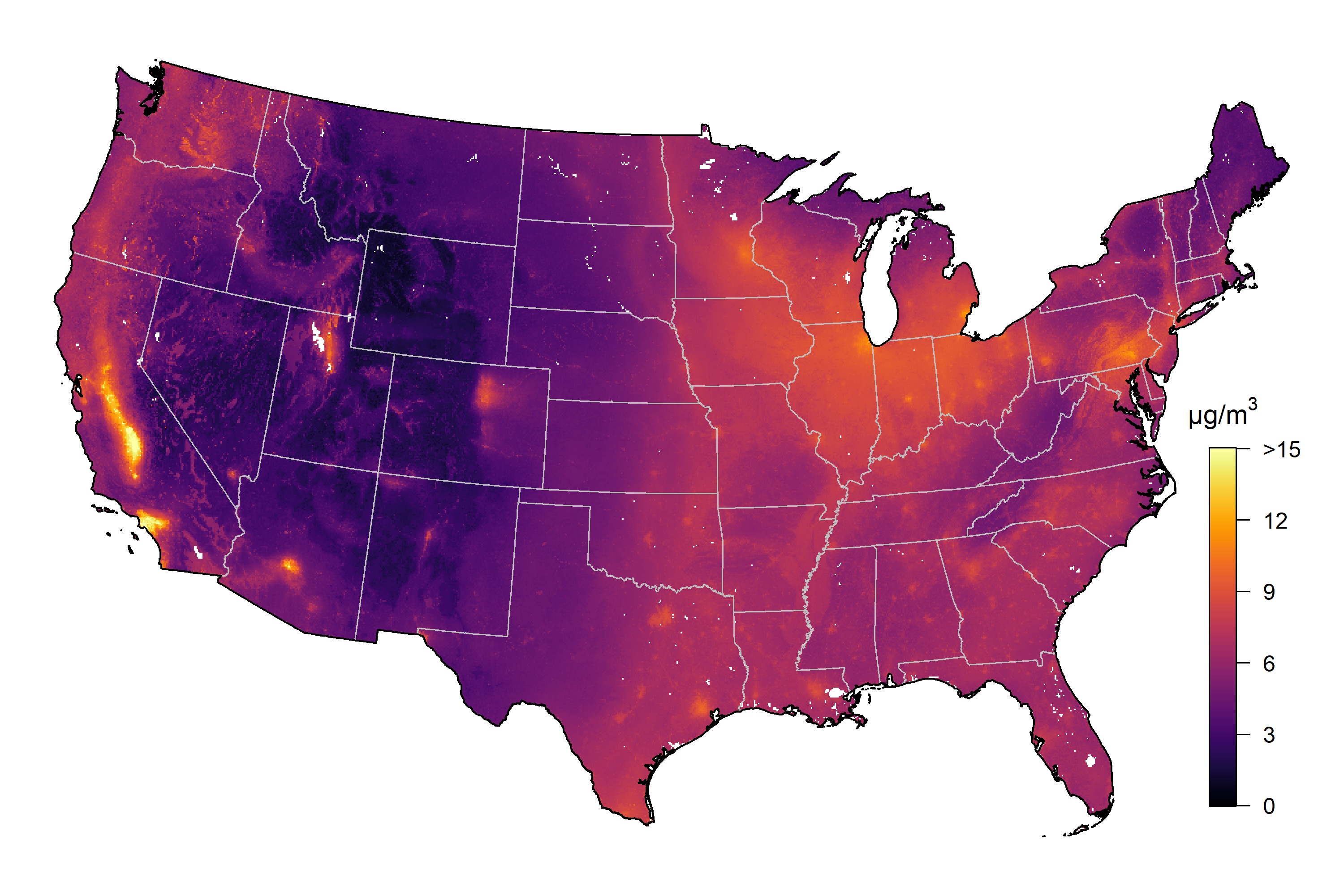

(d) Winter (December, January, February)

**Figure S8.** Comparing annual average PM_2.5_ concentration in 2021 (unit: μg/m^3^) at 1-km and 10-km resolution in eight major cities across the CONUS.

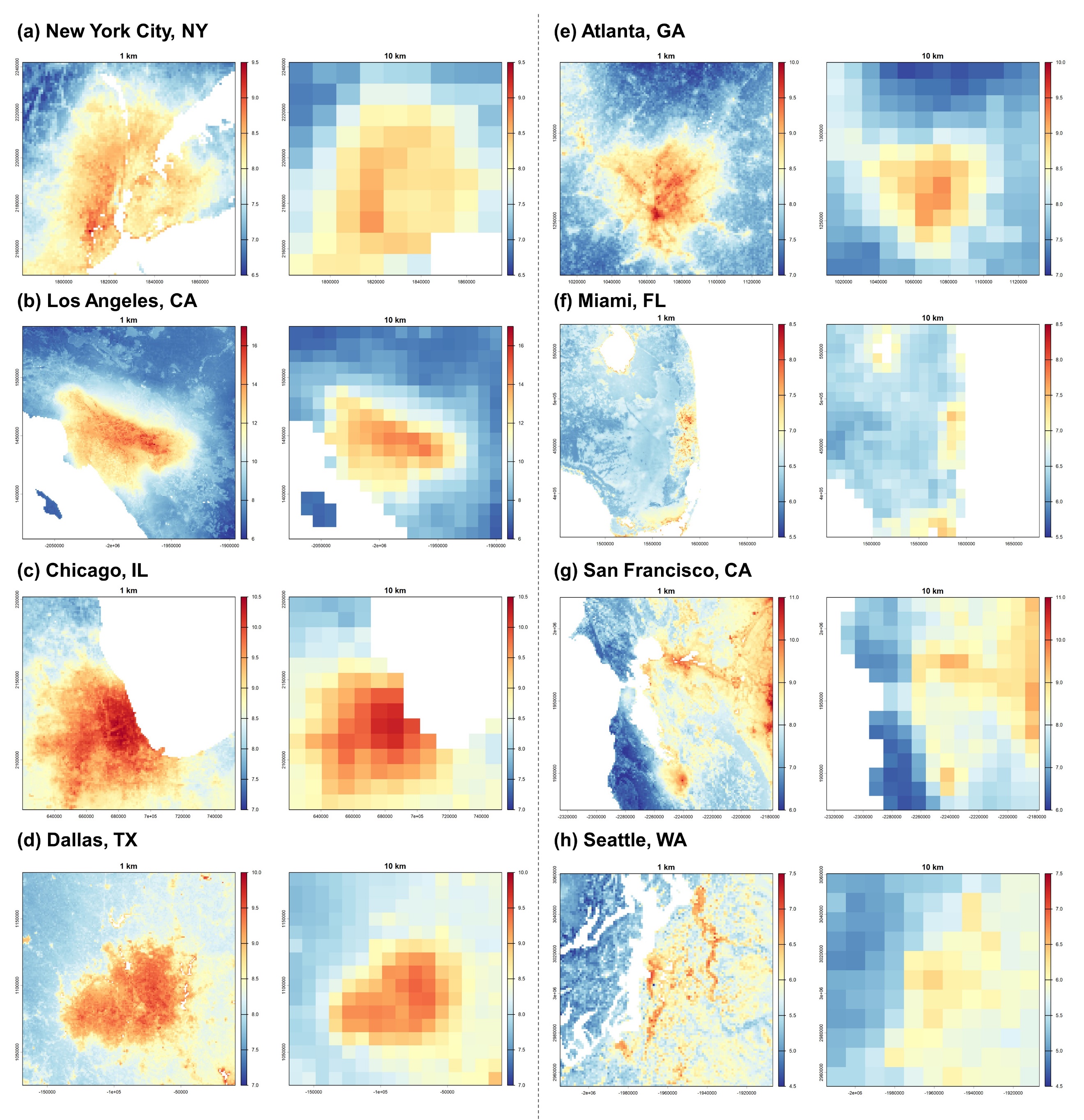

**Figure S9.** Temporal trend of population-weighted annual average non-wildfire and wildfire PM_2.5_ exposure in nine divisions between 2018 and 2024.

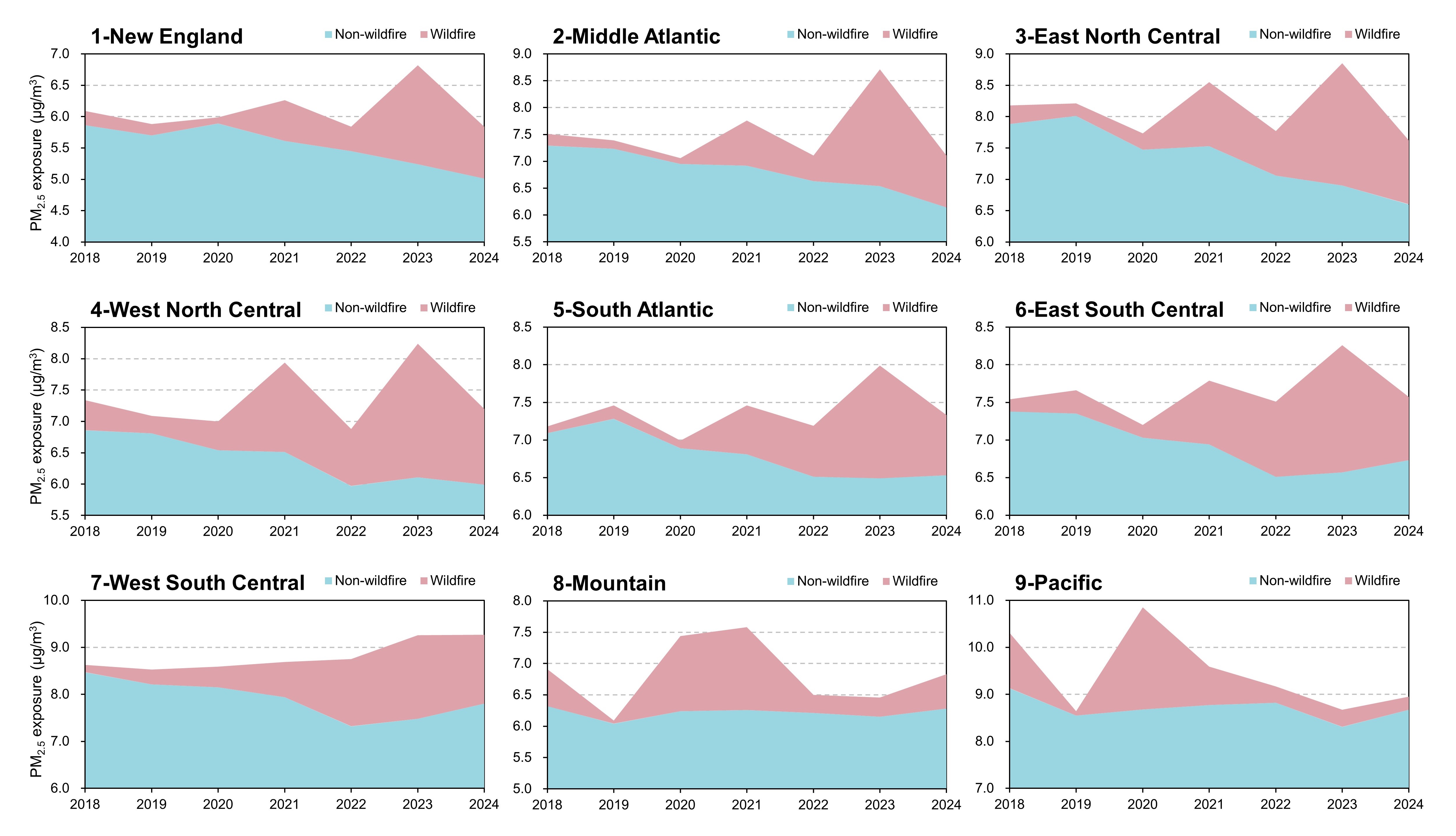

13.	Carnell, R. lhs: Latin Hypercube Samples (R package version 1.2.0). <https://github.com/bertcarnell/lhs>

14.	Greenwell, B. fastshap: Fast Approximate Shapley Values (R package version 0.1.1). <https://github.com/bgreenwell/fastshap>

15.	Greenwell, B. M., pdp: An R Package for Constructing Partial Dependence Plots. *R J* **2017,** *9*, (1), 421-436.

16.	Walker, K., tigris: An R Package to Access and Work with Geographic Data from the US Census Bureau. *R J* **2016,** *8*, (2), 231-242.

17.	Walker, K.; Herman, M. tidycensus: Load US Census Boundary and Attribute Data as 'tidyverse' and 'sf'-Ready Data Frames (R package version 1.7.5). <https://walker-data.com/tidycensus/>

18.	U.S. Environmental Protection Agency Outdoor air quality data. <https://aqs.epa.gov/aqsweb/airdata/download_files.html> (March 18, 2026),

19.	Keller, C. A.; Knowland, K. E.; Duncan, B. N.; Liu, J. H.; Anderson, D. C.; Das, S.; Lucchesi, R. A.; Lundgren, E. W.; Nicely, J. M.; Nielsen, E.; Ott, L. E.; Saunders, E.; Strode, S. A.; Wales, P. A.; Jacob, D. J.; Pawson, S., Description of the NASA GEOS Composition Forecast Modeling System GEOS-CF v1.0. *J Adv Model Earth Sy* **2021,** *13*, (4), e2020MS002413.

20.	Lyapustin, A.; Wang, Y. MODIS/Terra+Aqua Land Aerosol Optical Depth Daily L2G Global 1km SIN Grid V061. <https://developers.google.com/earth-engine/datasets/catalog/MODIS_061_MCD19A2_GRANULES?hl=en>

21.	Copernicus Atmosphere Monitoring Service CAMS global emission inventories. <https://doi.org/10.24381/1d158bec>

22.	Muñoz Sabater, J.; Comyn-Platt, E.; Hersbach, H.; Bell, B.; Berrisford, P.; Biavati, G.; Horányi, A.; Muñoz Sabater, J.; Nicolas, J.; Peubey, C.; Radu, R.; Rozum, I.; Schepers, D.; Simmons, A.; Soci, C.; Dee, D.; Thépaut, J.-N.; Cagnazo, C.; Cucchi, M. ERA5-land post-processed daily-statistics from 1950 to present. <https://doi.org/10.24381/cds.e9c9c792>

23.	Hersbach, H.; Bell, B.; Berrisford, P.; Hirahara, S.; Horányi, A.; Muñoz-Sabater, J.; Nicolas, J.; Peubey, C.; Radu, R.; Schepers, D.; Simmons, A.; Soci, C.; Abdalla, S.; Abellan, X.; Balsamo, G.; Bechtold, P.; Biavati, G.; Bidlot, J.; Bonavita, M.; De Chiara, G.; Dahlgren, P.; Dee, D.; Diamantakis, M.; Dragani, R.; Flemming, J.; Forbes, R.; Fuentes, M.; Geer, A.; Haimberger, L.; Healy, S.; Hogan, R. J.; Hólm, E.; Janisková, M.; Keeley, S.; Laloyaux, P.; Lopez, P.; Lupu, C.; Radnoti, G.; de Rosnay, P.; Rozum, I.; Vamborg, F.; Villaume, S.; Thépaut, J. N., The ERA5 global reanalysis. *Q J Roy Meteor Soc* **2020,** *146*, (730), 1999-2049.

24.	Bondarenko, M.; Priyatikanto, R.; Tejedor-Garavito, N.; Zhang, W.; McKeen, T.; Cunningham, A.; Woods, T.; Hilton, J.; Cihan, D.; Nosatiuk, B.; Brinkhoff, T.; Tatem, A.; Sorichetta, A. Constrained estimates of 2015-2030 total number of people per grid square at a resolution of 30 arc (approximately 1km at the equator) R2025A version v1. <https://doi.org/10.5258/SOTON/WP00840>

25.	Colorado School of Mines VIIRS Nighttime Day/Night Band Composites Version 1. <https://developers.google.com/earth-engine/datasets/catalog/NOAA_VIIRS_DNB_MONTHLY_V1_VCMCFG?hl=en#description> (March 18, 2026),

26.	Didan, K. MODIS/Terra Vegetation Indices Monthly L3 Global 1km SIN Grid V061. <https://doi.org/10.5067/MODIS/MOD13A3.061>

27.	U.S. Geological Survey Annual NLCD (National Land Cover Database)—The next generation of land cover mapping: U.S. Geological Survey Fact Sheet 2025–3001. <https://www.sciencebase.gov/catalog/item/6810c1a4d4be022940554075>

28.	U.S. Census Bureau TIGER/Line Shapefiles. <https://www.census.gov/geographies/mapping-files/time-series/geo/tiger-line-file.html>

29.	Jarvis, A.; Reuter, H. I.; Nelson, A.; Guevara, E. Hole-filled SRTM for the globe Version 4. <https://srtm.csi.cgiar.org> (March 18, 2026),

30.	National Oceanic and Atmospheric Administration Hazard mapping system fire and smoke product. <https://www.ospo.noaa.gov/products/land/hms.html#data> (March 18, 2026),

31.	Di, Q.; Amini, H.; Shi, L. H.; Kloog, I.; Silvern, R.; Kelly, J.; Sabath, M. B.; Choirat, C.; Koutrakis, P.; Lyapustin, A.; Wang, Y. J.; Mickley, L. J.; Schwartz, J., An ensemble-based model of PM2.5 concentration across the contiguous United States with high spatiotemporal resolution. *Environ Int* **2019,** *130*, 104909.

32.	Just, A. C.; Arfer, K. B.; Rush, J.; Lyapustin, A.; Kloog, I., XIS-PM2.5: A daily spatiotemporal machine-learning model for PM2.5 in the contiguous United States. *Environ Res* **2025,** *271*, 120948.

33.	Wang, Z.; Crooks, J. L.; Regan, E. A.; Karimzadeh, M., High-Resolution Estimation of Daily PM2.5 Levels in the Contiguous US Using Bi-LSTM with Attention. *Remote Sens (Basel)* **2025,** *17*, (1), 126.

34.	Kayastha, S. G.; Ghahremanloo, M.; Park, J.; Singh, D.; Westenbarge, D.; Choi, Y., A Deep Learning Framework for Satellite-Derived Surface PM2.5 Estimation: Enhancing Spatial Analysis in the United States. *Artif Intell Earth S* **2024,** *3*, (4), e240028.
